# Mobility network modeling explains higher SARS-CoV-2 infection rates among disadvantaged groups and informs reopening strategies

**DOI:** 10.1101/2020.06.15.20131979

**Authors:** Serina Chang, Emma Pierson, Pang Wei Koh, Jaline Gerardin, Beth Redbird, David Grusky, Jure Leskovec

## Abstract

**Fine-grained epidemiological modeling of the spread of SARS-CoV-2—capturing who is infected at which locations—can aid the development of policy responses that account for heterogeneous risks of different locations as well as the disparities in infections among different demographic groups. Here, we develop a metapopulation SEIR disease model that uses dynamic mobility networks, derived from US cell phone data, to capture the hourly movements of millions of people from local neighborhoods (census block groups, or CBGs) to points of interest (POIs) such as restaurants, grocery stores, or religious establishments. We simulate the spread of SARS-CoV-2 from March 1–May 2, 2020 among a population of 98 million people in 10 of the largest US metropolitan statistical areas. We show that by integrating these mobility networks, which connect 57k CBGs to 553k POIs with a total of 5.4 billion hourly edges, even a relatively simple epidemiological model can accurately capture the case trajectory despite dramatic changes in population behavior due to the virus. Furthermore, by modeling detailed information about each POI, like visitor density and visit length, we can estimate the impacts of fine-grained reopening plans: we predict that a small minority of “superspreader” POIs account for a large majority of infections, that reopening some POI categories (like full-service restaurants) poses especially large risks, and that strategies restricting maximum occupancy at each POI are more effective than uniformly reducing mobility. Our models also predict higher infection rates among disadvantaged racial and socio-economic groups solely from differences in mobility: disadvantaged groups have not been able to reduce mobility as sharply, and the POIs they visit (even within the same category) tend to be smaller, more crowded, and therefore more dangerous. By modeling who is infected at which locations, our model supports fine-grained analyses that can inform more effective and equitable policy responses to SARS-CoV-2**.

## Introduction

In response to the SARS-CoV-2 crisis, numerous stay-at-home orders were enacted across the United States in order to reduce contact between individuals and slow the spread of the virus.^1^ As of May 2020, these orders are being relaxed, businesses are beginning to reopen, and mobility is increasing, causing concern among public officials about the potential resurgence of cases.^2^ Epidemiological models that can capture the effects of changes in mobility on virus spread are a powerful tool for evaluating the effectiveness and equity of various strategies for reopening or responding to a resurgence. In particular, findings of SARS-CoV-2 “superspreader” events^3–7^ motivate models that can reflect the heterogeneous risks of visiting different locations, while well-reported racial and socioeconomic disparities in infection rates^8–14^ require models that can explain the disproportionate impact of the virus on disadvantaged demographic groups.

To address these needs, we construct a fine-grained dynamic mobility network using US cell phone geolocation data from March 1–May 2, 2020. This network maps the hourly movements of millions of people from different census block groups (CBGs), which are geographical units that typically contain 600–3,000 people, to more than half a million specific points of interest (POIs), which are non-home locations that people visit such as restaurants, grocery stores, and religious establishments. (Table S1 provides the 50 POI categories accounting for the largest fraction of visits in this data.) On top of this dynamic bipartite network, we overlay a metapopulation SEIR disease model with only three free parameters that accurately tracks the infection trajectories of each CBG over time as well as the POIs at which these infections are likely to have occurred. The key idea is that combining even a relatively simple epidemiological model with our finegrained, dynamic mobility network allows us to not only accurately model the case trajectory, but also identify the most risky POIs; the most at-risk populations; and the impacts of different reopening policies. This builds upon prior work that models disease spread using mobility data, which has used aggregate^15–21^, historical^22–24^, or synthetic^25–27^ mobility data; separately, other work has directly analyzed mobility data and the effects of mobility reductions in the context of SARS-CoV-2, but without an underlying epidemiological model of disease spread.^28–33^

We use our model to simulate the spread of SARS-CoV-2 within 10 of the largest metropolitan statistical areas (MSAs) in the US, starting from a low, homogeneous prevalence of SARS-CoV-2 across CBGs. For each MSA, we examine the infection risks at individual POIs, the effects of past stay-at-home policies, and the effects of reopening strategies that target specific types of POIs. We also analyze disparities in infection rates across racial and socioeconomic groups, assess the disparate impacts of reopening policies on these groups, and identify mobility-related mechanisms driving these disparities. We find that people from lower-income CBGs have not reduced mobility as sharply, and tend to visit POIs which, even within the same category, are smaller, more crowded, and therefore more dangerous.

## Results

### Mobility network modeling

#### Mobility network

We use geolocation data from SafeGraph, a data company that aggregates anonymized location data from mobile applications, to study mobility patterns from March 1–May 2, 2020 among a population of 98 million people in 10 of the largest US metropolitan statistical areas (MSAs). For each MSA, we represent the movement of individuals between census block groups (CBGs) and points of interest (POIs, defined as specific point locations that people visit such as restaurants, hotels, parks, and stores) as a bipartite network with time-varying edges, where the weight of an edge between a CBG and POI is the number of visitors from that CBG to that POI at a given hour (Figure 1a). SafeGraph also provides the area in square feet of each individual POI, as well as its North American Industry Classification System (NAICS) category (e.g., fitness center or full-service restaurant). We validated the SafeGraph data by comparing to Google mobility data (SI Section S1), and used iterative proportional fitting^34^ to derive hourly POI-CBG networks from the raw SafeGraph data. Overall, these networks comprise 5.4 billion hourly edges between 56,945 CBGs and 552,758 POIs (Extended Data Table 1).

**Figure 1:**
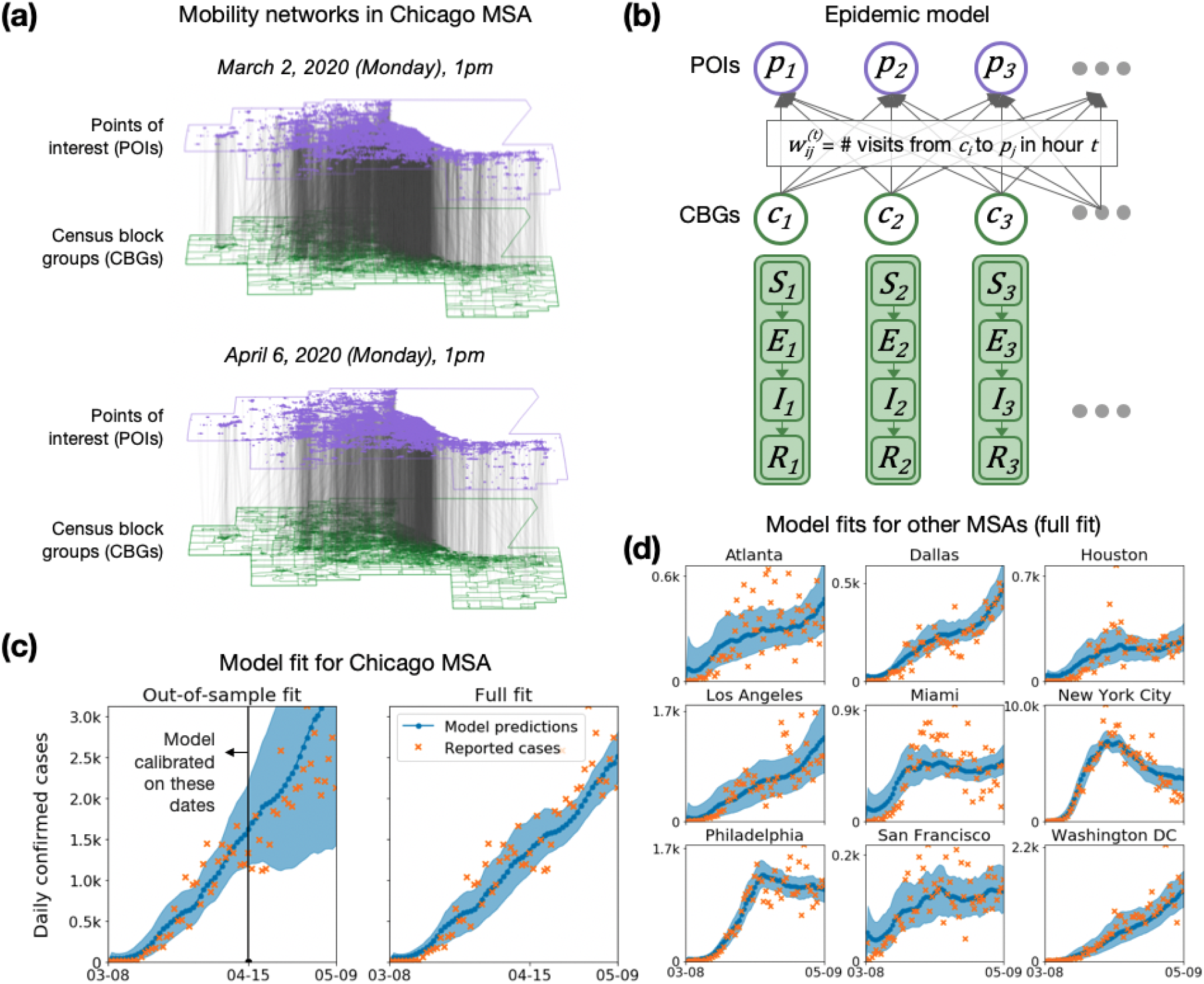
Model description and fit. **(a)** The mobility network captures hourly visits from each census block group (CBG) to each point of interest (POI). The vertical lines indicate that most visits are between nearby POIs and CBGs. Visits dropped dramatically from March (top) to April (bottom), as indicated by the lower density of grey lines. **(b)** We overlaid an SEIR disease model on the mobility network, with each CBG having its own set of SEIR compartments. New infections occur at both POIs and CBGs. The model for each MSA has three free parameters, which remain fixed over time, scaling transmission rates at POIs; transmission rates at CBGs; and the initial fraction of infected individuals. To determine the transmission rate at a given time at each POI we use the mobility network, which captures population movements as well as visit duration and the POI physical area, to estimate the density of visitors at each POI. **(c)** Left: To test out-of-sample prediction, we calibrated the model on data before April 15, 2020 (vertical black line). Even though its parameters remain fixed over time, the model accurately predicts the case trajectory after April 15 by using mobility data. Shaded regions denote 2.5th and 97.5th percentiles across sampled parameters and stochastic realizations. Right: Model fit further improved when we calibrated the model on the full range of data. **(d)** We fit separate models to 10 of the largest US metropolitan statistical areas (MSAs), modeling a total population of 98 million people; here, we show full model fits, as in (c)-Right. While we use the Chicago MSA as a running example throughout the paper, we include results for all other MSAs in the SI.

#### Model

We overlay a SEIR disease model on each mobility network,^15,22^ where each CBG maintains its own susceptible (S), exposed (E), infectious (I), and removed (R) states (Figure 1b). New infections occur at both POIs and CBGs, with the mobility network governing how subpopulations from different CBGs interact as they visit POIs. We use the inferred density of infectious individuals at each POI to determine its transmission rate. The model has only three free parameters, which scale (1) transmission rates at POIs, (2) transmission rates at CBGs, and (3) the initial proportion of infected individuals. All three parameters remain constant over time. We calibrate a separate model for each MSA using confirmed case counts from the *The New York Times*.^35^

#### Model validation

Our models accurately fit observed daily incident case counts in all 10 MSAs from March 8–May 9, 2020 (Figure 1c,d). Additionally, models only calibrated on case counts from March 8–April 14 could predict case counts reasonably well on the held-out time period from April 15–May 9, 2020 (Figure 1c and Extended Data Figure 1a). Our key technical result is that the fine-grained mobility network allows even this relatively simple SEIR model to fit real case trajectories with just three free parameters which remain fixed over time, despite changing social distancing policies and behaviors during that period.

To assess the importance of the detailed mobility network, we tested two alternate models: an aggregate mobility model that uses the total number of POI visits in each hour without taking into account the type of POI or the CBGs from which visitors originate; and a baseline model that does not use mobility data at all. Our network model substantially outperformed both the aggregate mobility model and the baseline model on out-of-sample prediction (Extended Data Figure 1). Furthermore, both alternate models predict very similar rates of infection across all CBGs, which does not concord with previous work showing substantial heterogeneity in infection rates across neighborhoods.^8–14^ This includes higher rates of infection among disadvantaged racial and socioeconomic groups, which our model captures but the alternate models fail to reflect; we discuss this later in Figure 3. These results demonstrate that our network model can better recapitulate observed trends than an aggregate mobility model or a model that does not use mobility data, while also allowing us to assess fine-grained questions like the effects of POI-specific reopening policies.

### Evaluating mobility reduction and reopening policies

We can estimate the impact of a wide range of mobility reduction and reopening policies by applying our model to a modified mobility network that reflects the expected effects of a hypothetical policy. We start by studying the effect of the magnitude and timing of mobility reduction policies from March 2020. We then assess several fine-grained reopening plans, such as placing a maximum occupancy cap or only reopening certain categories of POIs, by leveraging the detailed information that the mobility network contains on each POI, like its average visit length and visitor density at each hour.

#### The magnitude of mobility reduction is as important as its timing

US population mobility dropped sharply in March 2020 in response to SARS-CoV-2; for example, overall mobility in the Chicago MSA fell by 54.8% between the first week of March and the first week of April 2020. We constructed counterfactual mobility networks by scaling the magnitude of mobility reduction down and by shifting the timeline of this mobility reduction earlier and later (Figure 2a), and used our model to simulate the resulting infection trajectories. As expected, shifting the onset of mobility reduction earlier decreased the predicted number of infections incurred, and shifting it later or reducing the magnitude of reduction both increased predicted infections. What was notable was that reducing the magnitude of reduction resulted in far larger increases in predicted infections than shifting the timeline later (Figure 2a). For example, if only a quarter of mobility reduction had occurred in the Chicago MSA, the predicted number of infections would have increased by 3.3× (95% CI, 2.8-3.8), compared to a 1.5 × (95% CI, 1.4-1.6) increase had people begun reducing their mobility one full week later. We observe similar trends across other MSAs (Tables S2 and S3). Our results concord with earlier findings that mobility reductions can dramatically reduce infections.^21,36^

**Figure 2:**
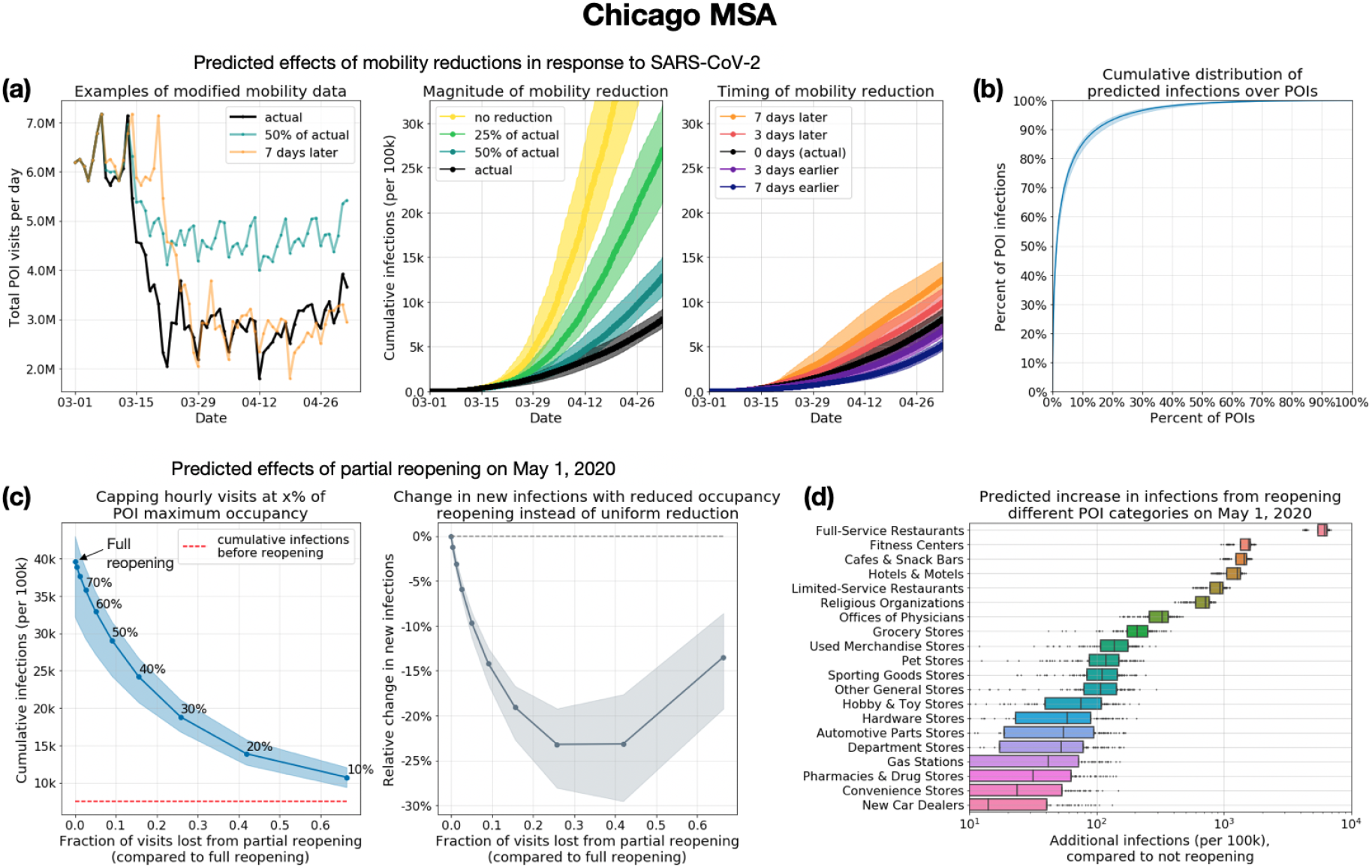
Assessing mobility reduction and reopening policies. **(a)** Counterfactual simulations (left) of the mobility reduction in March 2020—scaling its magnitude down, or shifting the timeline earlier or later—illustrate that the magnitude of mobility reduction (middle) was at least as important as its timing (right). Shaded regions denote 2.5th and 97.5th percentiles across sampled parameters and stochastic realizations. **(b)** Most infections at POIs occur at a small fraction of “superspreader” POIs: 10% of POIs account for more than 80% of the total infections that occurred at POIs in the Chicago MSA (results for other MSAs in Extended Data Figure 3). **(c)** Left: We simulated partial reopening by capping hourly visits if they exceeded a fraction of each POI's maximum occupancy. We plot cumulative infections at the end of one month of reopening against the fraction of visits lost by partial instead of full reopening; the annotations within the plot show the fraction of maximum occupancy used as the cap. Full reopening leads to an additional 32% of the population becoming infected by the end of the month, but capping at 20% maximum occupancy cuts down new infections by more than 80%, while only losing 42% of overall visits. Right: Compared to partially reopening by uniformly reducing visits, the reduced occupancy strategy—which disproportionately targets high-risk POIs with sustained high occupancy—always results in a smaller increase in infections for the same number of visits. The y-axis plots the relative difference between the increase in cumulative infections (from May 1 to May 31) under the reduced occupancy strategy as compared to the uniform reduction strategy. **(d)** We simulated reopening each POI category while keeping reduced mobility levels at all other POIs. Boxes indicate the interquartile range across parameter sets and stochastic realizations. Reopening full-service restaurants has the largest predicted impact on infections, due to the large number of restaurants as well as their high visit densities and long dwell times.

#### A minority of POIs account for a majority of infections

Since overall mobility reduction reduces infections, we next investigated if *how* we reduce mobility—i.e., to which POIs—matters. Using the observed mobility networks to simulate the infection trajectory from March 1–May 2, 2020, we computed the number of expected infections that occurred at each POI and found that a majority of predicted infections occurred at a small fraction of “superspreader” POIs; e.g., in the Chicago MSA, 10% of POIs accounted for 85% (95% CI, 83%-87%) of the predicted infections at POIs (Figure 2b; Extended Data Figure 3 shows similar results across MSAs; across the 10 MSAs, the top 10% of POIs accounted for between 75% and 96% of infections at POIs). These “superspreader” POIs are smaller and more densely occupied, and their occupants stay longer, suggesting that it is especially important to reduce transmission at these high-risk POIs. For example, in the Chicago MSA, the median number of hourly visitors per square foot was 3.2× higher for the riskiest 10% of POIs than for the remaining POIs (0.0041 versus 0.0012 visitors/foot^2^); the median dwell time was 3.5× higher (81 versus 23 minutes). Note that infections at POIs represent a majority, but not all, of the total predicted infections, since we also model infections within CBGs; across MSAs, the median proportion of total predicted infections that occur at POIs is 70%.

#### Reducing mobility by capping maximum occupancy

We simulated the effects of two reopening strategies, implemented beginning on May 1, on the increase in infections by the end of May. First, we evaluated a reduced occupancy reopening strategy, in which hourly visits to each POI return to those in the first week of March (prior to widespread adoption of stay-at-home measures), but are capped if they exceed a fraction of the POI’s maximum occupancy,^37^ which we estimated as the maximum hourly number of visitors ever recorded at that POI. A full return to early March mobility levels without reducing maximum occupancy produces a spike in predicted infections: in the Chicago MSA, we project that an additional 32% (95% CI, 25%-35%) of the population will be infected within a month (Figure 2c). However, capping maximum occupancy substantially reduces risk without sharply reducing overall mobility: capping at 20% maximum occupancy in the Chicago MSA cuts down new infections by more than 80% but only loses 42% of overall visits, and we observe similar trends across other MSAs (Extended Data Figure 4). This highlights the non-linearity of infections as a function of visits: one can achieve a disproportionately large reduction in infections with a small reduction in visits.

We also compared the reduced occupancy strategy to a baseline that uniformly reduces visits to each POI from their levels in early March. Reduced occupancy always results in fewer infections for the same total number of visits: e.g., capping at 20% maximum occupancy reduces new infections by 23% (95% CI, 18%-30%), compared to the uniform baseline for the same total number of visits in the Chicago MSA (Figure 2c). This is because reduced occupancy takes advantage of the heterogeneous risks across POIs, disproportionately reducing visits at high-risk POIs with sustained high occupancy, but allowing lower-risk POIs to return fully to prior mobility levels.

#### Relative risk of reopening different categories of POIs

We assessed the relative risk of re-opening different categories of POIs by reopening each category in turn on May 1 (and returning its mobility patterns to early March levels) while keeping mobility patterns at all other POIs at their reduced, stay-at-home levels (Figure 2d). Following prior work,^30^ we excluded several categories of POIs from this analysis, including schools and hospitals, because of concerns that the cell phone mobility dataset might not contain all POIs in the category or capture all relevant risk factors; see Methods M6. We find a large variation in reopening risks: on average across the 10 MSAs (Extended Data Figure 5), full-service restaurants, gyms, hotels, cafes, religious organizations, and limited-service restaurants produce the largest increases in infections when reopened. Reopening full-service restaurants is particularly risky: in the Chicago MSA, we predict an additional 596k (95% CI, 434k-686k) infections by the end of May, more than triple the next riskiest POI category. These risks are the total risks summed over all POIs in the category, but the relative risks after normalizing by the number of POIs are broadly similar, with restaurants, gyms, hotels, cafes, and religious establishments predicted to be the most dangerous on average per individual POI (Extended Data Figure 5). These categories are more dangerous because their POIs tend to have higher visit densities and/or visitors stay there longer (Figures S10-S19).

### Infection disparities between socioeconomic and racial groups

We characterize the differential spread of SARS-CoV-2 along demographic lines by using US Census data to annotate each CBG with its racial composition and median income, then tracking how infection disparities arise across groups. We use this approach to study the mobility mechanisms behind disparities and to quantify how different reopening strategies impact disadvantaged groups.

#### Mobility patterns contribute to disparities in infection rates

Despite only having access to mobility data and no other demographic information, our models correctly predicted higher risks of infection among disadvantaged racial and socioeconomic groups.^8–14^ Across all MSAs, individuals from CBGs in the bottom decile for income were substantially likelier to have been infected by the end of the simulation, even though all individuals began with equal likelihoods of infection in our simulation (Figure 3a). This overall disparity was driven primarily by a few POI categories (e.g., full-service restaurants), which infected far larger proportions of lower-income CBGs than higher-income CBGs (Figure 3c; similar trends hold across all MSAs in Figure S1). We similarly found that CBGs with fewer white residents had higher relative risks of infection, although results were more variable across MSAs (Figure 3b). Our models also recapitulated known associations between population density and infection risk^38^ (median Spearman correlation between CBG density and cumulative incidence proportion, 0.39 across MSAs), despite not being given any information on population density. In SI Section S2, we confirm that the magnitude of the disparities our model predicts are generally consistent with real-world disparities and further explore the large predicted disparities in Philadelphia, which stem from substantial differences in visit densities at the POIs that are frequented by visitors from different socioeconomic and racial groups. In the analysis below, we focus on the mechanisms producing higher relative risks of infection among lower-income CBGs, and we show in Extended Data Figure 6 and Table S5 that similar results hold for racial disparities as well.

**Figure 3:**
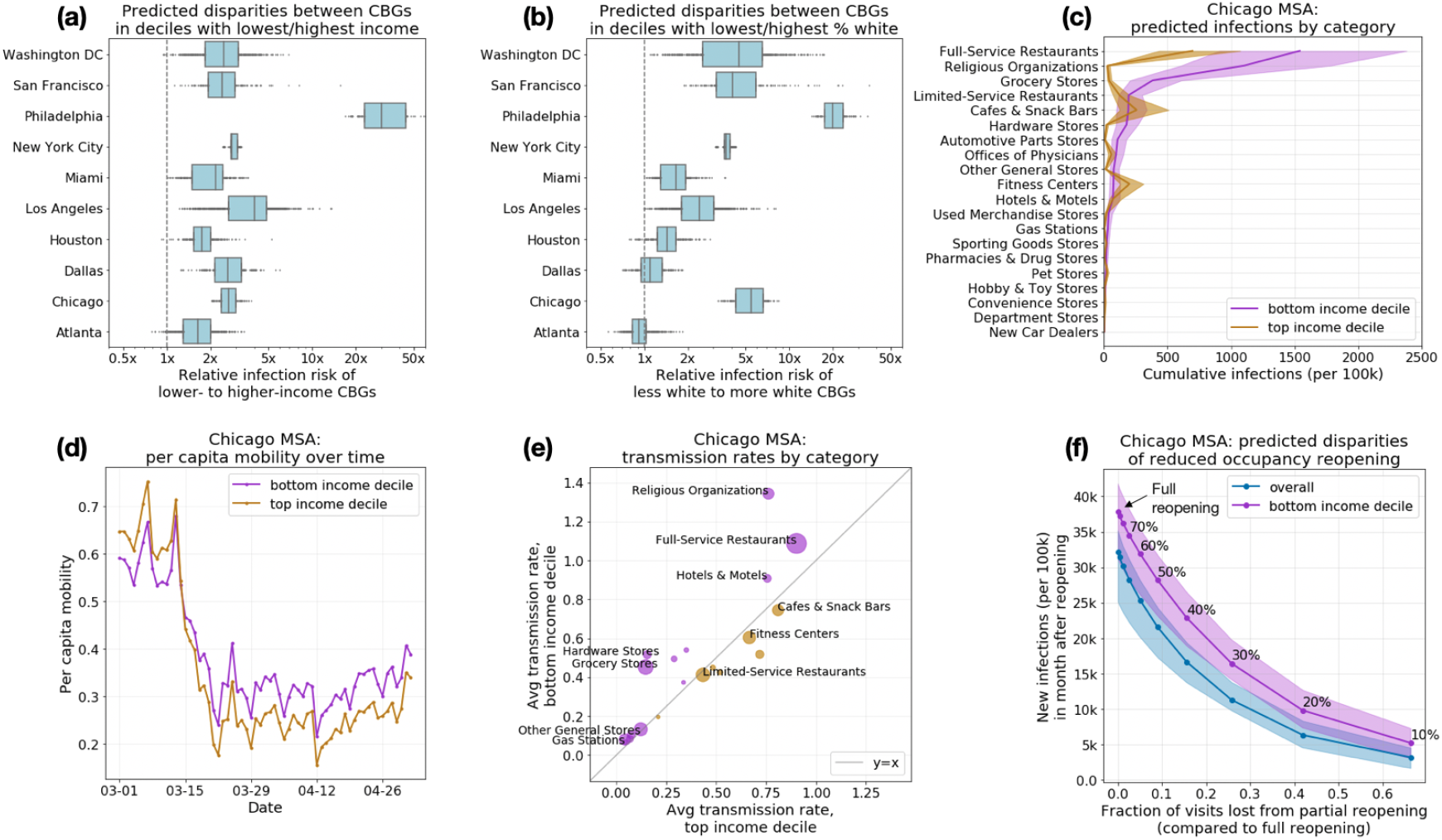
Mobility patterns give rise to socioeconomic and racial disparities in infections. (**a**) Across all MSAs, our model predicts that people in lower-income census block groups (CBGs) are more likely to be infected, even though they start with equal probabilities of being infected. Disparities are especially prominent in Philadelphia, which we discuss in SI Section S2. Boxes indicate the interquartile range across parameter sets and stochastic realizations. (**b**) Racial disparities are similar: people in non-white CBGs are typically more likely to be infected, although results are more variable across MSAs. (c-e) illustrate how mobility patterns give rise to socioeconomic disparities; similar mechanisms underlie racial disparities (Extended Data Figure 6, Table S5). (**c**) The overall disparity is driven by a few POI categories like full-service restaurants. Shaded regions denote 2.5th and 97.5th percentiles across sampled parameters and stochastic realizations. (**d**) One reason for the disparities is that higher-income CBGs were able to reduce their overall mobility levels below those of lower-income CBGs. (**e**) Within each category, people from lower-income CBGs tend to visit POIs that are smaller and more crowded and therefore have higher transmission rates. Thus, even if a lower-income and a higher-income person went out equally often and went to the same types of places, the lower-income person would still have a greater risk of infection. The size of each dot indicates the total number of visits to that category. (**f**) We predict the effect of reopening (at different levels of reduced occupancy) on different demographic groups. Reopening leads to more infections in lower-income CBGs (purple) than the overall population (blue), underscoring the need to account for disadvantaged subpopulations when assessing reopening plans.

#### Lower-income CBGs saw smaller reductions in mobility

Across all MSAs, we found that lower-income CBGs did not reduce their mobility as sharply in the first few weeks of March 2020, and had higher mobility than higher-income CBGs for most of March through May (Figure 3d, Extended Data Figure 6). For example, over the month of April, lower-income CBGs in the Chicago MSA had 27% more POI visits per capita than higher-income CBGs. Differences in mobility patterns within categories partially explained the within-category infection disparities: e.g., lower-income CBGs made substantially more visits per capita to grocery stores than did higher-income CBGs, and consequently experienced more infections at that category (Extended Data Figure 7).

#### POIs visited by lower-income CBGs tend to have higher transmission rates

Differences in the number of visits per capita between lower- and higher-income CBGs do not fully explain the infection disparities: for example, Cafes & Snack Bars were visited more frequently by people from higher-income CBGs in every MSA (Extended Data Figure 7), but they caused more predicted infections among people from lower-income CBGs in the majority of MSAs (Figure S1). We found that even within a POI category, the transmission rate at POIs frequented by people from lower-income CBGs tended to be higher than the corresponding rate for higher-income CBGs (Figure 3e; Table S4), because these POIs tended to be smaller and more crowded. It follows that, even if a lower-income and higher-income person had the same mobility patterns and went to the same types of places, the lower-income person would still have a greater risk of infection.

As a case study, we examine grocery stores in further detail. In 8 of the 10 MSAs, visitors from lower-income CBGs encountered higher transmission rates at grocery stores than those from higher-income CBGs (median transmission rate ratio of 2.19, Table S4). Why was one visit to the grocery store twice as dangerous for a lower-income individual? Taking medians across MSAs, we found that the average grocery store visited by lower-income individuals had 59% more hourly visitors per square foot, and their visitors stayed 17% longer on average. These findings highlight how fine-grained differences in mobility patterns—how often people go out, which categories of places they go to, which POIs they choose within those categories—can ultimately contribute to dramatic disparities in infection outcomes.

#### Reopening plans must account for disparate impact

Because disadvantaged groups suffer a larger burden of infection, it is critical to not just consider the overall impact of reopening plans but also their disparate impact on disadvantaged groups specifically. For example, our model predicted that full reopening in the Chicago MSA would result in an additional 39% (95% CI, 31%-42%) of the population of CBGs in the bottom income decile being infected within a month, compared to 32% (95% CI, 25%-35%) of the overall population (Figure 3f; results for all MSAs in Extended Data Figure 4). Similarly, Extended Data Figure 8 illustrates that reopening individual POI categories tends to have a larger impact on the bottom income decile. More conservative reopening plans produce smaller absolute disparities in infections—e.g., we predict that reopening at 20% of maximum occupancy would result in infections among an additional 6% (95% CI, 4%-8%) of the overall population and 10% (95% CI, 7%-13%) of the population in CBGs in the bottom income decile (Figure 3f)—though the relative disparity remains.

## Discussion

We model the spread of SARS-CoV-2 using a dynamic mobility network that encodes the hourly movements of millions of people between 57k neighborhoods (census block groups, or CBGs) and 553k points of interest (POIs). Because our data contains detailed information on each POI, like visit length and visitor density, we can estimate the impacts of fine-grained reopening plans—predicting that a small minority of “superspreader” POIs account for a large majority of infections, that reopening some POI categories (like full-service restaurants) poses especially large risks, and that strategies that restrict the maximum occupancy at each POI are more effective than uniformly reducing mobility. Because we model infections in each CBG, we can infer the approximate demographics of the infected population, and thereby assess the disparate socioeconomic and racial impacts of SARS-CoV-2. Our model correctly predicts that disadvantaged groups are more likely to become infected, and also illuminates two mechanisms that drive these disparities: (1) disadvantaged groups have not been able to reduce their mobility as dramatically (consistent with previously-reported data, and likely in part because lower-income individuals are more likely to have to leave their homes to work^10^) and (2) when they go out, they visit POIs which, even within the same category, are smaller, more crowded, and therefore more dangerous.

The cell phone mobility dataset we use has limitations: it does not cover all populations (e.g., prisoners, children under 13, or adults without smartphones), does not contain all POIs (e.g., nursing homes are undercovered, and we exclude schools and hospitals from our analysis of POI category risks), and cannot capture sub-CBG heterogeneity in demographics. Individuals may also be double-counted in the dataset if they carry multiple cell phones. These limitations notwithstanding, cell phone mobility data in general and SafeGraph data in particular have been instrumental and widely used in modeling SARS-CoV-2 spread.^15–17,28–32,39^ Our model itself is parsimonious, and does not include such relevant features as asymptomatic transmission; variation in household size; travel and seeding between MSAs; differentials in susceptibility due to pre-existing conditions or access to care; age-related variation in mortality rates or susceptibility (e.g., for modeling transmission at elementary and secondary schools); various time-varying transmission-reducing behaviors (e.g., hand-washing, mask-wearing); and some POI-specific risk factors (e.g., ventilation). Although our model recovers case trajectories and known infection disparities even without incorporating these features, we caution that this predictive accuracy does not mean that our predictions should be interpreted in a narrow causal sense. Because certain types of POIs or subpopulations may disproportionately select for certain types of omitted processes, our findings on the relative risks of different POIs should be interpreted with due caution. However, the predictive accuracy of our model suggests that it broadly captures the relationship between mobility and transmission, and we thus expect our broad conclusions—e.g., that people from lower-income CBGs have higher infection rates in part because because they tend to visit smaller, denser POIs and because they have not been able to reduce mobility by as much (likely in part because they cannot as easily work from home^10^)—to hold robustly.

Our results can guide policymakers seeking to assess competing approaches to reopening and tamping down post-reopening resurgence. Despite growing concern about racial and socioeconomic disparities in infections and deaths, it has been difficult for policymakers to act on those concerns; they are currently operating without much evidence on the disparate impacts of reopening policies, prompting calls for research which both identifies the causes of observed disparities and suggests policy approaches to mitigate them.^11,14,40,41^ Our fine-grained mobility modeling addresses both these needs. Our results suggest that infection disparities are not the unavoidable consequence of factors that are difficult to address in the short term, like disparities in preexisting conditions; on the contrary, short-term policy decisions substantially affect infection disparities by altering the overall amount of mobility allowed, the types of POIs reopened, and the extent to which POI occupancies are clipped. Considering the disparate impact of reopening plans may lead policymakers to, e.g., (1) favor more conservative reopening plans, (2) increase testing in disadvantaged neighborhoods predicted to be high risk (especially given known disparities in access to tests^8^), and (3) prioritize distributing masks and other personal protective equipment to disadvantaged populations who have not reduced their mobility as much and frequent riskier POIs.

As society reopens and we face the possibility of a resurgence in cases, it is critical to build models which allow for fine-grained assessments of the effects of reopening policies. We hope that our approach, by capturing heterogeneity across POIs, demographic groups, and cities, helps address this need.

## Data Availability

Census data, case and death counts from The New York Times, and Google mobility data are publicly available. Cell phone mobility data is freely available to researchers, non-profits, and governments through the SafeGraph COVID-19 Data Consortium (https://www.safegraph.com/covid-19-data-consortium).

https://github.com/nytimes/covid-19-data

https://google.com/covid19/mobility/

https://census.gov/programssurveys/acs

## Methods

The Methods section is structured as follows. We describe the datasets we use in Methods M1 and the mobility network that we derive from these datasets in Methods M2. In Methods M3, we discuss the SEIR model we overlay on the mobility network; in Methods M4, we describe how we calibrate this model and quantify uncertainty in its predictions; in Methods M5, we introduce a series of sensitivity analyses and robustness checks that we performed to further validate our model. In Methods M6, we provide details on the experimental procedures used for our analysis of physical distancing, reopening, and demographic disparities. Finally, in Methods M7, we elaborate on how we estimate the mobility network from the raw mobility data.

### M1 Datasets

#### SafeGraph

We use geolocation data provided by SafeGraph, a data company that aggregates anonymized location data from numerous mobile applications. We obtained IRB exemption for SafeGraph data from the Northwestern University IRB office. SafeGraph data captures the movement of people between census block groups (CBGs), which are geographical units that typically contain a population of between 600 and 3,000 people, and points of interest (POIs) like restaurants, grocery stores, or religious establishments. Specifically, we use the following SafeGraph datasets:

1. Places Patterns^44^ and Weekly Patterns (v1)^45^. These datasets contain, for each POI, hourly counts of the number of visitors, estimates of median visit duration in minutes (the “dwell time”), and aggregated weekly and monthly estimates of visitors’ home CBGs. We use visitor home CBG data from the Places Patterns dataset, as described below: for privacy reasons, SafeGraph excludes a home CBG from this dataset if fewer than 5 devices were recorded at the POI from that CBG over the course of the month. For each POI, SafeGraph also provides their North American Industry Classification System (NAICS) category, and an estimate of their physical area in square feet. (Area is computed using the footprint polygon SafeGraph assigns to the POI.^46,47^) We analyze Places Patterns data from January 1, 2019 to February 29, 2020 and Weekly Patterns data from March 1, 2020 to May 2, 2020.
2. Social Distancing Metrics,^48^ which contains hourly estimates of the proportion of people staying home in each CBG. We analyze Social Distancing Metrics data from March 1, 2020 to May 2, 2020.

We focus on 10 of the largest metropolitan statistical areas (MSAs) in the US (Extended Data Table 1). We chose these MSAs by taking a random subset of the SafeGraph Patterns data and picking the 10 MSAs with the most POIs in the data. Our methods in this paper can be straightforwardly applied, in principle, to the other MSAs in the original SafeGraph data. For each MSA, we include all POIs that meet all of the following requirements: (1) the POI is located in the MSA; (2) SafeGraph has visit data for this POI for every hour that we model, from 12am on March 1, 2020 to 11pm on May 2, 2020; (3) SafeGraph has recorded the home CBGs of this POI’s visitors for at least one month from January 2019 to February 2020; (4) the POI is not a “parent” POI. “Parent” POIs comprise a small fraction of POIs in the dataset which overlap and include the visits from their “child” POIs: for example, many malls in the dataset are parent POIs which include the visits from stores which are their child POIs. To avoid double-counting visits, we remove all parent POIs from the dataset.

After applying these POI filters, we include all CBGs that have at least 1 recorded visit to at least 10 of the remaining POIs; this means that CBGs from outside the MSA may be included if they visit this MSA frequently enough. Summary statistics of the post-processed data are in Extended Data Table 1. Overall, we analyze 57k CBGs from the 10 MSAs, and over 310M visits from these CBGs to over 552k POIs.

SafeGraph data has been used to study consumer preferences^49^ and political polarization.^50^ More recently, it has been used as one of the primary sources of mobility data in the US for tracking the effects of the SARS-CoV-2 pandemic.^28,30,51–53^ In SI Section S1, we show that aggregate trends in SafeGraph mobility data broadly match up to aggregate trends in Google mobility data in the US,^54^ before and after the imposition of stay-at-home measures. Previous analyses of SafeGraph data have shown that it is geographically representative: for example, it does not systematically over-represent individuals from CBGs in different counties or with different racial compositions, income levels, or educational levels.^55,56^

#### US Census

Our data on the demographics of census block groups (CBGs) comes from the US Census Bureau’s American Community Survey (ACS).^57^ We use the 5-year ACS (2013-2017) to extract the median household income, proportion of white residents, and proportion of black residents of each CBG. For the total population of each CBG, we use the most recent one-year estimates (2018); one-year estimates are noisier but we wish to minimize systematic downward bias in our total population counts (due to population growth) by making them as recent as possible.

#### New York Times

We calibrate our models using the SARS-CoV-2 dataset published by the *The New York Times*.^35^ Their dataset consists of cumulative counts of cases and deaths in the United States over time, at the state and county level. For each MSA that we model, we sum over the county-level counts to produce overall counts for the entire MSA. We convert the cumulative case and death counts to daily case and death counts for the purposes of model calibration, as described below.

### M2 Mobility network

We consider a complete undirected bipartite graph 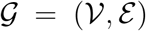 with time-varying edges. The vertices 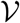 are partitioned into two disjoint sets 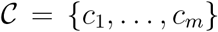, representing m census block groups (CBGs), and 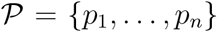, representing n points of interest (POIs). The weight 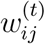 on an edge (*c_i_*,*p_j_*) at time *t* represents our estimate of the number of individuals from CBG *c_i_* visiting POI *p_j_* at the *t*-th hour of simulation. We record the number of edges (with non-zero weights) in each MSA and over all hours from March 1, 2020 to May 2, 2020 in Extended Data Table 1. Across all 10 MSAs, we study 5.4 billion edges between 56,945 CBGs and 552,758 POIs.

From US Census data, each CBG c_i_ is labeled with its population 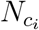, income distribution, and racial and age demographics. From SafeGraph data, each POI *p_j_* is similarly labeled with its category (e.g., restaurant, grocery store, or religious organization), its physical size in square feet 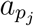, and the median dwell time *d_pj_* of visitors to *p_j_*.

The central technical challenge in constructing this network is estimating the network weights 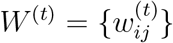 from SafeGraph data, since this visit matrix is not directly available from the data. Because the estimation procedure is involved, we defer describing it in detail until Methods M7; in Methods M3–M6, we will assume that we already have the network weights.

### M3 Model dynamics

To model the spread of SARS-CoV-2, we overlay a metapopulation disease transmission model on the mobility network defined in Methods M2. The transmission model structure follows prior work on epidemiological models of SARS-CoV-2^15,22^ but incorporates a fine-grained mobility network into the calculations of the transmission rate (Methods M3.1). We construct separate mobility networks and models for each metropolitan statistical area (MSA).

We use a SEIR model with susceptible (*S*), exposed (*E*), infectious (*I*), and removed (*R*) compartments. Susceptible individuals have never been infected, but can acquire the virus through contact with infectious individuals, which may happen at POIs or in their home CBG. They then enter the exposed state, during which they have been infected but are not infectious yet. Individuals transition from exposed to infectious at a rate inversely proportional to the mean latency period. Finally, they transition into the removed state at a rate inversely proportional to the mean infectious period. The removed state represents individuals who cannot infect others, because they have recovered, self-isolated, or died.

Each CBG q maintains its own SEIR instantiation, with 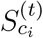, 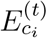, 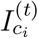, and 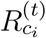 representing how many individuals in CBG *c_i_* are in each disease state at hour *t*, and 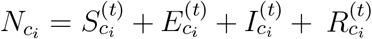. At each hour *t*, we sample the transitions between states as follows:

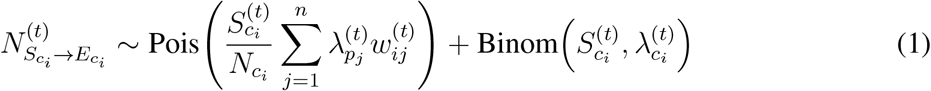

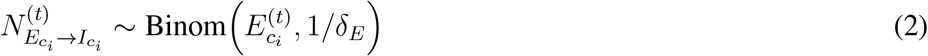

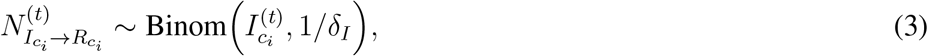

where 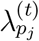 is the rate of infection at POI *p_j_* at time *t*; 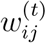, the *ij*-th entry of the visit matrix from the mobility network (Methods M2), is the number of visitors from CBG *c_i_* to POI *p_j_* at time *t*; 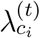 is the base rate of infection that is independent of visiting POIs; *δ_E_* is the mean latency period; and *δ_I_*, is the mean infectious period.

We then update each state to reflect these transitions. Let 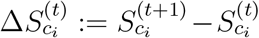, and likewise for 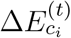, 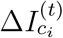, and 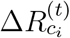. Then,

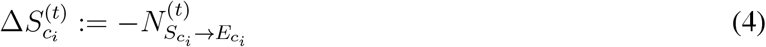

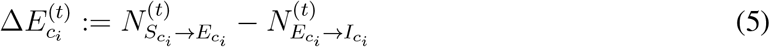

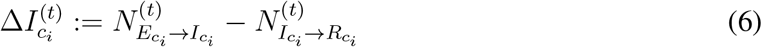

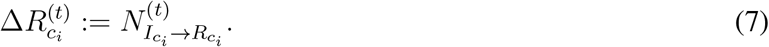

#### M3.1 The number of new exposures 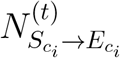

We separate the number of new exposures 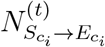 in CBG *c_i_* at time *t* into two parts: cases from visiting POIs, which are sampled from 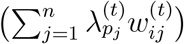, and other cases not captured by visiting POIs, which are sampled from 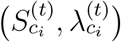.

##### New exposures from visiting POIs

We assume that any susceptible visitor to POI *p_j_* at time *t* has the same independent probability 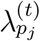 of being infected and transitioning from the susceptible (*S*) to the exposed (*E*) state. Since there are 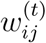 visitors from CBG *c_i_* to POI *p_j_* at time *t*, and we assume that a 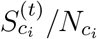 fraction of them are susceptible, the number of new exposures among these visitors is distributed as Binom 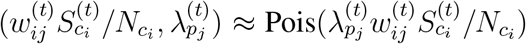. The number of new exposures among all outgoing visitors from CBG *c_i_* is therefore distributed as the sum of the above expression over all POIs, 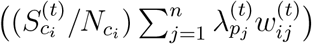.

We model the infection rate at POI *p_j_* at time *t*, 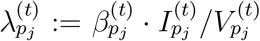, as the product of its transmission rate 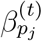 and proportion of infectious individuals 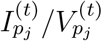, where 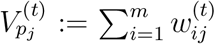 is the total number of visitors to *p_j_* at time *t*,

We model the transmission rate at POI *p_j_* at time *t* as

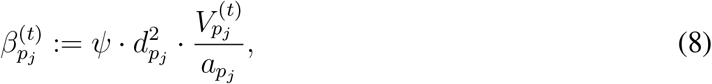

where *a_p_j__* is the physical area of *p_j_*, and *ψ* is a transmission constant (shared across all POIs) that we fit to data. The inverse scaling of transmission rate with area *a_p_j__* is a standard simplifying assumption.^42^ The dwell time fraction *d_p_j__* ∊ [0,1] is what fraction of an hour an average visitor to *p_j_* at any hour will spend there (Methods M7.1); it has a quadratic effect on the POI transmission rate 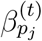 because it reduces both (1) the time that a susceptible visitor spends at *p_j_* and (2) the density of visitors at *p_j_*.

With this expression for the transmission rate 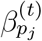, we can calculate the infection rate at POI *p_j_* at time *t* as

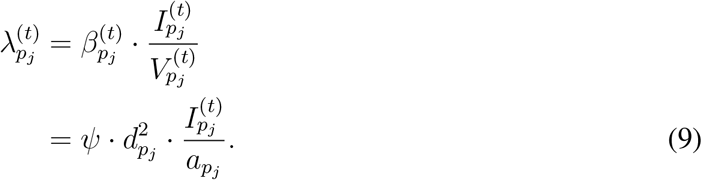

For sufficiently large values of *ψ* and a sufficiently large proportion of infected individuals, the expression above can sometimes exceed 1. To address this, we simply clip the infection rate to 1. However, this occurs very rarely for the parameter settings and simulation duration that we use.

Finally, to compute the number of infectious individuals at *p_j_* at time *t*, 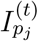, we assume that the proportion of infectious individuals among the 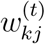 visitors to *p_j_* from a CBG *c_k_* mirrors the overall density of infections 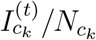 in that CBG, although we note that the scaling factor *ψ* can account for differences in the ratio of infectious individuals who visit POIs. This gives

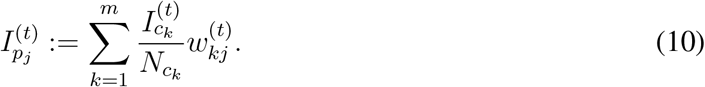

##### Base rate of new exposures not captured by visiting POIs

In addition to the new exposures from infections at POIs, we model a CBG-specific base rate of new exposures that is independent of POI visit activity. This captures other sources of infections, e.g., household infections or infections at POIs that are absent from the SafeGraph data. We assume that at each hour, every susceptible individual in CBG *c_i_* has a 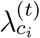 probability of becoming infected and transitioning to the exposed state, where

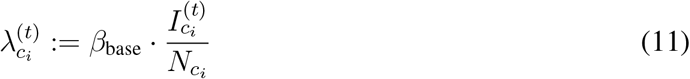

is proportional to the infection density at CBG *c_i_*, and *β*_base_ is a constant that we fit to data.

##### Overall number of new exposures

Putting all of the above together yields the expression for the distribution of new exposures in CBG *c_i_* at time *t*,

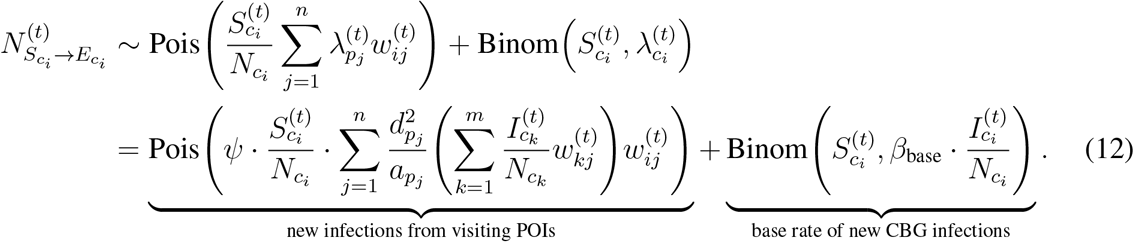

#### M3.2 The number of new infectious and removed cases

We model exposed individuals as becoming infectious at a rate inversely proportional to the mean latency period *δ_E_*. At each time step *t*, we assume that each exposed individual has a constant, time-independent probability of becoming infectious, with

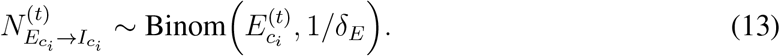

Similarly, we model infectious individuals as transitioning to the removed state at a rate inversely proportional to the mean infectious period *δ_I_*, with

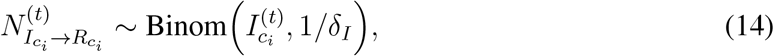

We estimate both *δ_E_* and *δ_I_*, from prior literature; see Methods M4.

#### M3.3 Model initialization

In our experiments, *t* = 0 is the first hour of March 1, 2020. We approximate the infectious *I* and removed *R* compartments at *t* = 0 as initially empty, with all infected individuals in the exposed *E* compartment. We further assume the same expected initial prevalence *p*_0_ in every CBG *c_i_*. At *t* = 0, every individual in the MSA has the same independent probability *p*_0_ of being exposed *E* instead of susceptible *S*. We thus initialize the model state by setting

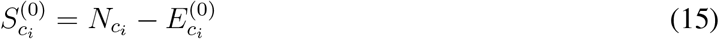

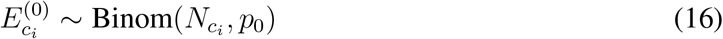

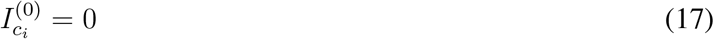

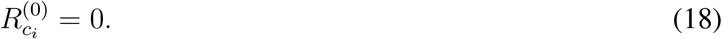

### M4 Model calibration and validation

Most of our model parameters can either be estimated from SafeGraph and US Census data, or taken from prior work (see Extended Data Table 2 for a summary). This leaves 3 model parameters that do not have direct analogues in the literature, and that we therefore need to calibrate with data:

1. The transmission constant in POIs, *ψ* (Equation (9))
2. The base transmission rate, *β*_base_ (Equation (11))
3. The initial proportion of exposed individuals at time *t* = 0, *p*_0_ (Equation (16)).

In this section, we describe how we fit these parameters to published numbers of confirmed cases, as reported by *The New York Times*. We fit models for each MSA separately. In Methods M4.4, we show that the resulting models can accurately predict the number of confirmed cases in out-of sample data that was not used for model fitting.

#### M4.1 Selecting parameter ranges

##### Transmission rate factors *ψ* and *β*_base_

We select parameter ranges for the transmission rate factors *ψ* and *β*_base_ by checking if the model outputs match plausible ranges of the basic reproduction number *R*_0_ pre-lockdown, since *R*_0_ has been the study of substantial prior work on SARS-CoV-2.^58^ Under our model, we can decompose *R*_0_ = *R*_base_ + *R*_POI_, where *R*_POI_ describes transmission due to POIs and *R*_base_ describes the remaining transmission (as in Equation (12)). We first establish plausible ranges for *R*_base_ and *R*_POI_ before translating these into plausible ranges for *β*_base_ and *ψ*.

We assume that *R*_base_ ranges from 0.1–2. *R*_base_ models transmission that is not correlated with activity at POIs in the SafeGraph dataset, including within-household transmission and transmission at POI categories (like subways or nursing homes) which are not well-captured in the SafeGraph dataset. We chose the lower limit of 0.1 because beyond that point, base transmission would only contribute minimally to overall *R*, whereas previous work suggests that within-household transmission is a substantial contributor to overall transmission.^59–61^ Household transmission alone is not estimated to be sufficient to tip overall *R*_0_ above 1; for example, a single infected individual has been estimated to cause an average of 0.32 (0.22, 0.42) secondary within-household infections.^59^. However, because *R*_base_ may also capture transmission at POIs not captured in the SafeGraph dataset, to be conservative, we chose an upper limit of *R*_base_ = 2; as we describe below, the best-fit models for all 10 MSAs have *R*_base_ < 2, and 9 out of 10 have *R*_base_ < 1. We allow *R*_POI_ to range from 1–3, which corresponds to allowing *R*_0_ = *R*_POI_ + *R*_base_ to range from 1.1-5. This is a conservatively wide range, since prior work estimates a pre-lockdown *R*_0_ of 2–3.^58^

To determine the values of *R*_base_ and *R*_POI_ that a given pair of *β*_base_ and *ψ* imply, we seeded a fraction of index cases and then ran the model on looped mobility data from the first week of March to capture pre-lockdown conditions. We initialized the model by setting *p*_0_, the initial proportion of exposed individuals at time *t* = 0, to *p*_0_ = 10^−4^, and then sampling in accordance with Equation (16). Let *N*_0_ be the number of initial exposed individuals sampled. We computed the number of individuals that these *N*_0_ index cases went on to infect through base transmission, *N*_base_, and POI transmission, *N*_POI_, which gives

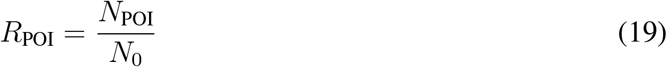

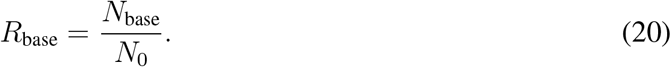

We averaged these quantities over stochastic realizations for each MSA. Figure S2 shows that, as expected, *R*_base_ is linear in *β*_base_ and *R*_POI_ is linear in *ψ*. *R*_base_ lies in the plausible range when *β*_base_ ranges from 0.0012–0.024, and *R*_POI_ lies in the plausible range (for at least one MSA) when *ψ* ranges from 515–4,886, so these are the parameter ranges we consider when fitting the model. As described in Methods M4.2, we verified that case count data for all MSAs can be fit using parameter settings for *β*_base_ and *ψ* within these ranges.

##### Initial prevalence of exposures, *p*_0_

The extent to which SARS-CoV-2 infections had spread in the U.S. by the start of our simulation (March 1, 2020) is currently unclear.^62^ To account for this uncertainty, we allow *p*_0_ to vary across a large range between 10^−5^ and 10^−2^. As described in Methods M4.2, we verified that case count data for all MSAs can be fit using parameter settings for *p*_0_ within this range.

#### M4.2 Fitting to the number of confirmed cases

Using the parameter ranges above, we grid searched over *ψ*, *β*_base_, and *p*_0_ to find the models that best fit the number of confirmed cases reported by *The New York Times* (NYT).^35^ For each of the 10 MSAs studied, we tested 1,500 different combinations of *ψ*, *β*_base_, and *p*_0_ in the parameter ranges specified above, with parameters linearly spaced for *ψ* and *β*_base_ and logarithmically spread for *p*_0_.

In Methods M3, we directly model the number of infections but not the number of confirmed cases. To estimate the number of confirmed cases, we assume that an *r_c_* = 0.1 proportion of infections will be confirmed, and moreover that they will confirmed exactly *δ_c_* = 168 hours (7 days) after becoming infectious; these parameters are estimated from prior work (Extended Data Table 2). From these assumptions, we can calculate the predicted number of newly confirmed cases across all CBGs in the MSA on day *d*,

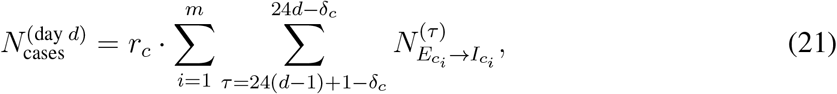

where *m* indicates the total number of CBGs in the MSA and for convenience we define 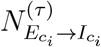 the number of newly infectious people at hour *τ*, to be 0 when *τ* < 1.

From NYT data, we have the reported number of new cases 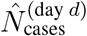 for each day *d*, summed over each county in the MSA. We compare the reported number of cases and the number of cases that our model predicts by computing the root-mean-squared-error (RMSE) between each of the 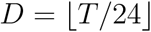 days of our simulations,

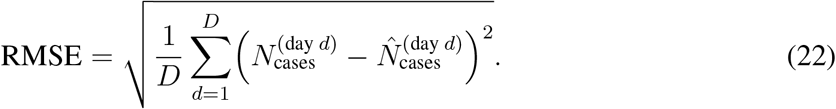

For each combination of model parameters and for each MSA, we quantify model fit with the NYT data by running 30 stochastic realizations and averaging their RMSE. Note that we measure model fit based on the daily number of new reported cases (as opposed to the cumulative number of reported cases).^63^

Our simulation spans March 1 to May 2, 2020, and we use mobility data from that period. However, because we assume that cases will be confirmed *δ_C_* = 7 days after individuals become infectious (Extended Data Table 2), we predict the number of cases with a 7 day offset, from March 8 to May 9, 2020.

#### M4.3 Parameter selection and uncertainty quantification

Throughout this paper, we report aggregate predictions from different parameter sets of *ψ*, *β*_base_, and *p*_0_ and multiple stochastic realizations. For each MSA, we:

1. Find the best-fit parameter set, i.e., with the lowest average RMSE over stochastic realizations.
2. Select all parameter sets that achieve an RMSE (averaged over stochastic realizations) within 20% of the RMSE of the best-fit parameter set.
3. Pool together all predictions across those parameter sets and all of their stochastic realizations, and report their mean and 2.5th/97.5th percentiles.

On average, each MSA has 9.7 parameter sets that achieve an RMSE within 20% of the best-fitting parameter set (Table S8). For each parameter set, we have results for 30 stochastic realizations. All uncertainty intervals in our results show the 2.5th/97.5th percentiles across these pooled results.

This procedure corresponds to rejection sampling in an Approximate Bayesian Computation framework,^15^ where we assume an error model that is Gaussian with constant variance; we pick an acceptance threshold based on what the best-fit model achieves; and we use a uniform parameter grid instead of sampling from a uniform prior. It quantifies uncertainty from two sources. First, the multiple realizations capture stochastic variability between model runs with the same parameters. Second, simulating with all parameter sets that are within 20% of the RMSE of the best fit captures uncertainty in the model parameters *ψ*, *β*_base_, and *p*_0_. The latter is equivalent to assuming that the posterior probability over the true parameters is uniformly spread among all parameter sets within the 20% threshold.

#### M4.4 Model validation on out-of-sample cases

We validate our models by showing that they predict the number of confirmed cases on out-of-sample data when we have access to corresponding mobility data. For each MSA, we split the available NYT dataset into a training set (spanning March 8, 2020 to April 14, 2020) and a test set (spanning April 15, 2020 to May 9, 2020). We fit the model parameters *ψ*, *β*_base_, and *p*_0_, as described in Methods M4.2, but only using the training set. We then evaluate the predictive accuracy of the resulting model on the test set. When running our models on the test set, we still use mobility data from the test period. Thus, this is an evaluation of whether the models can accurately predict the number of cases, given mobility data, in a time period that was not used for model calibration. Extended Data Figure 1a shows that the models fit the out-of-sample case data fairly well, demonstrating that they can extrapolate beyond the training set to future time periods. Note that we only use this train/test split to evaluate out-of-sample model accuracy. All other results are generated using parameter sets that best fit the *entire* dataset, as described in Methods M4.2.

### M5 Sensitivity analyses and robustness checks

#### M5.1 Aggregate mobility and no-mobility baseline models

##### Comparison to aggregate mobility model

Our model uses a detailed mobility network to simulate disease spread. To test if this detailed model is necessary, or if our model is simply making use of aggregate mobility patterns, we tested an alternate SEIR model that uses the aggregate number of visits made to any POI in the MSA in each hour, but not the breakdown of visits between specific CBGs to specific POIs. Like our model, the aggregate mobility model also captures transmission due to POI visits and mixing within CBGs; thus, the two models have the same three free parameters (*ψ*, scaling transmission rates at POIs; *β*_base_, scaling transmission rates at CBGs; and *p*_0_, the initial fraction of infected individuals).

As in our network model, transmission under the aggregate mobility model happens at POIs and at CBGs. For POI transmission, we take the probability that a susceptible person (from any CBG) will become infected due to a POI visit at time *t* as equal to

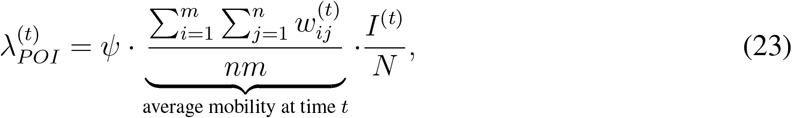

where *m* is the number of CBGs, *n* is the number of POIs, *I*^(^*^t^*^)^ is the total number of infectious individuals at time *t*, and *N* is the total population size of the MSA. For CBG transmission, we assume the same process as in our network model: the probability 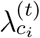 that a susceptible person in CBG *c_i_* will become infected in their CBG in time t is equal to, *β*_base_ times the current infectious fraction of *c_i_* (Equation 11). Putting it together, the aggregate mobility model defines the number of new exposures in CBG *c_i_* at time *t* as

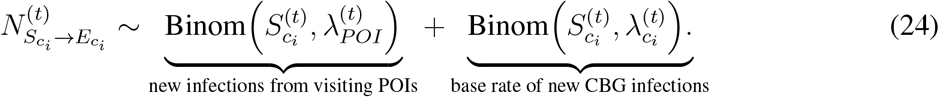

All other dynamics are the same between the aggregate mobility model and our network model, as described in Methods M3. We determined parameter ranges and calibrated the aggregate mobility model in the exact same way as we did for our network model, as described in Methods M4. As discussed in the main text, we found that our network model substantially outperformed the aggregate mobility model in out-of-sample cases prediction (Extended Data Figure 1).

##### Comparison to baseline that does not use mobility data

To determine the extent to which mobility data might aid in modeling the case trajectory, we also compared our model to a baseline SEIR model that does not use mobility data and simply assumes that all individuals within an MSA mix uniformly. In this no-mobility baseline, an individual’s risk of being infected and transitioning to the exposed state at time *t* is

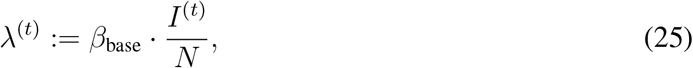

where *I*^(^*^t^*^)^ is the total number of infectious individuals at time *t*, and *N* is the total population size of the MSA. As above, the other model dynamics are identical, and for model calibration we performed a similar grid search over *β*_base_ and *p*_0_. As expected, we found both the network and aggregate mobility models outperformed the no-mobility model on out-of-sample case predictions (Extended Data Figure 1).

#### M5.2 Modifying the parametric form for POI transmission rates

Recall from Methods M3, Equation (8), that in our model, the transmission rate at a POI *p_j_* at an hour *t*,

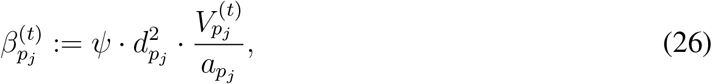

depends on two key ingredients: 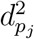, which reflects how much time visitors spend there, and 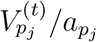, which reflects the density (number of visitors per sq ft) of the POI in that hour. These assumptions are based on prior expectations that a visit is more dangerous for a susceptible individual if they spend more time there and/or if the place is more crowded. To assess empirically the role that each of these two terms play, we compared our transmission rate formula to two perturbed versions of it: one that removed the dwell time term, and another that removed the density term. For each of these formulas, we computed the risk of visiting a POI category as the average transmission rate of the category, with the rate of each POI weighted by the proportion of category visits that went to that POI. Then, we evaluated whether the relative risks predicted by each formula con-corded with the rankings of POI categories proposed by independent epidemiological experts.^64,65^ In our evaluations, we included all of the categories that we analyzed (i.e., the 20 categories with the most visits in SafeGraph data; see Section M6) that overlapped with categories described in the external rankings. To compare against Emanuel et al.^64^, we also converted their categorical groupings into numerical score, i.e., “Low” → 1, “Low/Medium” → 2, etc., up to “High” → 5. Sims et al.^65^ already provided numerical ratings so we did not have to perform a conversion.

As shown in Figure S5, we find that the predicted relative risks match external sources best when we use our original parametric form that accounts for both dwell time and density: restaurants, cafes, religious organizations, and gyms are among the most dangerous, while grocery stores and retail (e.g., clothing stores) are less dangerous. However, when we assume only dwell time matters and remove the density term, we see unrealistic changes in the ranking: e.g., restaurants drop close to grocery stores, despite both sets of experts deeming them far apart in terms of risk. When we assume only density matters and remove dwell time, we also see unrealistic changes: e.g., limited-service restaurants are predicted to be far riskier than full-service restaurants, and gyms and religious organizations are no longer predicted as risky, which contradicts both of our sources. These findings demonstrate that both factors — the dwell time and density — are important toward faithfully modeling transmission at POIs, since the predictions become less realistic when either factor is taken out.

#### M5.3 Parameter identifiability

We assess the identifiability of the fitted model parameters *ψ*, *β*_base_, and *p*_0_ as follows. First, we verify that the model-fitting procedure is able to recover the true parameters when fit on simulated data for which the true parameters are known. For each MSA, we simulate daily case counts using the best-fit parameters for that MSA (i.e., those with the minimum RMSE to daily case counts, as reported in Table S8). We then run our grid search fitting procedure on the simulated case counts. For all 10 MSAs, as Figure S8 illustrates, the parameters in our grid search that obtain the lowest RMSE on the simulated data are always the true parameters that were used to generate that data. This demonstrates that our model and fitting procedure can correctly recover the true parameters on simulated data.

As a further assessment of model identifiability, in Figure S9 we plot RMSE on true (not simulated) daily case counts (that is, the metric used to perform model calibration) as a function of model parameters, *β*_base_ and *ψ*. (We take the minimum RMSE over values of *p_0_* so the plots can be visualized in two dimensions.) As these plots illustrate, *β*_base_ and *ψ* are correlated, which is unsurprising because they scale the growth of infections at CBGs and POIs respectively. We account for the uncertainty caused by this correlation throughout the analysis, by aggregating results from all parameter settings which achieve an RMSE within 20% of the best-fit model for each MSA, as described in Section M4.3.

#### M5.4 Stochastic sampling of cases

Instead of assuming that a fixed proportion of infections are confirmed after a fixed confirmation delay, we also tried stochastically sampling the number of confirmed cases and the confirmation delay. For each day *d*, we first computed 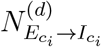, the number of people who became infectious on this day; we then sampled from 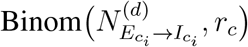 to get the number of confirmed cases that would result from this group of infections. For each case that was to be confirmed, we drew its confirmation delay – i.e., delay from becoming infectious to being confirmed – from distributions fitted on empirical line-list data: either Gamma(1.85, 3.57)^22^ or Exp(6.1).^43^

We found that our model predictions barely changed when we sampled case trajectories stochastically using either delay distribution, as opposed to assuming a fixed confirmation rate and delay (Figure S4). However, an advantage of our fixed method is that it allows us to predict confirmed cases up to *δ_c_* (i.e., 7) days after the last day of simulation, whereas we cannot do the same when we sample confirmed cases and delays stochastically. This is because, if delays are stochastic, predicting the number of confirmed cases on, for example, the 5*^th^* day after the simulation ends depends on the number of newly infectious individuals every day before and including that day, but since the simulation ended days before, the model would not have sufficient information to make the prediction. On the other hand, the fixed method simply translates and scales the newly infectious curve, so we can predict the number of confirmed cases 5 days after the simulation ends, since it only depends on the number of newly infectious individuals 2 days before the end of simulation. Due to this advantage, we opted to use the fixed method, as described in Methods M4.

#### M5.5 Model calibration metrics

Finally, we tested three alternative model calibration procedures using different metrics for measuring when to accept or reject a model parameter setting. For each procedure, we recomputed our downstream analyses and verified that our key results on superspreader POIs (Extended Data Figure 3), the effects of reopening (Figure S6), and group disparities (Figure 3) all remained similar.

##### Poisson likelihood model

Our model calibration procedure, which uses RMSE to assess fit, implicitly assumes that error in the number of observed cases is drawn from a normal (Gaussian) distribution. As a sensitivity analysis, we tested a Poisson error model instead, using negative log-likelihood as a measure of fit, and using the same 20% threshold for model calibration as in Methods M4. We note that the homoscedastic Gaussian model will likely prioritize fitting parts of the case trajectory that have higher case counts, whereas a Poisson model will comparatively prioritize fitting parts of the case trajectory with lower case counts. We found that ranking models via Poisson likelihood was consistent with ranking models using RMSE (both computed on daily incident cases, as described above): the median Spearman correlation over MSAs between models ranked by Poisson likelihood vs. RMSE was 0.97.

##### Model acceptance threshold

As described in Methods M4, we set the acceptance threshold for model calibration (i.e., the threshold for rejection sampling in the Approximate Bayesian Computation framework) to 20% of the RMSE of the best-fit model. We selected this threshold because beyond that point, model fit qualitatively deteriorated based on inspection of the case trajectories. As a sensitivity analysis, we selected a different threshold (10%), corresponding to selecting a subset of the models that had a better fit, and verified that the key results remained similar.

##### Fitting to deaths

In addition to the number of confirmed cases, the NYT data also contains the daily reported number of deaths due to COVID-19 by county. As an additional sensitivity analysis, we calibrated our models to fit this death data instead of case data. To estimate the number of deaths *N*_deaths_, we use a similar process as for the number of cases *N*_cases_, except that we replace *r_c_* with *r_d_* = 0.66%, the estimated infection fatality rate for COVID-19,^66^ and *δ_c_* with *δ_d_* = 432 hours (18 days), the number of days between becoming infectious and dying^66^ (Extended Data Table 2 provides references for all parameters). This gives

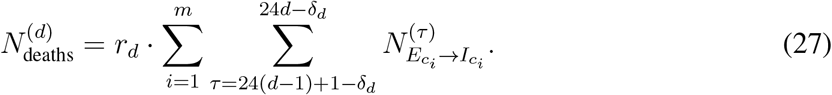

Because we assume that deaths occur *δ_d_* =18 days after individuals become infectious, we compared with NYT death data starting on March 19, 2020 (18 days after our simulation begins). Extended Data Figure 2 shows that while the daily death data is noisy, the calibrated models can also fit the trends in the death counts well. Ranking models using RMSE on deaths was consistent with ranking models using RMSE on cases, with a median Spearman correlation over MSAs of 0.99, and as with the above sensitivity analyses (changing the likelihood model and the acceptance threshold), we found that our key results remained similar.

### M6 Analysis details

In this section, we include additional details about the experiments underlying the figures in the paper. We omit explanations for figures that are completely described in the main text.

#### Comparing the magnitude vs. timing of mobility reduction (Figure 2a)

To simulate what would have happened if we changed the magnitude or timing of mobility reduction, we modify the real mobility networks from March 1–May 2, 2020, and then run our models on the hypothetical data. In Figure 2a, we report the cumulative incidence proportion at the end of the simulation (May 2, 2020), i.e., the total fraction of people in the exposed, infectious, and removed states at that time.

To simulate a smaller magnitude of mobility reduction, we interpolate between the mobility network from the first week of simulation (March 1–7, 2020), which we use to represent typical mobility levels (prior to mobility reduction measures), and the actual observed mobility network for each week. Let *W*^(^*^t^*^)^ represent the observed visit matrix at the *t*-th hour of simulation, and let *f*(*t*) = *t* mod 168 map t to its corresponding hour in the first week of simulation, since there are 168 hours in a week. To represent the scenario where people had committed to a e [0,1] times the actual observed reduction in mobility, we construct a visit matrix 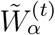 that is an α-convex combination of *W*^(^*^t^*^)^ and *W^f^*^(^*^t^*^)^,

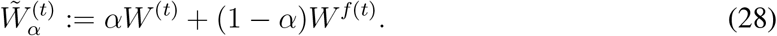

If *α* is 1, then 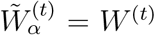, and we use the actual observed mobility network for the simulation. On the other hand, if *α* = 0, then 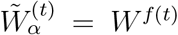, and we assume that people did not reduce their mobility levels at all by looping the visit matrix for the first week of March throughout the simulation. Any other *α* ∈ [0,1] interpolates between these two extremes.

To simulate changing the timing of mobility reduction, we shift the mobility network by *d* ∈ [−7, 7] days. Let *T* represent the last hour in our simulation (May 2, 2020, 11PM), let *f*(*t*) = *t* mod 168 map t to its corresponding hour in the first week of simulation as above, and similarly let *g*(*t*) map t to its corresponding hour in the last week of simulation (April 27-May 2, 2020). We construct the time-shifted visit matrix 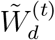

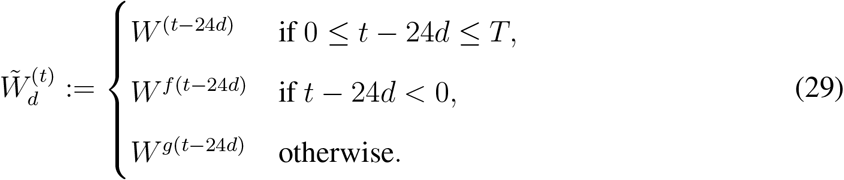

If *d* is positive, this corresponds to starting mobility reduction *d* days later; if we imagine time on a horizontal line, this shifts the time series to the right by 24*d* hours. However, doing so leaves the first 24*d* hours without visit data, so we fill it in by reusing visit data from the first week of simulation. Likewise, if *d* is negative, this corresponds to starting mobility reduction *d* days earlier, and we fill in the last 24*d* hours with visit data from the last week of simulation.

#### A minority of POIs account for a majority of infections (Figure 2b and Extended Data Figure 3)

To evaluate the distribution of infections over POIs, we run our models on the observed mobility data from March 1–May 2, 2020 and record the number of infections that occur at each POI. Specifically, for each hour *t*, we compute the number of expected infections that occur at each POI *p_j_* by taking the number of susceptible people who visit *p_j_* in that hour multiplied by the POI infection rate 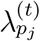 (Equation (9)). Then, we count the total expected number of infections per POI by summing over hours. In Figure 2b, we sort the POIs by their expected number of infections and report the proportion of all infections caused by the top *x*% of POIs.

#### Reducing mobility by clipping maximum occupancy (Figure 2c, Extended Data Figure 4)

We implemented two partial reopening strategies: one that uniformly reduced visits at POIs to a fraction of full activity, and the other that “clipped” each POI’s hourly visits to a fraction of the POI’s maximum occupancy. For each reopening strategy, we started the simulation at March 1, 2020 and ran it until May 31, 2020, using the observed mobility network from March 1–April 30, 2020, and then using a hypothetical post-reopening mobility network from May 1–31, 2020, corresponding to the projected impact of that reopening strategy. Because we only have observed mobility data from March 1–May 2, 2020, we impute the missing mobility data up to May 31, 2020 by looping mobility data from the first week of March, as in the above analysis on the effect of past reductions in mobility. Let *T* represent the last hour for which we have observed mobility data (May 2, 2020, 11PM). To simplify notation, we define

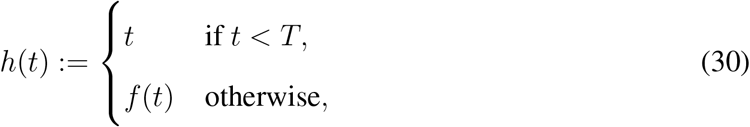

where, as above, *f*(*t*) = *t* mod 168. This function leaves *t* unchanged if there is observed mobility data at time *t*, and otherwise maps *t* to the corresponding hour in the first week of our simulation.

To simulate a reopening strategy that uniformly reduced visits to an *γ*-fraction of their original level, where *γ* ∊ [0,1], we constructed the visit matrix

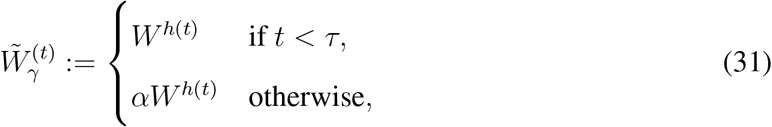

where *τ* represents the first hour of reopening (May 1, 2020, 12AM). In other words, we use the actual observed mobility network up until hour t, and then subsequently simulate an *γ*-fraction of full mobility levels.

To simulate the clipping strategy, we first estimated the maximum occupancy *M_pj_*, of each POI *p_j_* as the maximum number of visits that it ever had in one hour, across all of March 1 to May 2, 2020. As in previous sections, let 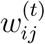 represent the *i*, *j*-th entry in the observed visit matrix *W*^(^*^t^*^)^, i.e., the number of people from CBG *c_i_* who visited *p_j_* in hour *t*, and let 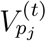 represent the total number of visitors to *p_j_* in that hour, i.e., 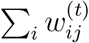. We simulated clipping at a *β*-fraction of maximum occupancy, where *β* ∊ [0,1], by constructing the visit matrix 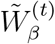 whose *i*, *j*-th entry is

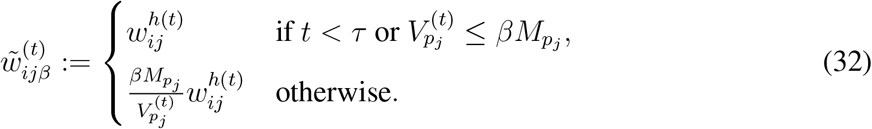

This corresponds to the following procedure: for each POI *p_j_* and time *t*, we first check if *t* < *τ* (reopening has not started) or if 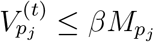 < (the total number of visits to *p_j_* at time *t* is below the allowed maximum *βM_pj_*). If so, we leave 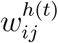 unchanged. Otherwise, we compute the scaling factor 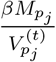 that would reduce the total visits to *p_j_* at time *t* down to the allowed maximum *βM_pj_*, and then scale down all visits from each CBG *c_i_* to *p_j_* proportionately.

For both reopening strategies, we calculate the increase in cumulative incidence at the end of the reopening period (May 31, 2020), compared to the start of the reopening period (May 1, 2020).

#### Relative risk of reopening different categories of POIs (Figure 2d, Extended Data Figures 5 and 8, Figures S10-S19)

We study separately reopening the 20 POI categories with the most visits in SafeGraph data. In this analysis, we exclude four categories, following prior work^30^: “Child Day Care Services” and “Elementary and Secondary Schools” (because children under 13 are not well-tracked by SafeGraph); “Drinking Places (Alcoholic Beverages)” (because SafeGraph seems to undercount these locations) and “Nature Parks and Other Similar Institutions” (because boundaries and therefore areas are not well-defined by SafeGraph). We also exclude “General Medical and Surgical Hospitals” and “Other Airport Operations” (because hospitals and air travel both involve many additional risk factors our model is not designed to capture). We do not filter out these POIs during model fitting, because including them still increases the proportion of overall mobility our dataset captures; we simply do not analyze these categories specifically, because we wish to be conservative and only focus on categories where we are most confident we are fully capturing transmission at the category. For the Drinking Places (Alcoholic Beverages) category, prior work reports that “SafeGraph staff suggest that part of the low count [of drinking places] is due to ambiguity in the division between restaurants and bars and pubs that serve food”,^30^ and some restaurants in the data are indeed described as establishments like bars, beer gardens, breweries, cocktail lounges, or Irish pubs. This suggests that some drinking places that also serve food are already accounted for in our model under restaurants.

This reopening analysis is similar to the above analysis on clipping vs. uniform reopening. As above, we set the reopening time *τ* to May 1, 2020, 12AM. To simulate reopening a POI category, we take the set of POIs in that category, 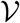, and set their activity levels after reopening to that of the first week of March. For POIs not in the category 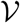, we keep their activity levels after reopening the same, i.e., we simply repeat the activity levels of the last week of our data (April 27-May 2, 2020): This gives us the visit matrix 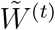 with entries

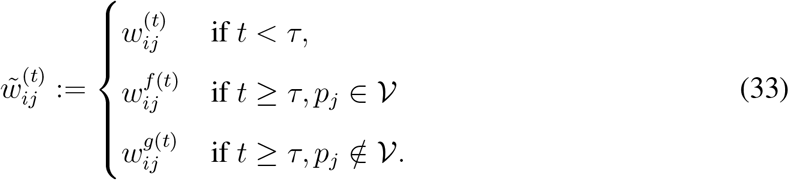

As in the above reopening analysis, *f*(*t*) maps t to the corresponding hour in the first week of March, and *g*(*t*) maps t to the corresponding hour in the last week of our data. For each category, we calculate the difference between (1) the cumulative fraction of people who have been infected by the end of the reopening period (May 31, 2020) and (2) the cumulative fraction of people infected by May 31 had we not reopened the POI category (i.e., if we simply repeated the activity levels of the last week of our data). This seeks to model the increase in cumulative incidence by end of May from reopening the POI category. In Extended Data Figure 5 and Figures S10-S19, the bottom right panel shows the increase for the category as a whole, and the bottom left panel shows the increase *per POI* (i.e., the total increase divided by the number of POIs in the category).

#### Per-capita mobility (Figure 3d, Extended Data Figures 6 and 7)

Each group of CBGs (e.g., the bottom income decile) comprises a set 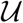 of CBGs that fit the corresponding criteria. In Extended Data 6, we show the daily per-capita mobilities of different pairs of groups (broken down by income and by race). To measure the per-capita mobility of a group on day *d*, we take the total number of visits made from those CBGs to any POI, 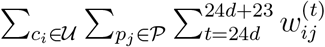, and divide it by the total population of the CBGs in the group, 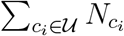. In Extended Data Figure 7, we show the total number of visits made by each group to each POI category, accumulated over the entire data period (March 1–May 2, 2020) and then divided by the total population of the group.

#### Average transmission rate of a POI category (Figure 3e)

We compute the average hourly transmission rate experienced by a group of CBGs 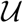 at a POI category 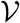 as

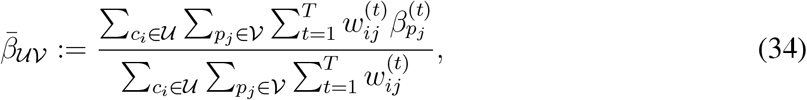

where, as above, 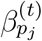 is the transmission rate at POI *p_j_* in hour *t* (Equation (8)), 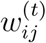 is the number of visitors from CBG *c_i_* at POI *p_j_* in hour *t*, and *T* is the last hour in our simulation. This represents the expected transmission rate encountered during a visit by someone from a CBG in group 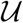 to a POI in category 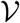.

### M7 Estimating the mobility network from SafeGraph data

Finally, we describe how we estimate the dwell time *d_pj_* (Methods M7.1) and visit matrix *W*^(^*^t^*^)^ (Methods M7.2) from SafeGraph data.

#### Quantities from SafeGraph data

We use the following quantities from SafeGraph data:

- The estimated visit matrix *Ŵ*^(^*^r^*^)^ aggregated for the month *r*, where we use *r* instead of *t* to denote time periods longer than an hour. This is taken from the Patterns dataset, and is aggregated at a monthly level. To account for non-uniform sampling from different CBGs, we weight the number of SafeGraph visitors from each CBG by the ratio of the CBG population and the number of SafeGraph devices with homes in that CBG.^67^
- 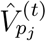: The number of visitors recorded in POI *p_j_* at hour *t*. This is taken from the Weekly Patterns v1 dataset.
- 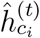: The estimated fraction of people in CBG *c_i_* who left their home in day ⎿t/24⏌. This is derived by taking 1 – `(completely_home_device_count/device_count)`. These are daily (instead of hourly) metrics in the Social Distancing Metrics dataset.
- 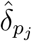: The median length of a visit to a POI *p_j_*. We estimate this by averaging over the weekly values in the `median_dwell` field in the Patterns datasets in March and April 2020. 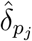, is measured to minute-level resolution and expressed in units of hours, e.g., 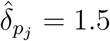 means a median visit time of 1.5 hours = 90 minutes.

#### M7.1 Data preprocessing and dwell time computation

##### Hourly visits

The raw SafeGraph data records the number of visitors that *newly arrive* at each POI *p_j_* at each hour. However, 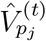 above represents the number of visitors that *are present* at a POI in an hour t; these visitors may have arrived prior to *t*. The aggregate visit matrix *Ŵ*^(^*^r^*^)^, as well as the visit matrix *W*^(^*^t^*^)^ used in our model, are defined similarly. To compute these quantities from the raw data, we make two assumptions: first, that every visitor to *p_j_* stays for exactly 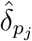, hours, where 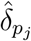, is the median length of a visit to *p_j_*, and second, that a visitor who newly arrives in an hour *t* is equally likely to arrive at any time from [*t*, *t* + 1). With these assumptions, we can convert the number of visitor arrivals in each hour into the expected number of visitors present at each hour: for example, if 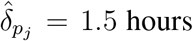, then we assume that a visitor who arrives sometime during an hour *t* will also be present in hour *t* +1 and be present half the time, on expectation, in hour *t* + 2. Note that under our definition, visits are still counted even if a visitor does not stay for the entire hour. For example, a visitor that arrives at 9:30am and leaves at 10:10am will be counted as two visits: one during the 9-10am hour and one during the 10-11am hour.

The dwell time correction factor 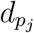.

To estimate the mean occupancy at each POI *p_j_* in an hour *t*, we multiply the expected number of visitors present at *p_j_* in hour *t* by the dwell time correction factor 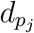, which measures the expected fraction of an hour that a visitor present at *p_j_* at any hour will spend there. In other words, conditioned on a visitor being at *p_j_* at some time within an hour *t*, *d_pj_*, is the expected fraction of the hour *t* that the visitor physically spends at *p_j_*. The same two assumptions above allow us to calculate *d_pj_*,: since each visitor stays for exactly 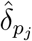, hours, and on average is counted as being present in 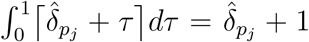 different hours, we have 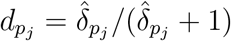.

##### Truncating outliers

As described in Methods M3.1, our model necessarily makes parametric assumptions about the relationship between POI characteristics (area, hourly visitors, and dwell time) and transmission rate at the POI; these assumptions may fail to hold for POIs which are outliers, particularly if SafeGraph data has errors. We mitigate this concern by truncating extreme values for POI characteristics to prevent data errors from unduly influencing our conclusions. Specifically, we truncate each POI’s area (i.e., square footage) to the 5th and 95th percentile of areas in the POI’s category; for every hour, we truncate the number of visitor arrivals for each POI to its category’s 95th percentile of visitor arrivals in that hour; and we truncate each POI’s median dwell time to its category’s 90th percentile of median dwell times in that period.

#### M7.2 Estimating the visit matrix *W*^(^*^t^*^)^

##### Overview

We estimate the visit matrix 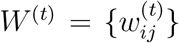, which captures the number of visitors from CBG *c_i_* to POI *p_j_* at each hour t from March 1, 2020 to May 2, 2020, through the iterative proportional fitting procedure (IPFP).^34^ The idea is as follows:

1. From SafeGraph data, we can derive a time-independent estimate 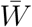 of the visit matrix that captures the aggregate distribution of visits from CBGs to POIs from January 2019 to February 2020.
2. However, visit patterns differ substantially from hour to hour (e.g., day versus night) and day to day (e.g., pre-versus post-lockdown). To capture these variations, we use current SafeGraph data to estimate the CBG marginals *U*^(^*^t^*^)^, i.e., the total number of visitors leaving each CBG at each time t, as well as the POI marginals *V*^(^*^t^*^)^, i.e., the total number of visitors present at each POI *p_j_* at time *t*.
3. We then use IPFP to estimate an hourly visit matrix *W*^(^*^t^*^)^ that is consistent with the hourly marginals *U*^(^*^t^*^)^ and *V*^(^*^t^*^)^ but otherwise “as similar as possible” to the distribution of visits in the aggregate visit matrix 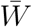. Here, similarity is defined in terms of Kullback-Leibler divergence; we provide a precise definition below.

##### Estimating the aggregate visit matrix 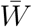

The estimated monthly visit matrices *Ŵ*^(^*^r^*^)^ are typically noisy and sparse: SafeGraph only matches a subset of visitors to POIs to their home CBGs, either for privacy reasons (if there are too few visitors from the given CBG) or because they are unable to link the visitor to a home CBG.^68^ To mitigate this issue, we aggregate these visit matrices, which are available at the monthly level, over the *R* =14 months from January 2019 to February 2020:

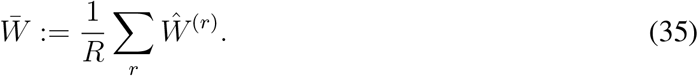

Each entry 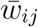 of 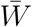 represents the estimated number of visitors from CBG *c_i_* that are present at POI *p_j_* in an hour, averaged over each hour. After March 2020, SafeGraph reports the visit matrices *Ŵ*^(^*^r^*^)^ on a weekly level in the Weekly Patterns v1 dataset. However, due to inconsistencies in the way SafeGraph processes the weekly vs. monthly matrices, we only use the monthly matrices up until February 2020.

##### Estimating the POI marginals *V*^(^*^t^*^)^

We estimate the POI marginals *V*^(^*^t^*^)^ ∊ ℝ*^n^*, whose *j*-th element 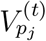 represents our estimate of the number of visitors at POI *p_j_* (from any CBG) at time *t*. The number of visitors recorded at POI *p_j_* at hour *t* in the SafeGraph data, 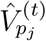 is an underestimate because the SafeGraph data only covers on a fraction of the overall population. To correct for this, we follow Benzell et al.^30^ and compute our final estimate of the visitors at POI *p_j_* in time *t* as

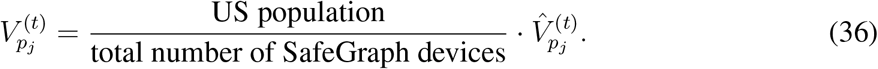

This correction factor is approximately 7, using population data from the most recent 1-year ACS (2018).

##### Estimating the CBG marginals *U*^(^*^t^*^)^

Next, we estimate the CBG marginals *U*^(^*^t^*^)^ ∊ ℝ*^m^*. Here, the i-th element 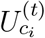 represents our estimate of the number of visitors leaving CBG *c_i_* (to visit any POI) at time *t*. We will also use *N_ci_*; recall that *N_ci_*. is the total population of *c_i_*, which is independent of *t*.

We first use the POI marginals *V*^(^*^t^*^)^ to calculate the total number of people who are out visiting any POI from any CBG at time *t*,

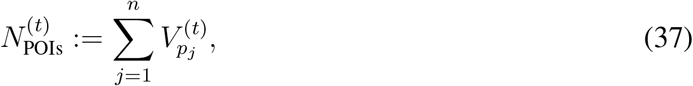

where *n* is the total number of POIs. Since the total number of people leaving any CBG to visit a POI must equal the total number of people at all the POIs, we have that 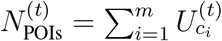, where *m* is the total number of CBGs.

Next, we estimate the number of people from each CBG *c_i_* who are not at home at time *t* as 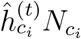. In general, the total number of people who are not at home in their CBGs, 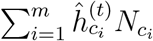, will not be equal to 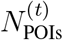, the number of people who are out visiting any POI. This discrepancy occurs for several reasons: for example, some people might have left their homes to travel to places that SafeGraph does not track, SafeGraph might not have been able to determine the home CBG of a POI visitor, etc.

To correct for this discrepancy, we assume that the relative proportions of POI visitors coming from each CBG follows the relative proportions of people who are not at home in each CBG. We thus estimate 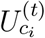 by apportioning the 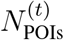 total POI visitors at time *t* according to the proportion of people who are not at home in each CBG *c_i_* at time *t*:

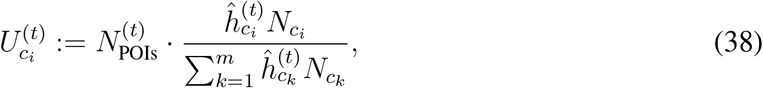

where *N_ci_* is the total population of CBG *i*, as derived from US Census data. This construction ensures that the POI and CBG marginals match, i.e., 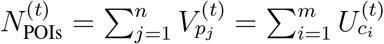.

##### Iterative proportional fitting procedure (IPFP)

IPFP is a classic statistical method^34^ for adjusting joint distributions to match pre-specified marginal distributions, and it is also known in the literature as biproportional fitting, the RAS algorithm, or raking.^69^ In the social sciences, it has been widely used to infer the characteristics of local subpopulations (e.g., within each CBG) from aggregate data.^70–72^

We estimate the visit matrix *W*^(^*^t^*^)^ by running IPFP on the aggregate visit matrix 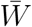, the CBG marginals *U*^(^*^t^*^)^, and the POI marginals *V*^(^*^t^*^)^ constructed above. Our goal is to construct a non-negative matrix *W*^(^*^t^*^)^ ∊ ℝ*^m^*^×^*^n^* whose rows sum up to the CBG marginals *U*^(^*^t^*^)^,

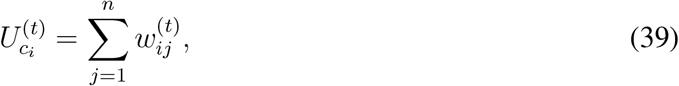

and whose columns sum up to the POI marginals 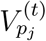,

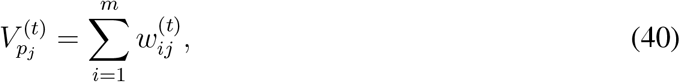

but whose distribution is otherwise “as similar as possible”, in the sense of Kullback-Leibler divergence, to the distribution over visits induced by the aggregate visit matrix 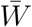.

###### Algorithm 1

Iterative proportional fitting procedure to estimate visit matrix *W*(*t*)

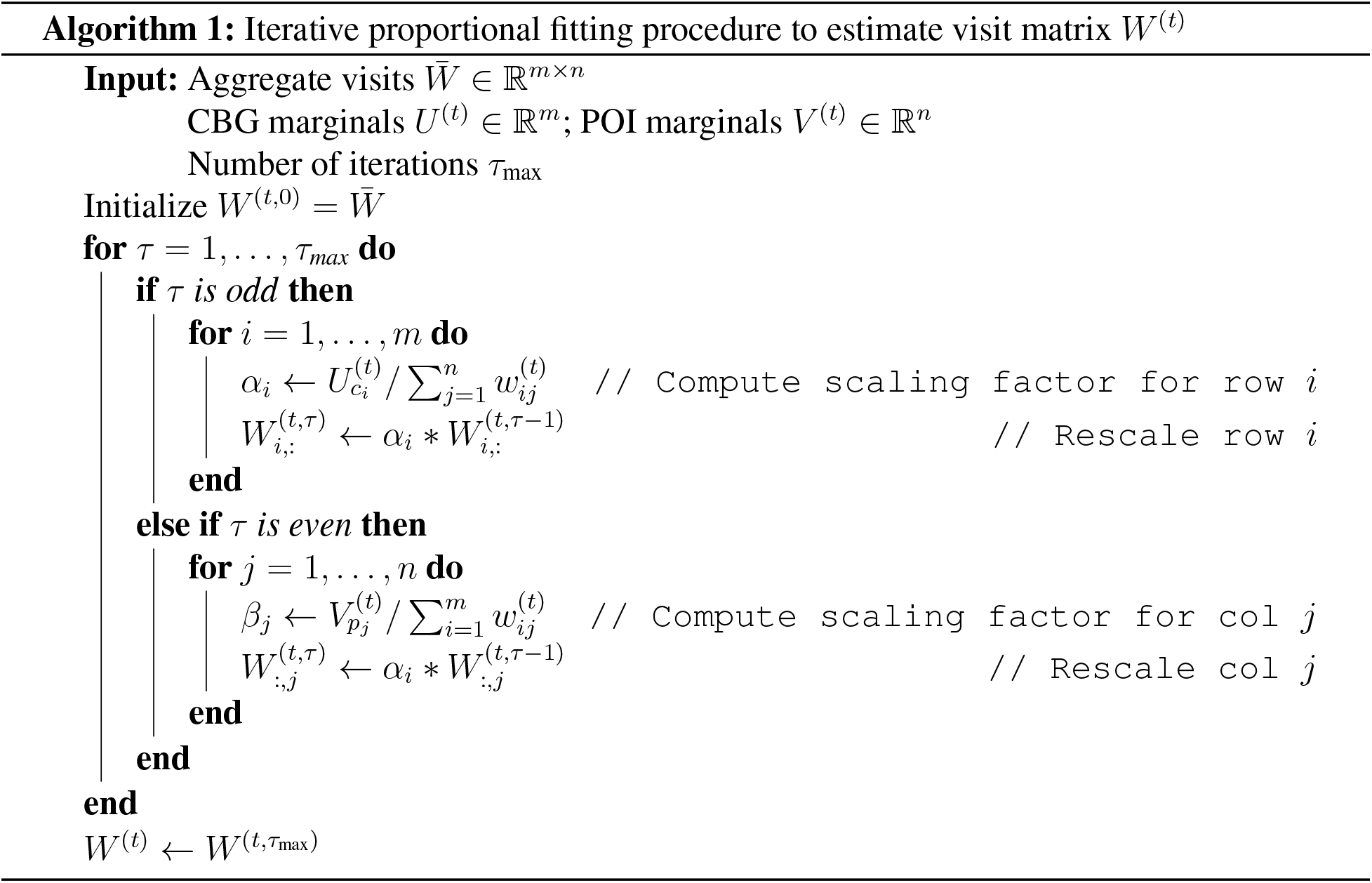

IPFP is an iterative algorithm that alternates between scaling each row to match the row (CBG) marginals *U*^(^*^t^*^)^ and scaling each column to match the column (POI) marginals *V*^(^*^t^*^)^. We provide pseudocode in Algorithm 1. For each value of t used in our simulation, we run IPFP separately for *τ*_max_ = 100 iterations. Note that IPFP is invariant to scaling the absolute magnitude of the entries in 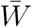, since the total number of visits it returns is fixed by the sum of the marginals; instead, its output depends only on the distribution over visits in 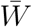.

The notion of similarity invoked above has a maximum likelihood interpretation: if IPFP converges, then it returns a visit matrix W^(t)^ whose induced distribution minimizes the Kullback-Leibler divergence to the distribution induced by 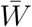.^73^ We further discuss the convergence of IPFP in our setting in SI Section S3.

## Data Availability

Census data, case and death counts from *The New York Times*, and Google mobility data are publicly available. Cell phone mobility data is freely available to researchers, non-profits, and governments through the SafeGraph COVID-19 Data Consortium.

## Code Availability

Code is in preparation and will be made publicly available at http://snap.stanford.edu/covid-mobility/.

## Acknowledgements

The authors thank Yong-Yeol Ahn, Nic Fishman, Tatsunori Hashimoto, Roni Rosenfeld, Jacob Steinhardt, Ryan Tibshirani, and our anonymous reviewers for helpful comments. We also thank Nick Singh, Ryan Fox Squire, Jessica Williams-Holt, Jonathan Wolf, Ruowei Yang, and others at SafeGraph for cell phone mobility data and helpful feedback. This research was supported by US National Science Foundation under OAC-1835598 (CINES), OAC-1934578 (HDR), CCF-1918940 (Expeditions), Chan Zuckerberg Biohub, Stanford Data Science Initiative, and the Stanford University Dean’s Research Fund. S.C. was supported by an NSF Fellowship. E.P. was supported by a Hertz Fellowship. P.W.K. was supported by the Facebook Fellowship Program. J.L. is a Chan Zuckerberg Biohub investigator.

## Author Contributions

S.C., E.P., and P.W.K. performed computational analysis. All authors jointly analyzed the results and wrote the paper.

## Author Information

The authors declare no conflict of interest. Correspondence should be addressed to.

## Extended data

**Extended Data Figure 1:**
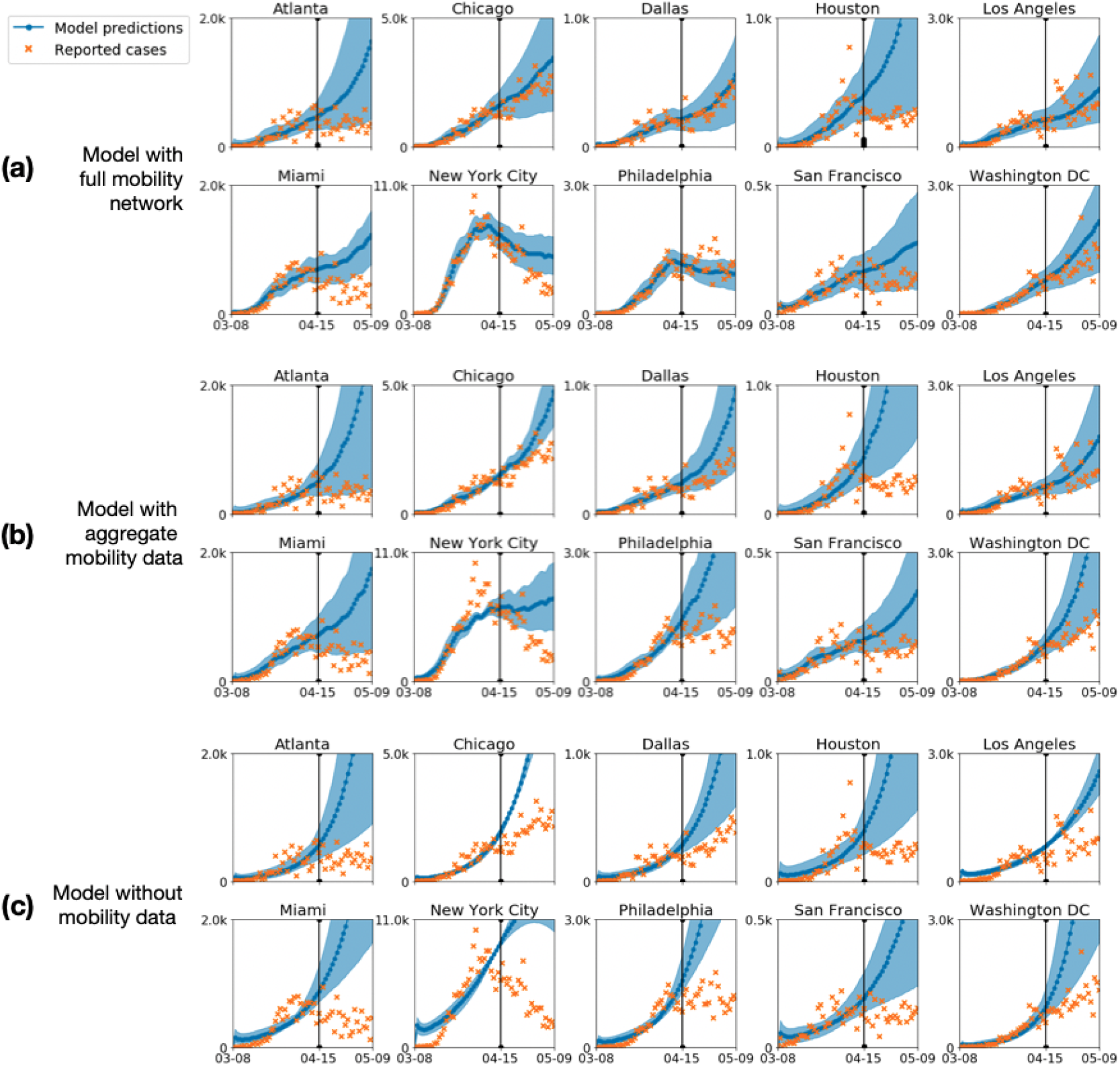
Predicted (blue) and true (orange) daily case counts for (a) our model, which uses hourly mobility networks, (b) an SEIR model which uses hourly aggregated mobility data, and (c) a baseline SEIR model which does not use mobility data. Incorporating mobility information improves out-of-sample fit and having a network, instead of an aggregate measure, further improves fit. All three models are calibrated on observed case counts before April 15, 2020 (vertical black line). Shaded regions denote 2.5th and 97.5th percentiles across sampled parameters and stochastic realizations. See Methods M4.4 and Methods M5.1 for details.

**Extended Data Figure 2:**
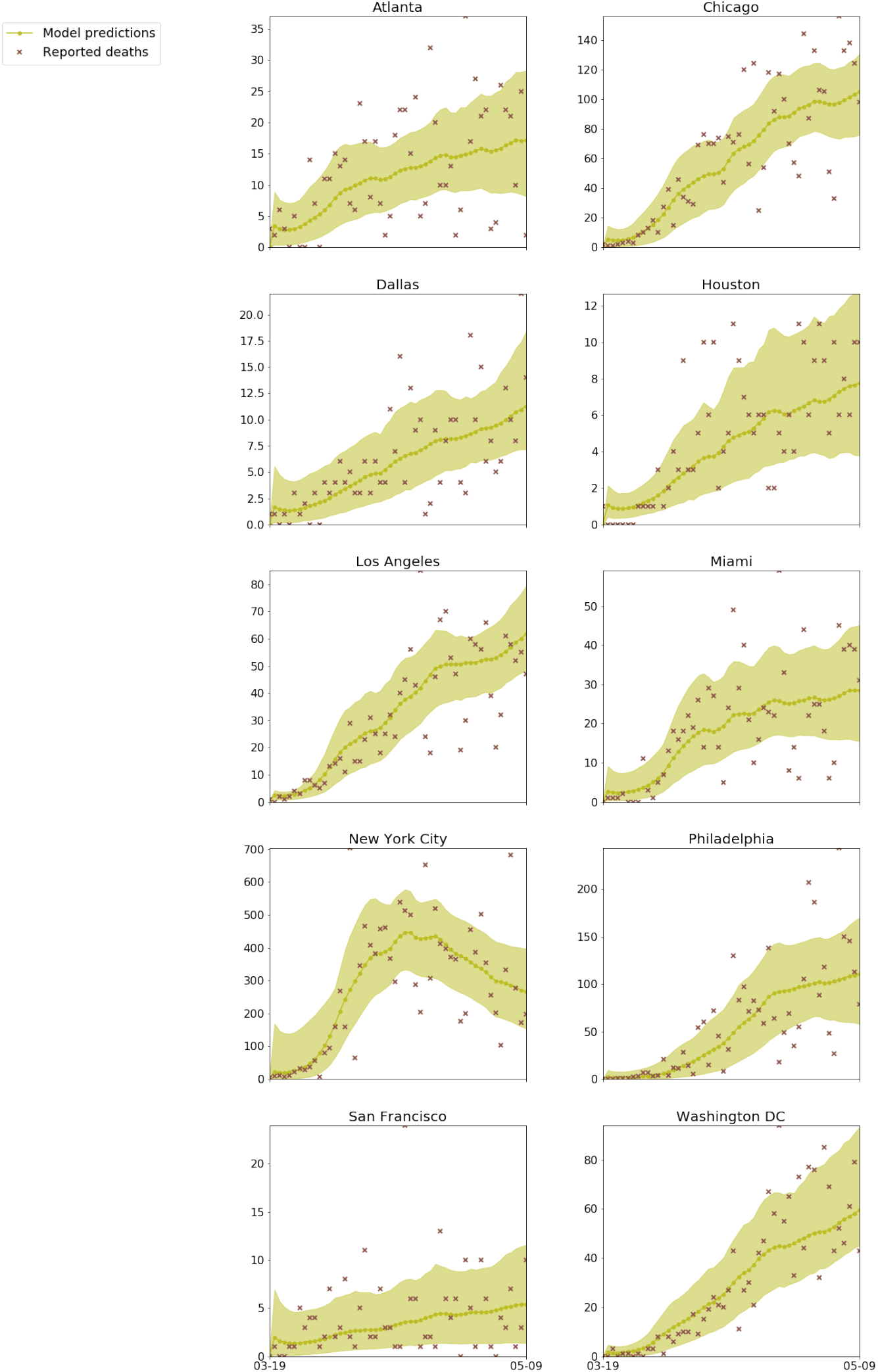
Predicted (green) and true (brown) daily death counts, when our model is calibrated on observed *death* counts from March 19 to May 9, 2020. Shaded regions denote 2.5th and 97.5th percentiles across sampled parameters and stochastic realizations. See Methods M5.5 for details.

**Extended Data Figure 3:**
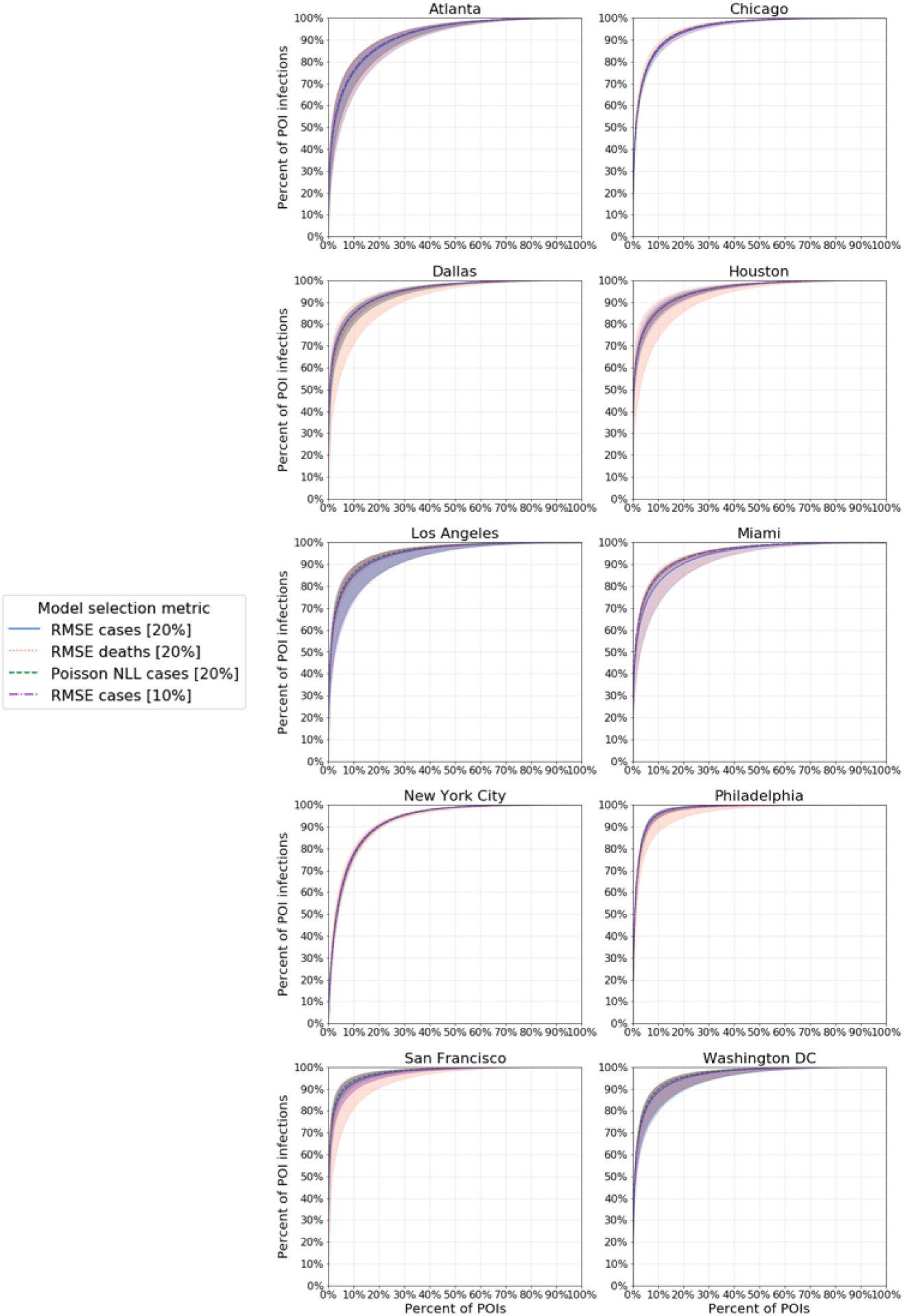
A small fraction of POIs account for a large fraction of the predicted infections at POIs. We additionally conducted a sensitivity analysis on which metric was used for model calibration and show that this key finding holds across all metrics. For each metric setting, we ran our models on the observed mobility data from March 1–May 2, 2020 and recorded the predicted number of infections that occurred at each POI. Shaded regions denote 2.5th and 97.5th percentiles across sampled parameters and stochastic realizations. See Methods M5.5 for details on model calibration metrics, and Methods M6 for details on this experiment.

**Extended Data Figure 4:**
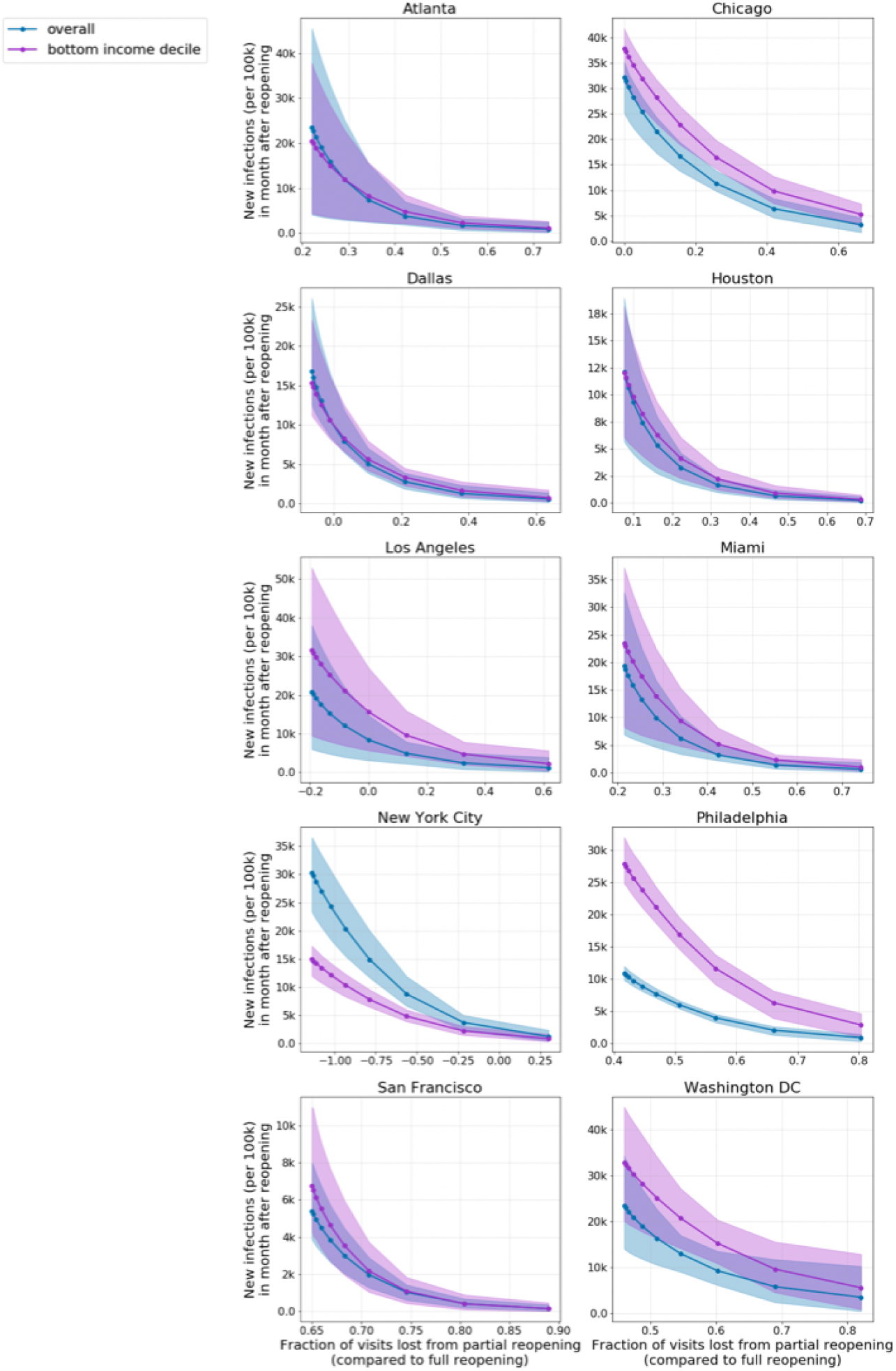
The predicted increase in infections under the reduced occupancy reopening strategy. We simulate reopening starting on May 1, 2020 and run the simulation until the end of the month. Each dot represents the level of occupancy reduction: e.g., capping visits at 50% of maximum occupancy, at 20% of maximum occupancy, etc. The y-coordinate of each dot represents the predicted number of new infections incurred after reopening (per 100k population) and its x-coordinate represents the fraction of visits lost from partial reopening compared to full reopening. Shaded regions denote 2.5th and 97.5th percentiles across sampled parameters and stochastic realizations. In 4 MSAs, the cost of new infections from reopening is roughly similar for lower-income CBGs and the overall population, but in 5 MSAs, the lower-income CBGs incur more infections from reopening. Notably, New York City (NYC) is the only MSA where this trend is reversed; this is because such a high fraction—65% (95% CI, 62%-68%)—of lower-income CBGs in NYC had been infected before reopening that after reopening, only a minority of the lower-income population is still susceptible (in comparison, the second highest fraction infected before reopening was 31% (95% CI, 28%-35%) for Philadelphia, and the rest ranged from 1%-14%). See Methods M6 for reopening details.

**Extended Data Figure 5:**
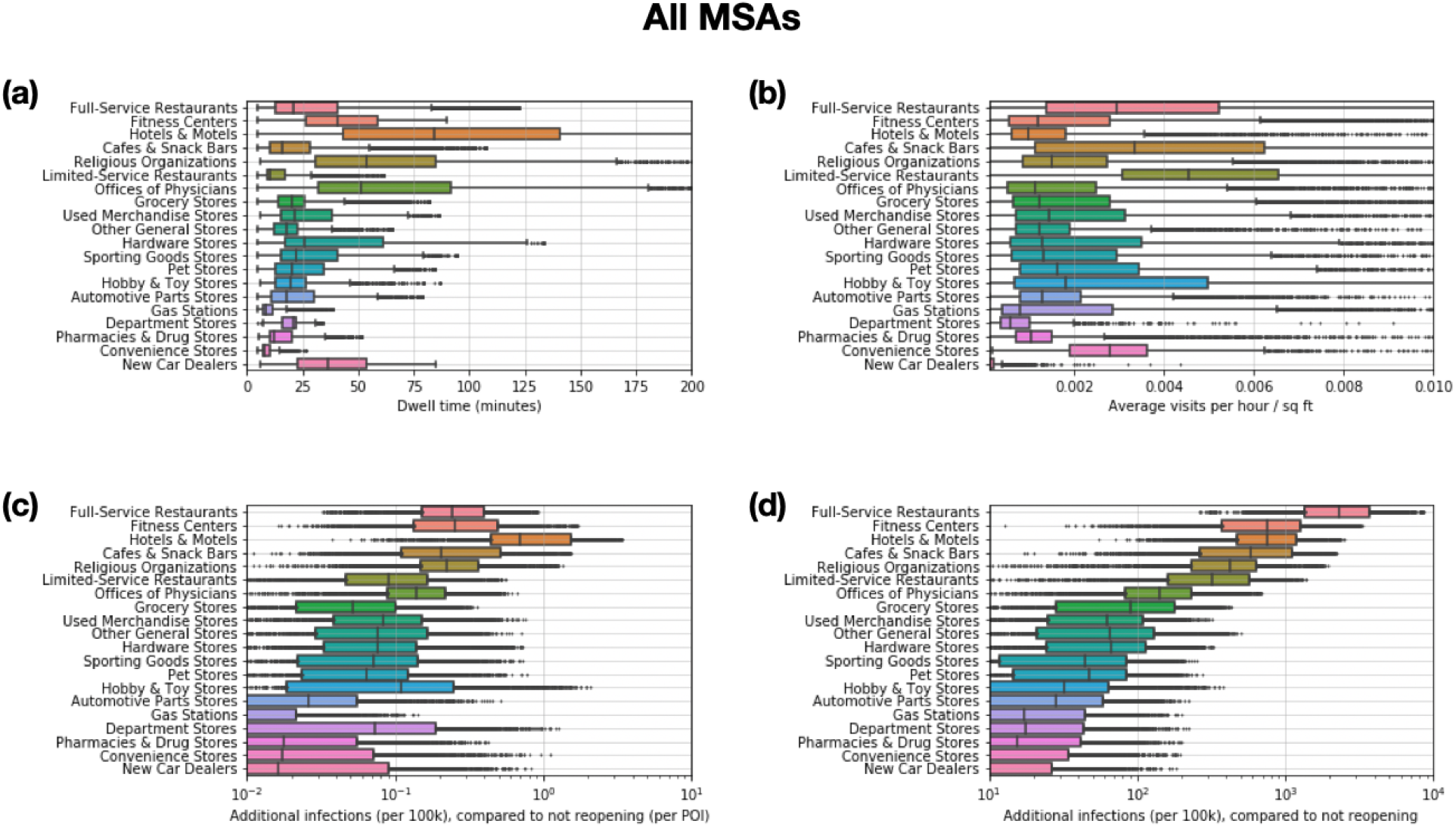
POI attributes in all 10 MSAs combined. The top two plots pool POIs from all MSAs, showing (**a**) the distribution of dwell time, and (**b**) the average number of hourly visitors divided by the area of the POI in square feet. Each point represents one POI; boxes depict the interquartile range across POIs. The bottom two plots pool across models from all MSAs, and show predictions for the increase in infections (per 100k population) from reopening a POI category: (**c**) per POI, and (**d**) for the category as a whole. Each point represents one model realization; boxes depict the interquartile range across sampled parameters and stochastic realizations. See Methods M6 for reopening details.

**Extended Data Figure 6:**
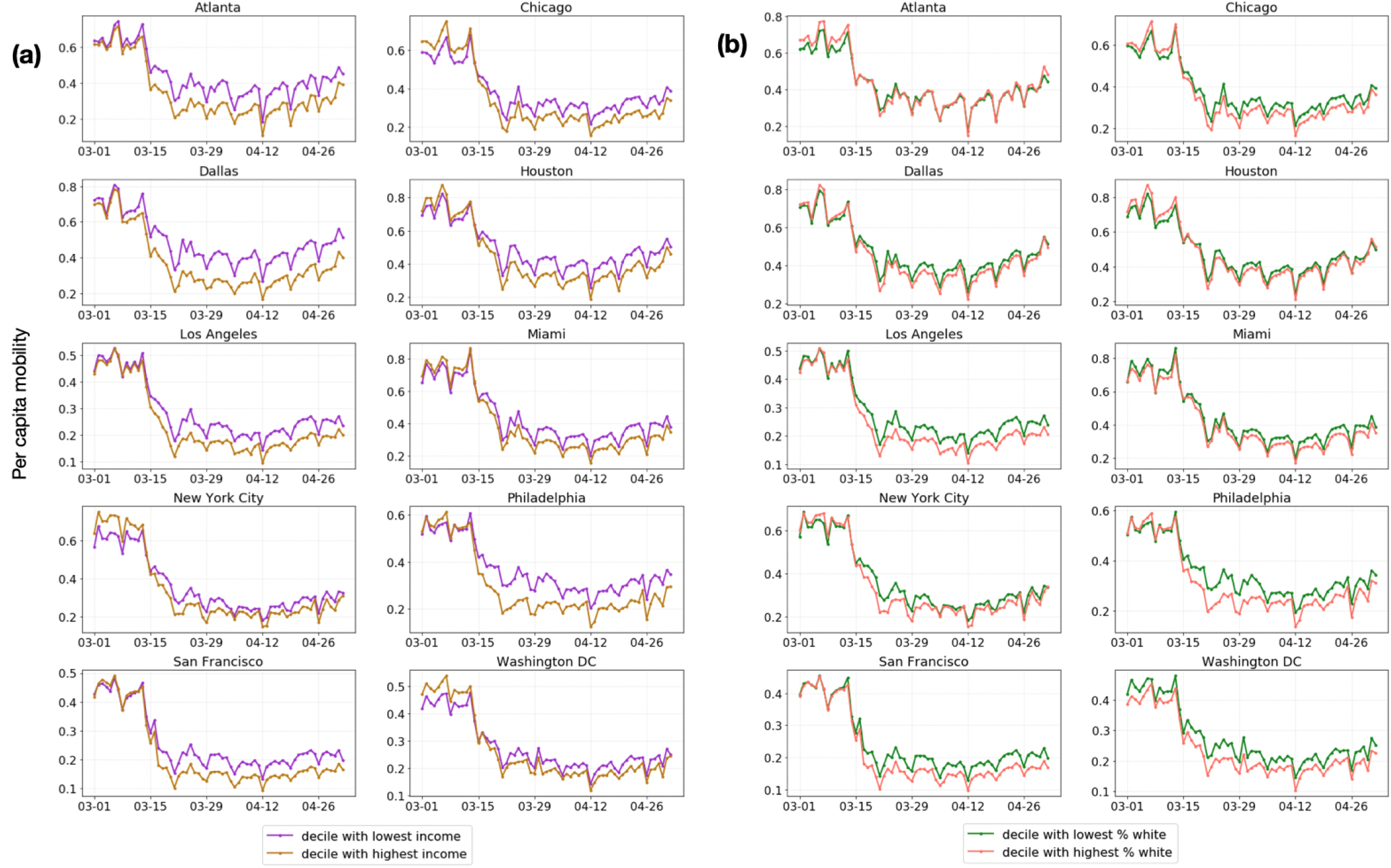
Daily per-capita mobility over time, (a) comparing lower-income to higher-income CBGs and (b) comparing less white to more white CBGs. See Methods M6 for details.

**Extended Data Figure 7:**
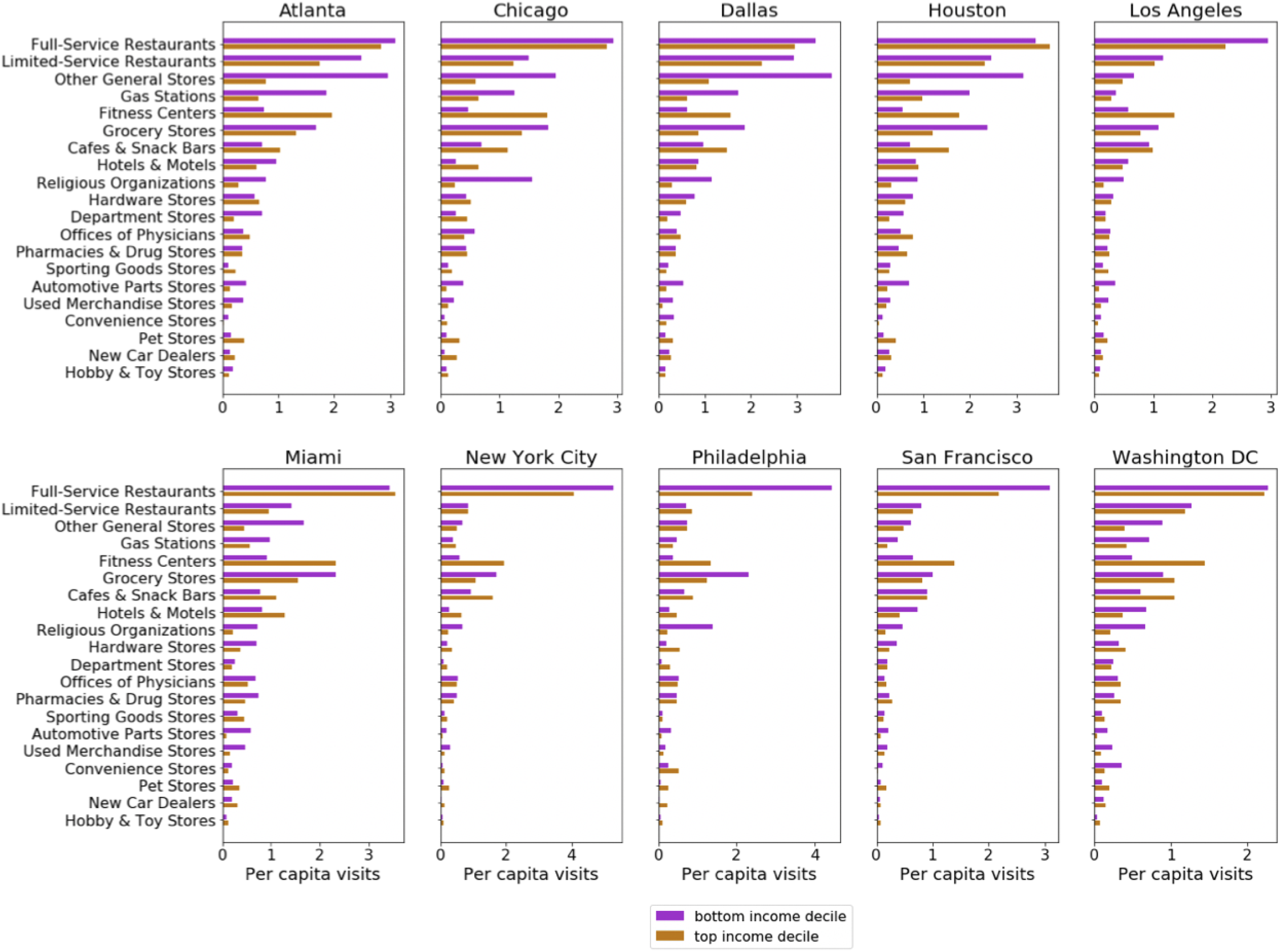
Visits per capita from CBGs in the bottom-(purple) and top-(gold) income deciles to each POI category, accumulated from March 1–May 2, 2020. See Methods M6 for details.

**Extended Data Figure 8:**
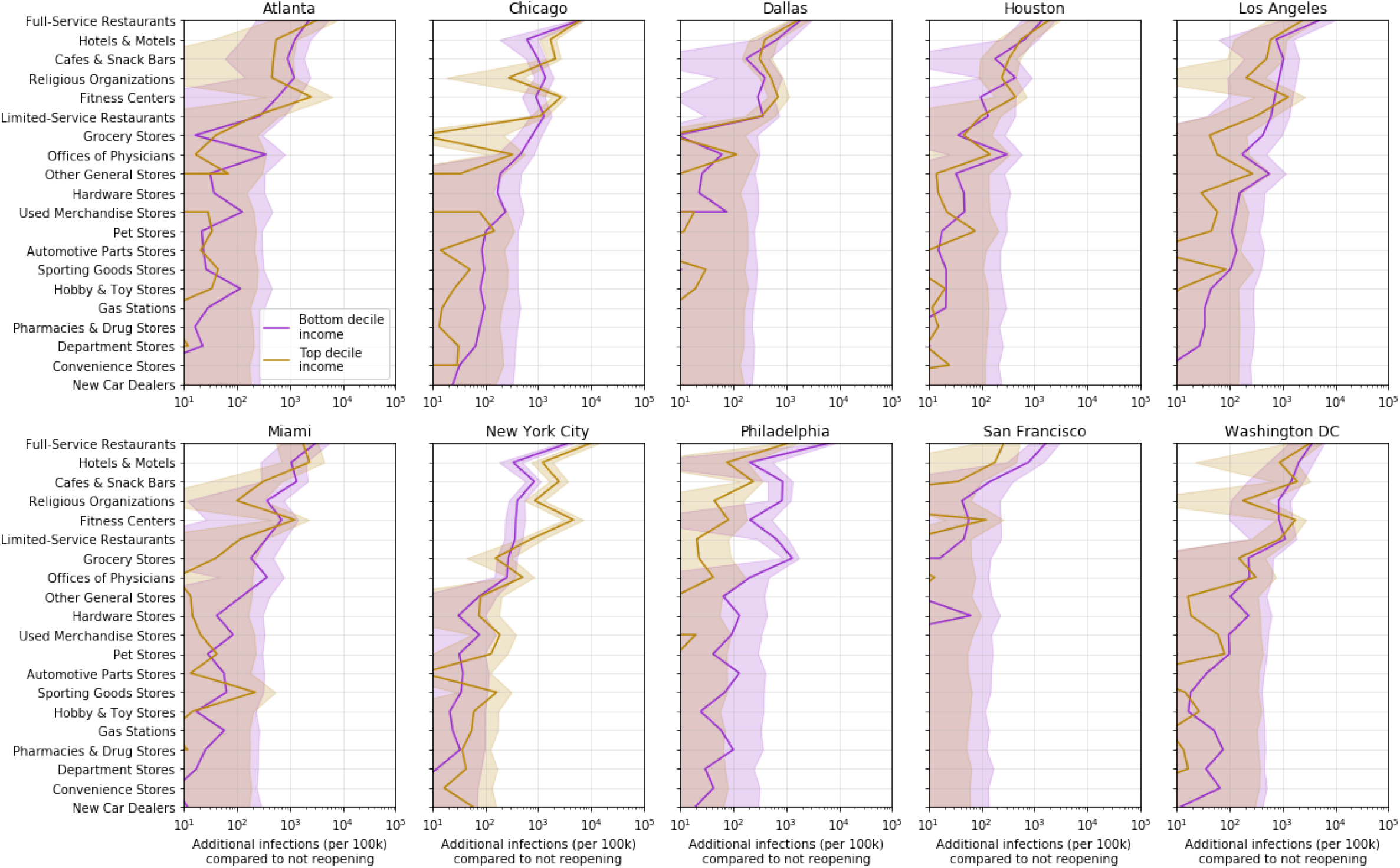
Predicted additional infections (per 100k population) from reopening each POI category, for CBGs in the top-(gold) and bottom-(purple) income deciles. Reopening impacts are generally worse for lower-income CBGs. See Methods M6 for reopening details.

**Extended Data Table 1:**
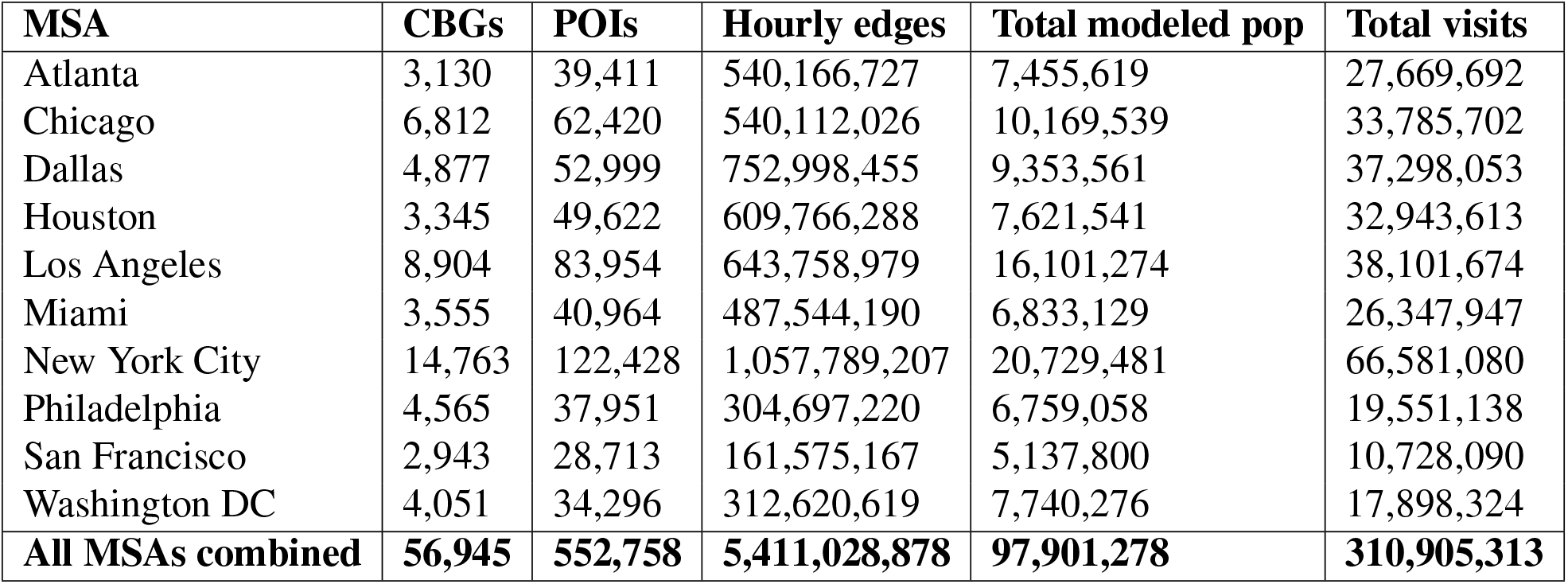
Dataset summary statistics from March 1–May 2,2020.

| **MSA** | **CBGs** | **POIs** | **Hourly edges** | **Total modeled pop** | **Total visits** |
| --- | --- | --- | --- | --- | --- |
| Atlanta | 3,130 | 39,411 | 540,166,727 | 7,455,619 | 27,669,692 |
| Chicago | 6,812 | 62,420 | 540,112,026 | 10,169,539 | 33,785,702 |
| Dallas | 4,877 | 52,999 | 752,998,455 | 9,353,561 | 37,298,053 |
| Houston | 3,345 | 49,622 | 609,766,288 | 7,621,541 | 32,943,613 |
| Los Angeles | 8,904 | 83,954 | 643,758,979 | 16,101,274 | 38,101,674 |
| Miami | 3,555 | 40,964 | 487,544,190 | 6,833,129 | 26,347,947 |
| New York City | 14,763 | 122,428 | 1,057,789,207 | 20,729,481 | 66,581,080 |
| Philadelphia | 4,565 | 37,951 | 304,697,220 | 6,759,058 | 19,551,138 |
| San Francisco | 2,943 | 28,713 | 161,575,167 | 5,137,800 | 10,728,090 |
| Washington DC | 4,051 | 34,296 | 312,620,619 | 7,740,276 | 17,898,324 |
| **All MSAs combined** | **56,945** | **552,758** | **5,411,028,878** | **97,901,278** | **310,905,313** |

**Extended Data Table 2:** Model parameters. If the parameter has a fixed value, we specify it under **Value**; otherwise, we write “Variable” to indicate that it varies across CBG / POI / MSA.

| Param. | Description | Value (Source) |
| --- | --- | --- |
| $\delta_E$ | mean latency period | 96 hours<sup>22,43</sup> |
| $\delta_I$ | mean infectious period | 84 hours<sup>22</sup> |
| $\delta_c$ | period from infectious to confirmed | 7 days<sup>22,74</sup> |
| $r_c$ | percentage of cases which are detected | 10%<sup>22,43,74–76</sup> |
| $\delta_d$ | period from infectious to death | 18 days<sup>66</sup> |
| $r_d$ | infection fatality rate | 0.66%<sup>66</sup> |
| $\beta_{\text{base}}$ | base CBG transmission rate | Variable (Estimated) |
| $N_{c_i}$ | population size of CBG $c_i$ | Variable (2018 US Census<sup>57</sup>) |
| $\psi$ | scaling factor for POI transmission | Variable (Estimated) |
| $w_{ij}^{(t)}$ | # visitors from CBG $c_i$ to POI $p_j$ at time $t$ | Variable (SafeGraph) |
| $a_{p_j}$ | area of POI $p_j$ in square feet | Variable (SafeGraph) |
| $p_0$ | initial proportion of latent population | Variable (Estimated) |
| $S_{c_i}^{(0)}$ | initial susceptible population in CBG $c_i$ | $(1 - p_0)N_{c_i}$ |
| $E_{c_i}^{(0)}$ | initial latent population in CBG $c_i$ | $p_0 N_{c_i}$ |
| $I_{c_i}^{(0)}$ | initial infected population in CBG $c_i$ | 0 |
| $R_{c_i}^{(0)}$ | initial removed population in CBG $c_i$ | 0 |

## Supplementary methods

### S1 Comparison of Google and SafeGraph mobility data

To assess the reliability of the SafeGraph datasets, we measured the correlation between mobility trends according to SafeGraph versus Google.^54^ Google provides a high-level picture of mobility changes around the world for several categories of places, such as grocery stores or restaurants. We analyzed three of the categories defined by Google: *Retail & recreation* (e.g., restaurants, shopping centers, movie theaters), *Grocery & pharmacy* (e.g., grocery stores, farmers markets, pharmacies), and *Residential* (i.e. places of residence). We omitted *Transit stations* because they are not well-covered by SafeGraph POIs, *Parks* because SafeGraph informed us that parks are sometimes inaccurately classified in their data (e.g., other POIs are categorized as parks), and *Workplaces* because we do not model whether people are at work. To analyze the *Retail & recreation* and *Grocery & pharmacy* categories, we used POI visits in the SafeGraph Patterns datasets, identifying POIs in each category based on their 6-digit North American Industry Classification System (NAICS) codes (Table S6). For the *Residential* category, we used SafeGraph Social Distancing Metrics, which provides daily counts of the number of people in each CBG who stayed at home for the entire day.

For each US region and category, Google tracks how the number of visits to the category has changed over the last few months, compared to baseline levels of activity before SARS-CoV-2. To set this baseline, they compute the median number of visits to the category for each day of the week, over a 5-week span from January 3–February 6, 2020. For a given day of interest, they then compute the relative change in number of visits seen on this day compared to the baseline for the corresponding day of week. We replicated this procedure on SafeGraph data, and compared the results to Google’s trends for Washington DC and 14 states that appear in the MSAs that we model. For each region and category, we measured the Pearson correlation between the relative change in number of visits according to Google versus Safegraph, from March 1–May 2, 2020. Across the 15 regions, we found that the median Pearson correlation was 0.96 for *Retail & recreation*, 0.79 for *Grocery & pharmacy*, and 0.88 for *Residential*. As an illustrative example, we visualize the results for New York state in Figure S3, and provide a full table of results for every state in Table S7. The high correlations demonstrate that the SafeGraph and Google mobility datasets agree well on the timing and directional changes of mobility over this time period, providing a validation of the reliability of SafeGraph data.

### S2 Plausibility of predicted racial/socioeconomic disparities

To assess the plausibility of the predicted disparities in infection rates in Figure 3, we compared the model’s predicted racial disparities to observed racial disparities in mortality rates. (Data on socioeconomic disparities in mortality was not systematically available on a national level.) The racial disparities in Figure 3 are generally of the same magnitude as reported racial disparities in mortality rates—for example, the overall reported black mortality rate is 2.4× higher than the white mortality rate,^77^ which is similar to the median racial disparity across MSAs of 3.0 × that our model predicts (Figure 3b). However, we note that this is an imperfect comparison because many factors besides mobility contribute to racial disparities in death rates.

In addition, we observed that our model predicted unusually large socioeconomic and racial disparities in infection rates in the Philadelphia MSA. To understand why the model predicted such large disparities, we inspected the mobility factors discussed in the main text; namely, how much each group was able to reduce their mobility, and whether disadvantaged groups encountered higher transmission rates at POIs.

First, we find in Philadelphia that higher-income CBGs were able to reduce their mobility substantially more than lower-income CBGs (Extended Data Figure 6 left). The CBGs with the greatest percentage of white residents were also able to reduce their mobility more than the CBGs with the lowest percentage of white residents (Extended Data Figure 6 right). These gaps are noticeable, but not obviously larger than those in other MSAs. The key to Philadelphia’s outlier status seems to lie in the comparison of transmission rates. Within the same category of POI—e.g., full-service restaurants—individuals from lower-income CBGs tend to visit POIs with higher transmission rates than individuals from high-income CBGs (Table S4). This is particularly true for Philadelphia; in 19 out of 20 categories, individuals from lower-income CBGs in Philadelphia encounter higher transmission rates than individuals from high-income CBGs, and CBGs with the lowest percentage of white residents encounter higher transmission rates than the CBGs with the highest percentage of white residents in all 20 categories (Table S5). The transmission rates encountered by individuals from lower-income CBGs in Philadelphia are often dramatically higher than those encountered by higher-income CBGs; for example, up to 10.4× higher for grocery stores. Digging deeper, this is because the average grocery store visited by lower-income CBGs has 5.3 × the number of hourly visitors per square foot, and visitors tend to stay 86% longer. Furthermore, Philadelphia’s large discrepancy in density between lower-income and higher-income POIs in SafeGraph data is consistent with Census data, which shows that the discrepancy in *population* density between lower-and higher-income CBGs is larger in Philadelphia than in any of the other MSAs that we examine. In Philadelphia, CBGs in the bottom income decile have a population density 8.2× those in the top income decile, a considerably larger disparity than the overall median across MSAs (3.3×) or the next-highest CBG (4.5×).

Since there are many other factors contributing to disparity that we do not model, we do not place too much weight on our model’s prediction that Philadelphia’s disparities will be larger than those of other cities. However, we consider this a valuable finding in terms of Philadelphia’s mobility patterns, suggesting that mobility may play an especially strong role in driving socioeconomic and racial infection disparities in this MSA, and we encourage policymakers to be aware of how differences in mobility patterns may exacerbate the disproportionate impact of SARS-CoV-2 on disadvantaged groups.

### S3 Convergence of iterative proportional fitting

For completeness, we briefly review the convergence properties of the iterative proportional fitting procedure (IPFP) used to infer our mobility networks. Consider the *L*_1_-error function

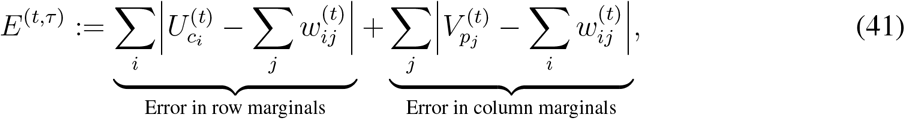

which sums up the errors in the row (CBG) and column (POI) marginals of the visit matrix *W*^(^*^t,τ^*^)^ from the *τ*-th iteration of IPFP. Each iteration of IPFP monotonically reduces this *L*_1_-error *E*^(^*^t,τ^*^)^, i.e., *E*^(^*^t,τ^*^)^ ≥ *E*^(^*^t,τ^*^+1)^ for all *τ* ≥ 0.^78^ In other words, the row and column sums of *W*^(^*^t,τ^*^)^ (which is initialized as 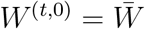) progressively get closer to (or technically, no further from) the target marginals as the iteration number *τ* increases. Moreover, IPFP maintains the cross-product ratios of the aggregate matrix 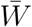, i.e.,

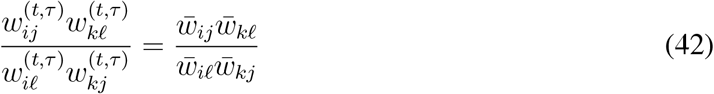

for all matrix entries indexed by *i*, *j*, *k*, *ℓ*, for all *t*, and for all iterations *τ*.

IPFP converges to a unique solution, in the sense that *W*^(^*^t^*^)^ = lim*_τ_*_→_*_∞_*∙*W*^(t,^*^τ^*^)^, if there exists a matrix *W*^(^*^t^*^)^ that fits the row and column marginals while maintaining the sparsity pattern (i.e., location of zeroes) of 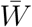.^78^ If IPFP converges, then the *L*_1_-error also converges to 0 as *τ*→*∞*,^78^ and *W*^(^*^t^*^)^ is the maximum likelihood solution in the following sense. For a visit matrix *W* = {*w_ij_*}, let *P_w_* represent a multinomial distribution over the *mn* entries of *W* with probability proportional to *W_j_*, and define 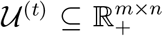 and 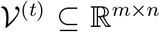 as the set of non-negative matrices whose row and column marginals match *U*^(^*^t^*^)^ and *V*^(^*^t^*^)^ respectively. Then, if IPFP converges,

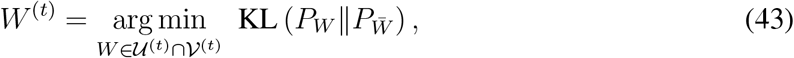

where KL (*p*||*q*) is the Kullback-Leibler divergence 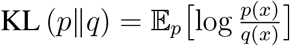. In other words, IPFP returns a visit matrix *W*^(^*^t^*^)^ whose induced distribution 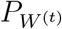 is the I-projection of the aggregate visit distribution 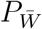 on the set of distributions with compatible row and column marginals.^73^ In fact, IPFP can be viewed as an alternating sequence of I-projections onto the row marginals and I-projections onto the column marginals.^73,79^

However, in our setting, IPFP typically does not return a unique solution and instead oscillates between two accumulation points, one that fits the row marginals and another that fits the column marginals.^79^ This is because 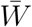 is highly sparse (there is no recorded interaction between most CBGs and POIs), so the marginals are sometimes impossible to reconcile. For example, suppose there is some CBG *c_i_* and POI *p_j_* such that 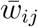 is the only non-zero entry in the *i*-th row and *j*-th column of 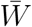, i.e., visitors from *c_i_* only travel to *p_j_* and conversely visitors from *p_j_* are all from *c_i_*. Then, if 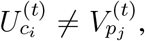 there does not exist any solution *W*^(^*^t^*^)^ such that 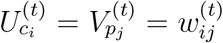. Note that in this scenario, IPFP still monotonically decreases the *L*_1_-error.^78^

In our implementation (Algorithm 1), we take *τ*_max_ = 100, so IPFP ends by fitting the column (POI) marginals. This ensures that our visit matrix *W*^(^*^t^*^)^ is fully compatible with the POI marginals *V*^(^*^t^*^)^, i.e.,

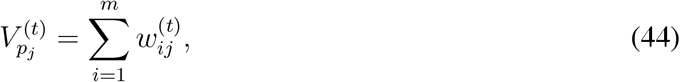

while still minimizing the *L*_1_-error *E*^(^*^t,τ^*^)^ with respect to the CBG marginals *U*^(^*^t^*^)^. Empirically, we find that *τ*_max_ = 100 iterations of IPFP are sufficient to converge to this oscillatory regime.

**Table S1:** The 50 POI subcategories accounting for the largest fraction of visits in the full SafeGraph dataset. Collectively they account for 88% of POI visits and 76% of POIs.

| Category | % visits | % POIs |
| --- | --- | --- |
| Full-Service Restaurants | 14.82% | 10.86% |
| Limited-Service Restaurants | 8.08% | 3.69% |
| Elementary and Secondary Schools | 6.36% | 3.06% |
| Other General Stores | 5.97% | 1.37% |
| Gas Stations | 4.56% | 2.94% |
| Fitness Centers | 4.55% | 2.98% |
| Grocery Stores | 4.16% | 2.17% |
| Cafes & Snack Bars | 4.01% | 2.70% |
| Hotels & Motels | 2.93% | 1.57% |
| Religious Organizations | 2.31% | 5.04% |
| Parks & Similar Institutions | 1.93% | 2.31% |
| Hardware Stores | 1.79% | 1.87% |
| Department Stores | 1.78% | 0.32% |
| Child Day Care Services | 1.71% | 2.76% |
| Offices of Physicians | 1.63% | 4.02% |
| Pharmacies & Drug Stores | 1.54% | 0.95% |
| Sporting Goods Stores | 1.16% | 1.05% |
| Automotive Parts Stores | 1.16% | 1.80% |
| Used Merchandise Stores | 1.15% | 1.01% |
| Colleges & Universities | 1.12% | 0.44% |
| Convenience Stores | 1.09% | 0.66% |
| Pet Stores | 0.93% | 0.85% |
| New Car Dealers | 0.73% | 0.43% |
| Hobby & Toy Stores | 0.73% | 0.36% |
| Offices of Dentists | 0.70% | 2.67% |
| Commercial Banking | 0.70% | 2.05% |
| Gift Stores | 0.69% | 0.57% |
| Liquor Stores | 0.61% | 0.82% |
| Women's Clothing Stores | 0.59% | 1.00% |
| Home Health Care Services | 0.55% | 1.02% |
| Furniture Stores | 0.53% | 0.89% |
| Electronics Stores | 0.51% | 0.72% |
| Used Car Dealers | 0.50% | 1.08% |
| Book Stores | 0.49% | 0.32% |
| Musical Instrument Stores | 0.49% | 0.50% |
| Optical Goods Stores | 0.47% | 0.76% |
| Family Clothing Stores | 0.46% | 0.49% |
| Car Repair Shops | 0.41% | 1.83% |
| Offices of Mental Health Practitioners | 0.41% | 1.05% |
| Tobacco Stores | 0.41% | 0.31% |
| Office Supplies | 0.40% | 0.33% |
| Beauty Salons | 0.39% | 1.58% |
| Paint and Wallpaper Stores | 0.38% | 0.56% |
| Other Gas Stations | 0.37% | 0.20% |
| Sports Teams and Clubs | 0.37% | 0.03% |
| Cosmetics & Beauty Stores | 0.36% | 0.71% |
| Jewelry Stores | 0.34% | 0.60% |
| Junior Colleges | 0.34% | 0.07% |
| Sewing & Piece Goods Stores | 0.34% | 0.39% |
| Senior Homes | 0.34% | 0.41% |
| Libraries & Archives | 0.3% | 0.3% |

**Table S2:** What if people had begun mobility reductions earlier or later? We report the expected ratio of the number of infections predicted under the counterfactual to the number of infections predicted using observed mobility data; a ratio lower than 1 means that fewer infections occurred under the counterfactual. The numbers in parentheses indicate the 2.5th and 97.5th percentiles across sampled parameters and stochastic realizations. See Methods M6 for details.

| MSA | 7 days earlier | 3 days earlier | 3 days later | 7 days later |
| --- | --- | --- | --- | --- |
| Atlanta | 0.586 (0.397, 0.834) | 0.803 (0.639, 0.956) | 1.359 (1.075, 1.741) | 1.981 (1.189, 2.761) |
| Chicago | 0.641 (0.563, 0.711) | 0.848 (0.769, 0.933) | 1.226 (1.143, 1.365) | 1.542 (1.446, 1.639) |
| Dallas | 0.642 (0.495, 0.782) | 0.855 (0.693, 1.013) | 1.298 (1.09, 1.577) | 1.722 (1.487, 1.966) |
| Houston | 0.656 (0.500, 0.812) | 0.848 (0.663, 1.021) | 1.288 (1.079, 1.541) | 1.731 (1.493, 2.064) |
| Los Angeles | 0.608 (0.407, 0.848) | 0.816 (0.639, 0.984) | 1.265 (1.041, 1.554) | 1.692 (1.216, 2.137) |
| Miami | 0.576 (0.424, 0.795) | 0.792 (0.669, 0.919) | 1.317 (1.117, 1.559) | 1.856 (1.281, 2.27) |
| New York City | 0.818 (0.795, 0.856) | 0.909 (0.890, 0.927) | 1.113 (1.094, 1.133) | 1.27 (1.246, 1.307) |
| Philadelphia | 0.799 (0.731, 0.868) | 0.916 (0.823, 1.005) | 1.12 (1.031, 1.206) | 1.287 (1.246, 1.351) |
| San Francisco | 0.609 (0.408, 0.798) | 0.815 (0.666, 1.012) | 1.271 (1.048, 1.527) | 1.689 (1.452, 2.029) |
| Washington DC | 0.671 (0.447, 0.879) | 0.848 (0.627, 1.045) | 1.207 (0.959, 1.586) | 1.488 (1.158, 1.789) |

**Table S3:**
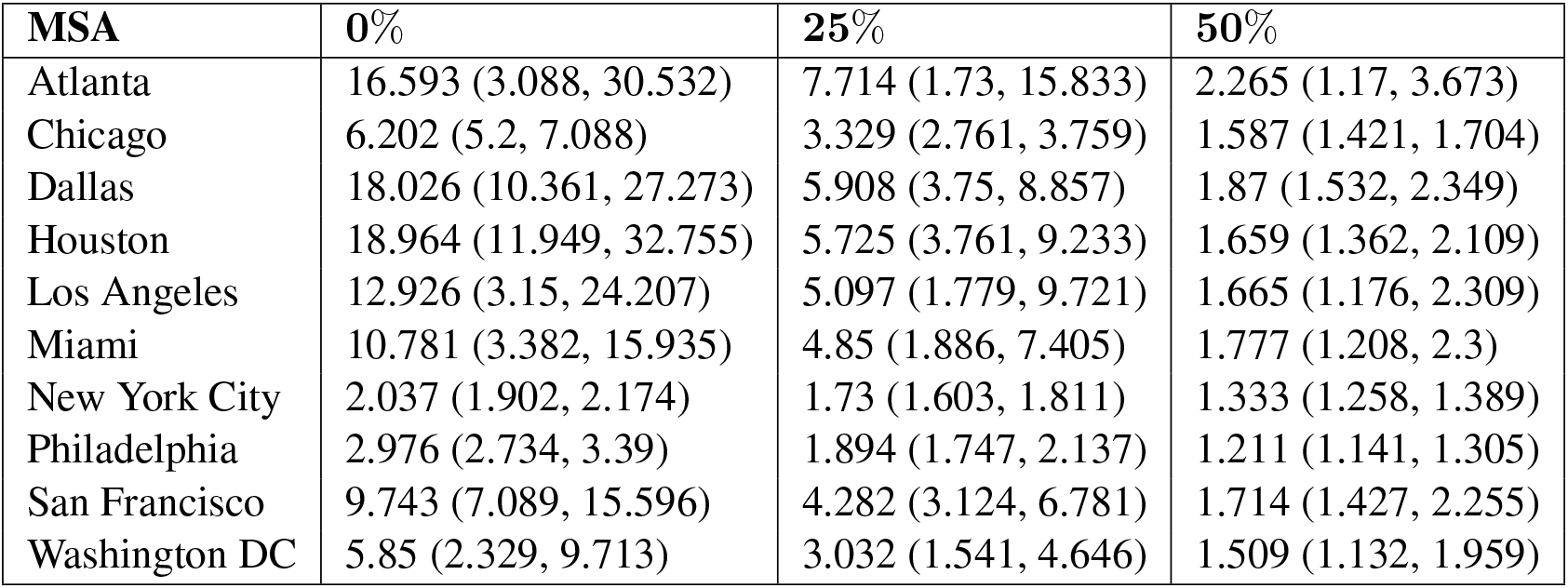
What if the magnitude of mobility reduction changed? Each column represents a counterfactual scenario where the magnitude of mobility reduction is only a some percentage of the observed mobility reduction, i.e., 0% corresponds to no mobility reduction, and 100% corresponds to the real, observed level of mobility reduction. We report the expected ratio of the number of infections predicted under the counterfactual to the number of infections predicted using observed mobility data; a ratio lower than 1 means that fewer infections occurred under the counterfactual. The numbers in parentheses indicate the 2.5th and 97.5th percentiles across sampled parameters and stochastic realizations. See Methods M6 for details.

**Table S4:** Transmission rate disparities at each POI category between income groups. We report the ratio of the average transmission rate encountered by visitors from CBGs in the bottom income decile to that for the top income decile. A ratio greater than 1 means that visitors from CBGs in the bottom income decile experienced higher (more dangerous) transmission rates. See Methods M6 for details.

|  | Atlanta | Chicago | Dallas | Houst. | LA | Miami | NY | Phila. | SF | DC | Median |
| --- | --- | --- | --- | --- | --- | --- | --- | --- | --- | --- | --- |
| Full-Service Restaurants | 0.764 | 1.204 | 0.956 | 1.000 | 1.445 | 1.232 | 2.035 | 2.883 | 1.758 | 1.171 | 1.218 |
| Limited-Service Restaurants | 0.940 | 0.950 | 1.002 | 0.906 | 1.067 | 0.872 | 1.901 | 1.614 | 0.994 | 0.962 | 0.978 |
| Other General Stores | 0.782 | 1.083 | 0.957 | 0.729 | 0.760 | 0.894 | 1.218 | 1.312 | 1.045 | 0.950 | 0.954 |
| Gas Stations | 1.326 | 1.865 | 1.310 | 1.515 | 2.254 | 2.195 | 1.899 | 6.461 | 1.357 | 1.870 | 1.868 |
| Fitness Centers | 0.536 | 0.907 | 0.708 | 0.670 | 1.461 | 0.789 | 1.151 | 1.516 | 0.995 | 1.160 | 0.951 |
| Grocery Stores | 0.948 | 3.080 | 0.838 | 1.333 | 2.408 | 1.498 | 4.984 | 10.437 | 2.478 | 1.977 | 2.192 |
| Cafes & Snack Bars | 1.385 | 0.919 | 0.716 | 1.120 | 1.327 | 2.168 | 1.943 | 1.757 | 0.982 | 0.932 | 1.224 |
| Hotels & Motels | 1.228 | 1.200 | 0.814 | 0.804 | 1.229 | 1.134 | 1.260 | 1.993 | 1.199 | 1.346 | 1.214 |
| Religious Organizations | 1.546 | 1.763 | 0.956 | 0.919 | 1.746 | 1.464 | 1.756 | 1.736 | 1.515 | 1.852 | 1.641 |
| Hardware Stores | 3.938 | 3.340 | 1.575 | 2.111 | 1.333 | 0.939 | 3.553 | 6.716 | 4.202 | 13.560 | 3.446 |
| Department Stores | 1.132 | 1.230 | 0.978 | 0.911 | 1.083 | 1.431 | 1.667 | 0.976 | 0.867 | 1.042 | 1.062 |
| Offices of Physicians | 1.235 | 0.721 | 0.667 | 1.036 | 1.141 | 1.687 | 1.307 | 1.319 | 1.193 | 0.445 | 1.167 |
| Pharmacies & Drug Stores | 1.636 | 1.389 | 1.176 | 0.854 | 1.718 | 1.555 | 2.577 | 5.624 | 1.200 | 1.699 | 1.596 |
| Sporting Goods Stores | 0.936 | 1.540 | 1.129 | 0.812 | 1.168 | 0.700 | 1.253 | 1.161 | 0.826 | 2.777 | 1.145 |
| Automotive Parts Stores | 0.890 | 1.707 | 0.862 | 1.086 | 1.990 | 1.414 | 1.524 | 2.697 | 1.753 | 1.246 | 1.469 |
| Used Merchandise Stores | 0.993 | 0.931 | 1.000 | 1.315 | 1.017 | 1.074 | 1.352 | 1.668 | 1.587 | 0.814 | 1.046 |
| Convenience Stores | 1.208 | 0.932 | 1.613 | 0.647 | 0.838 | 0.824 | 1.736 | 2.322 | 1.086 | 1.428 | 1.147 |
| Pet Stores | 1.260 | 0.820 | 1.192 | 1.487 | 1.536 | 0.776 | 3.558 | 1.652 | 2.124 | 0.905 | 1.374 |
| New Car Dealers | 2.036 | 1.471 | 0.741 | 0.809 | 1.180 | 1.377 | 2.022 | 1.129 | 0.395 | 0.872 | 1.154 |
| Hobby & Toy Stores | 1.168 | 1.110 | 1.165 | 0.853 | 1.771 | 1.520 | 1.525 | 1.088 | 0.883 | 0.926 | 1.138 |
| **Median** | 1.188 | 1.202 | 0.968 | 0.915 | 1.330 | 1.305 | 1.746 | 1.702 | 1.196 | 1.166 |  |

**Table S5:** Transmission rate disparities at each POI category between racial groups. We report the ratio of the average transmission rate encountered by visitors from CBGs with the lowest (bottom decile) proportion of white residents versus that for the top decile. A ratio greater than 1 means that visitors from CBGs in the bottom decile experienced higher (more dangerous) transmission rates. See Methods M6 for details.

|  | Atlanta | Chicago | Dallas | Houst. | LA | Miami | NY | Phila. | SF | DC | Median |
| --- | --- | --- | --- | --- | --- | --- | --- | --- | --- | --- | --- |
| Full-Service Restaurants | 0.802 | 1.354 | 0.981 | 0.965 | 1.065 | 1.167 | 2.418 | 2.661 | 1.223 | 1.013 | 1.116 |
| Limited-Service Restaurants | 0.940 | 1.144 | 1.028 | 0.940 | 0.820 | 0.919 | 2.136 | 1.523 | 0.799 | 1.346 | 0.984 |
| Other General Stores | 0.776 | 1.277 | 0.838 | 0.841 | 1.527 | 1.132 | 2.158 | 1.313 | 0.925 | 1.312 | 1.204 |
| Gas Stations | 1.402 | 1.891 | 1.389 | 1.190 | 1.336 | 1.857 | 1.818 | 2.286 | 2.321 | 1.316 | 1.610 |
| Fitness Centers | 0.607 | 1.167 | 0.670 | 0.831 | 0.780 | 1.066 | 1.447 | 1.977 | 1.103 | 1.205 | 1.084 |
| Grocery Stores | 0.589 | 3.664 | 0.613 | 1.195 | 2.386 | 0.950 | 5.864 | 13.705 | 2.243 | 2.262 | 2.252 |
| Cafes & Snack Bars | 1.308 | 1.104 | 0.845 | 0.840 | 0.976 | 2.619 | 1.767 | 2.456 | 1.045 | 0.867 | 1.074 |
| Hotels & Motels | 0.977 | 1.007 | 1.366 | 0.718 | 1.112 | 1.024 | 1.449 | 2.494 | 0.654 | 0.899 | 1.015 |
| Religious Organizations | 0.938 | 1.606 | 1.060 | 0.953 | 2.096 | 1.795 | 1.933 | 2.040 | 1.674 | 1.188 | 1.640 |
| Hardware Stores | 0.909 | 3.900 | 1.523 | 1.461 | 1.952 | 0.586 | 5.032 | 3.898 | 11.103 | 13.432 | 2.925 |
| Department Stores | 1.081 | 1.301 | 0.805 | 0.777 | 0.992 | 2.337 | 2.479 | 1.357 | 1.089 | 1.402 | 1.195 |
| Offices of Physicians | 0.894 | 1.323 | 1.006 | 1.415 | 0.898 | 1.117 | 1.652 | 2.073 | 0.694 | 1.911 | 1.220 |
| Pharmacies & Drug Stores | 0.888 | 1.376 | 0.930 | 0.732 | 1.538 | 1.674 | 3.315 | 3.366 | 1.135 | 1.715 | 1.457 |
| Sporting Goods Stores | 0.767 | 0.674 | 0.650 | 0.506 | 1.946 | 0.818 | 1.532 | 2.152 | 0.880 | 1.715 | 0.849 |
| Automotive Parts Stores | 1.049 | 1.479 | 1.010 | 1.353 | 2.998 | 2.657 | 1.740 | 3.387 | 1.646 | 0.601 | 1.562 |
| Used Merchandise Stores | 0.858 | 1.195 | 0.699 | 1.060 | 1.270 | 0.593 | 1.500 | 3.024 | 1.425 | 0.799 | 1.128 |
| Convenience Stores | 2.016 | 5.055 | 1.272 | 2.188 | 0.761 | 0.902 | 1.911 | 2.276 | 1.239 | 1.844 | 1.878 |
| Pet Stores | 0.925 | 1.624 | 0.724 | 1.465 | 1.506 | 0.881 | 2.715 | 10.182 | 1.568 | 2.408 | 1.537 |
| New Car Dealers | 1.008 | 1.398 | 0.812 | 0.736 | 0.942 | 0.998 | 1.977 | 0.866 | 0.772 | 0.383 | 0.904 |
| Hobby & Toy Stores | 2.569 | 0.853 | 0.628 | 0.979 | 1.373 | 1.388 | 2.237 | 0.825 | 0.864 | 1.286 | 1.132 |
| **Median** | 0.932 | 1.339 | 0.888 | 0.959 | 1.303 | 1.092 | 1.955 | 2.281 | 1.119 | 1.314 |  |

**Table S6:** Mapping of Google mobility data categories to NAICS categories. Google descriptions taken from https://www.google.com/covid19/mobility/data_documentation.html.

| Google category | Google description | NAICS categories |
| --- | --- | --- |
| Retail & recreation | Restaurants | Full-Service Restaurants |
|  | Cafes | Limited-Service Restaurants |
|  | Shopping centers | Snack and Nonalcoholic Beverage Bars |
|  | Theme parks | Drinking Places (Alcoholic Beverages) |
|  | Museums | Malls, Amusement and Theme Parks |
|  | Libraries | Museums, Libraries and Archives |
|  | Movie theaters | Motion Picture Theaters (except Drive-Ins) |
| Grocery & pharmacy | Grocery markets | Supermarkets and Other Grocery (except Convenience) Stores |
|  | Food warehouses | Food (Health) Supplement Stores |
|  | Farmers markets | Fish and Seafood Markets |
|  | Specialty food shops | All Other Specialty Food Stores |
|  | Drug stores | Pharmacies and Drug Stores |
|  | Pharmacies |  |

**Table S7:** Pearson correlations between the Google and SafeGraph mobility timeseries. We report correlations over the period of March 1–May 2, 2020 for the 15 states that we model. See SI Section S1 for details.

| State | Retail & recreation | Grocery & pharmacy | Residential |
| --- | --- | --- | --- |
| California | 0.947 | 0.834 | 0.876 |
| Delaware | 0.957 | 0.847 | 0.856 |
| Florida | 0.963 | 0.814 | 0.885 |
| Georgia | 0.948 | 0.682 | 0.868 |
| Illinois | 0.964 | 0.710 | 0.899 |
| Indiana | 0.956 | 0.741 | 0.877 |
| Maryland | 0.956 | 0.825 | 0.886 |
| New Jersey | 0.951 | 0.720 | 0.935 |
| New York | 0.958 | 0.763 | 0.909 |
| Pennsylvania | 0.971 | 0.850 | 0.875 |
| Texas | 0.965 | 0.789 | 0.886 |
| Virginia | 0.967 | 0.840 | 0.877 |
| Washington, DC | 0.959 | 0.889 | 0.780 |
| West Virginia | 0.960 | 0.740 | 0.814 |
| Wisconsin | 0.967 | 0.783 | 0.886 |
| **Median** | 0.959 | 0.789 | 0.877 |

**Table S8:** Model parameters used for each MSA. # sets counts the number of parameter sets that are within 20% of the RMSE of the best-fit parameter set, as described in Section M4. For each of *β*_base_ (which scales the transmission rates at CBGs), *ψ* (which scales the transmission rates at POIs), and *p*_0_ (the initial proportion of infected individuals), we show the best-fit parameter set and, in parentheses, the corresponding minimum and maximum within the 20% threshold.

| MSA | # sets | $\beta_{\text{base}}$ | $\psi$ | $p_0$ |
| --- | --- | --- | --- | --- |
| Atlanta | 16 | 0.004 (0.001, 0.014) | 2388 (515, 3325) | $5 \times 10^{-4} (1 \times 10^{-4}, 2 \times 10^{-3})$ |
| Chicago | 4 | 0.009 (0.006, 0.011) | 1764 (1139, 2076) | $2 \times 10^{-4} (2 \times 10^{-4}, 5 \times 10^{-4})$ |
| Dallas | 5 | 0.009 (0.004, 0.011) | 1452 (1139, 2388) | $2 \times 10^{-4} (1 \times 10^{-4}, 2 \times 10^{-4})$ |
| Houston | 8 | 0.001 (0.001, 0.009) | 2076 (1139, 2076) | $2 \times 10^{-4} (1 \times 10^{-4}, 5 \times 10^{-4})$ |
| Los Angeles | 25 | 0.006 (0.001, 0.016) | 2076 (515, 3637) | $2 \times 10^{-4} (2 \times 10^{-5}, 1 \times 10^{-3})$ |
| Miami | 7 | 0.001 (0.001, 0.011) | 2388 (515, 2388) | $2 \times 10^{-4} (2 \times 10^{-4}, 2 \times 10^{-3})$ |
| New York City | 7 | 0.001 (0.001, 0.009) | 2700 (1452, 3013) | $1 \times 10^{-4} (5 \times 10^{-5}, 1 \times 10^{-3})$ |
| Philadelphia | 3 | 0.009 (0.001, 0.009) | 827 (827, 1452) | $5 \times 10^{-4} (1 \times 10^{-4}, 5 \times 10^{-4})$ |
| San Francisco | 5 | 0.006 (0.001, 0.009) | 1139 (827, 1764) | $5 \times 10^{-4} (2 \times 10^{-4}, 1 \times 10^{-3})$ |
| Washington DC | 17 | 0.016 (0.001, 0.019) | 515 (515, 3949) | $5 \times 10^{-4} (2 \times 10^{-5}, 5 \times 10^{-4})$ |

**Figure S1:**
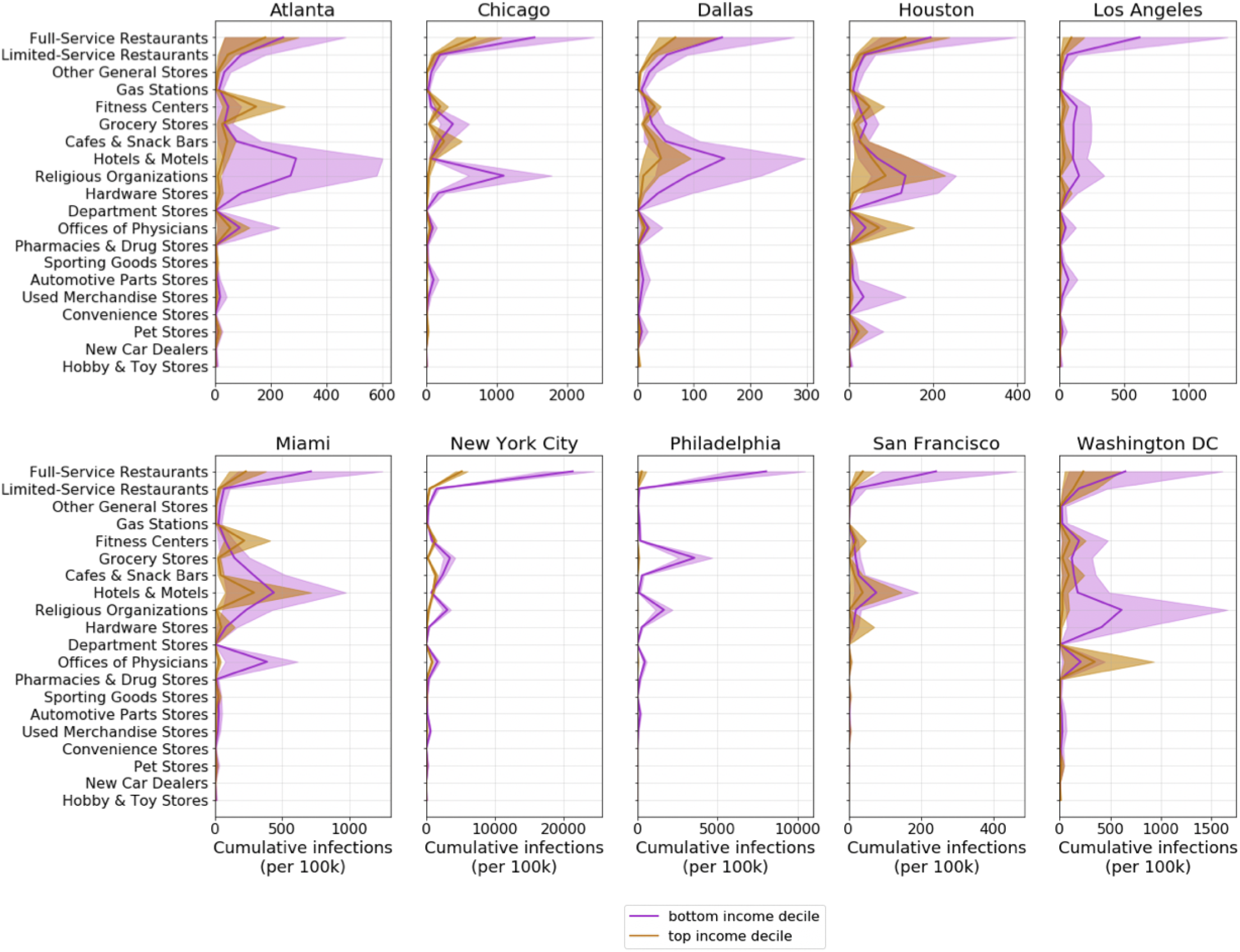
For each POI category, we plot the predicted cumulative number of infections (per 100k population) that occurred at that category for CBGs in the bottom-(purple) and top-(gold) income deciles. Shaded regions denote 2.5th and 97.5th percentiles across sampled parameters and stochastic realizations.

**Figure S2:**
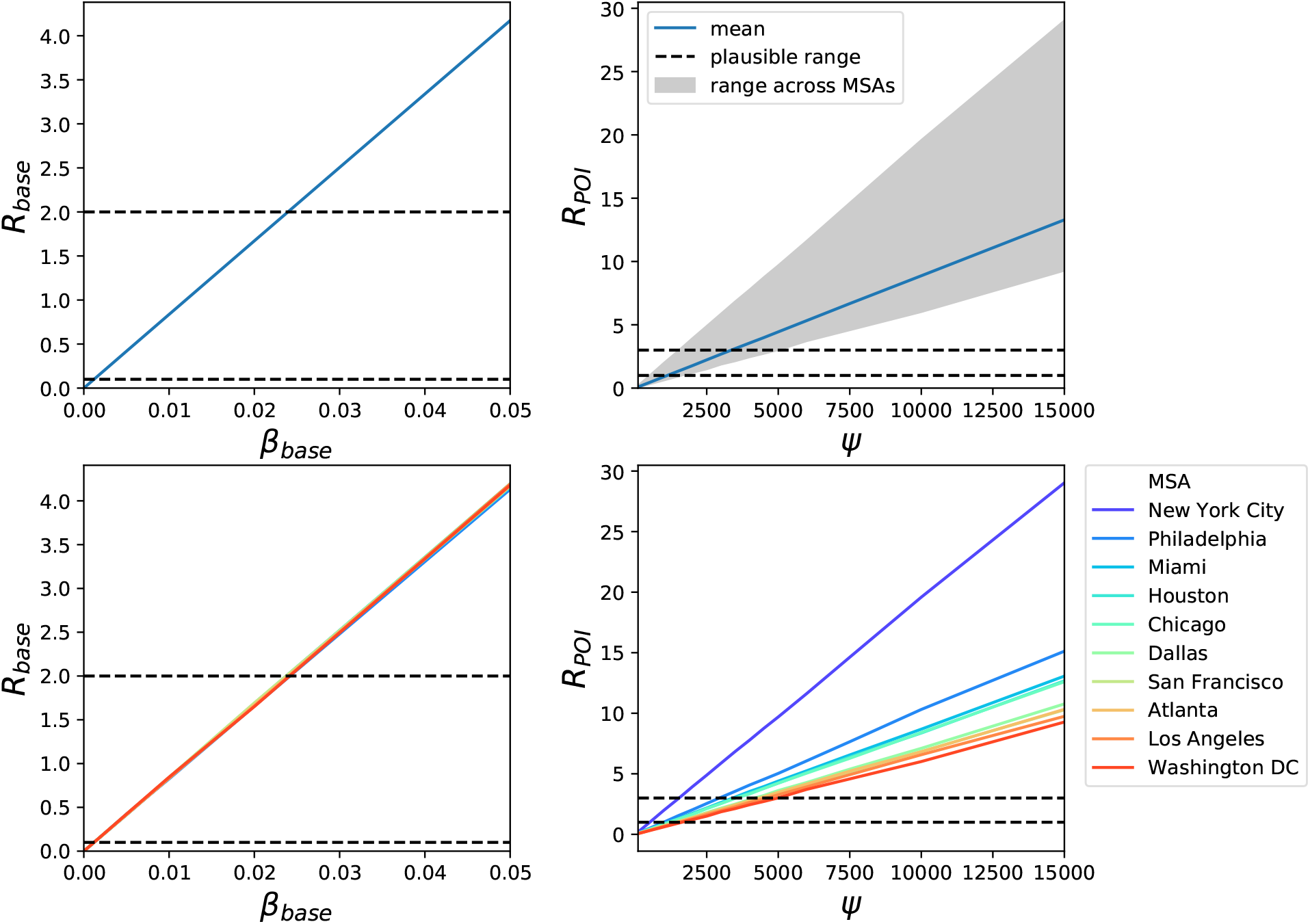
*R*_base_ and *R*_POI_ implied by model parameter settings. In the top two plots, dotted black lines denote plausible ranges from prior work, the blue line shows the mean across MSAs, and the grey shaded area indicates the range across MSAs. *R*_base_ does not vary across MSAs because it does not depend on MSA-specific social activity. The bottom two plots show the same results broken down by MSA. See Methods M4.1 for details.

**Figure S3:**
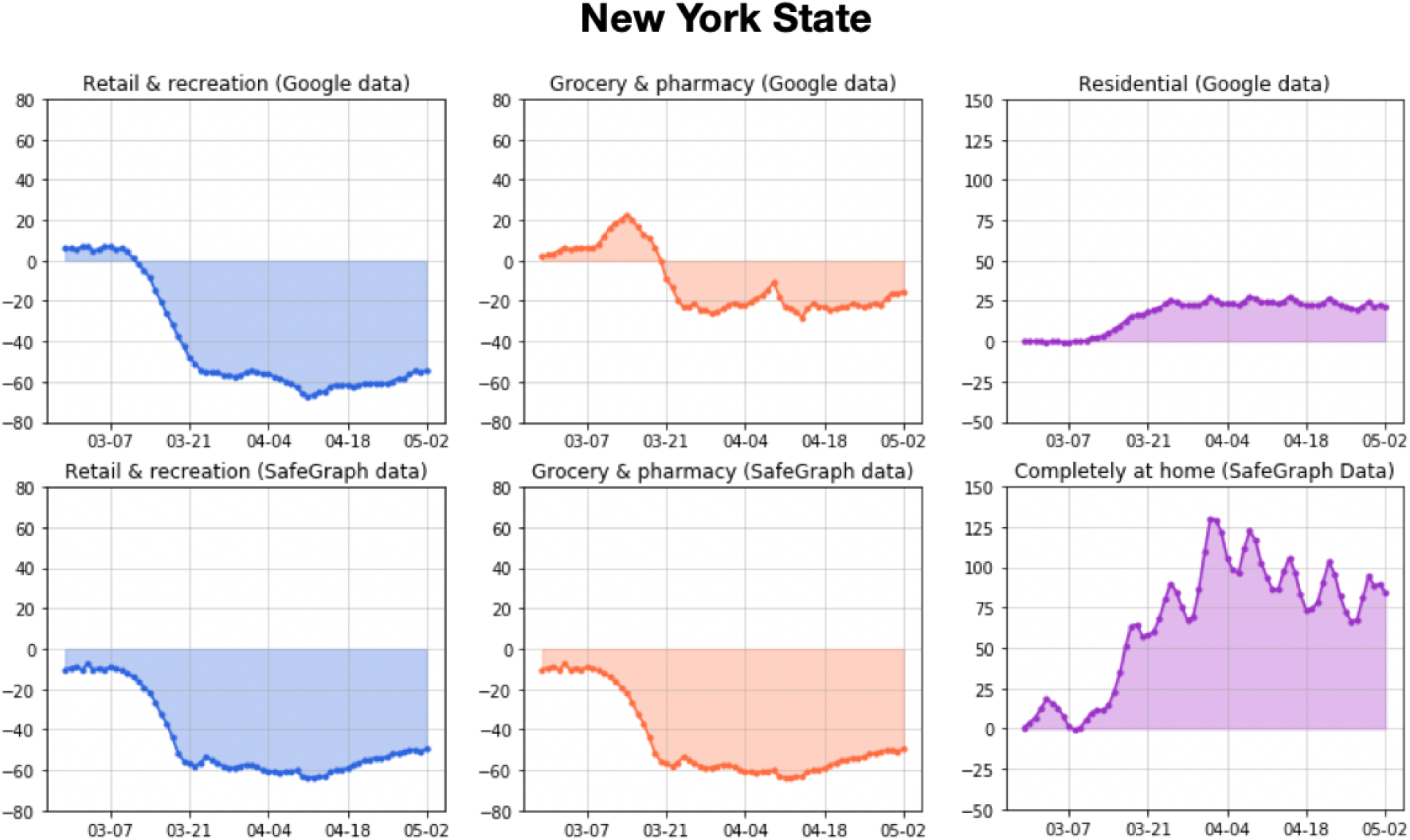
Google versus SafeGraph mobility trends for New York state. The y-axis represents mobility levels compared to baseline activity in January and February 2020. For the categories from left to right, the Pearson correlations between the datasets are 0.96, 0.76, and 0.91. See SI Section S1 for details.

**Figure S4:**
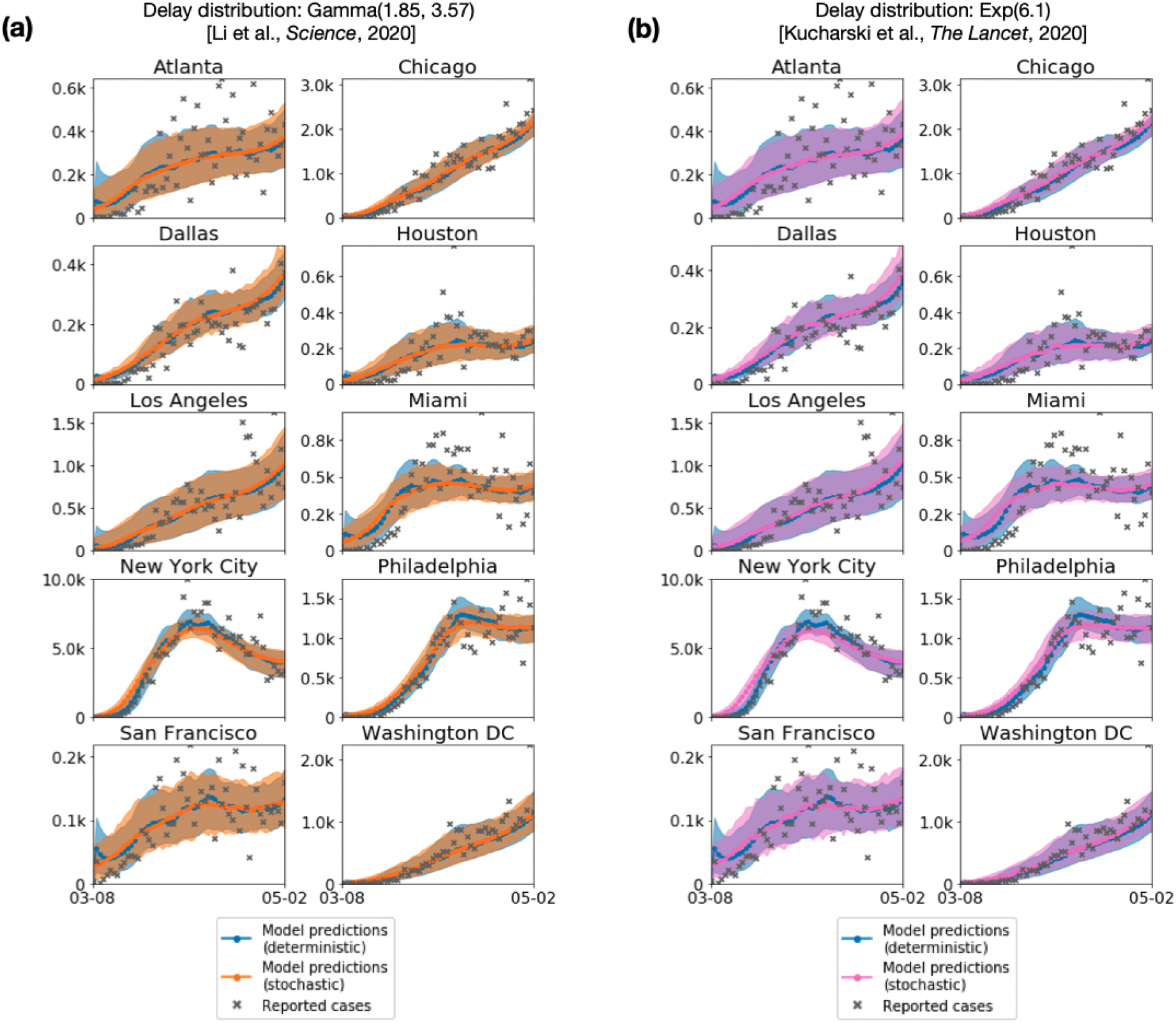
Sensitivity analysis on detection rate and delay. Instead of assuming a constant detection rate and constant infectious-to-confirmation delay on cases, we tested sampling the number of confirmed cases and delay distribution stochastically. The number of confirmed cases was sampled from a Binomial distribution, and we tried two different delay distributions that were fitted on empirical line list data, (a) Li et al. and (b) Kucharski et al. (For more details, see Methods M5.4.) For both delay distributions, we find that model predictions under the stochastic setting are highly similar to the predictions made under the constant rate and delay setting (labeled as “deterministic” in the plot). Note that the “deterministic” and “stochastic” labels only apply to the computation of confirmed cases from infectious individuals to confirmed cases; the underlying SEIR models are all stochastic, as described in Methods M3.

**Figure S5:**
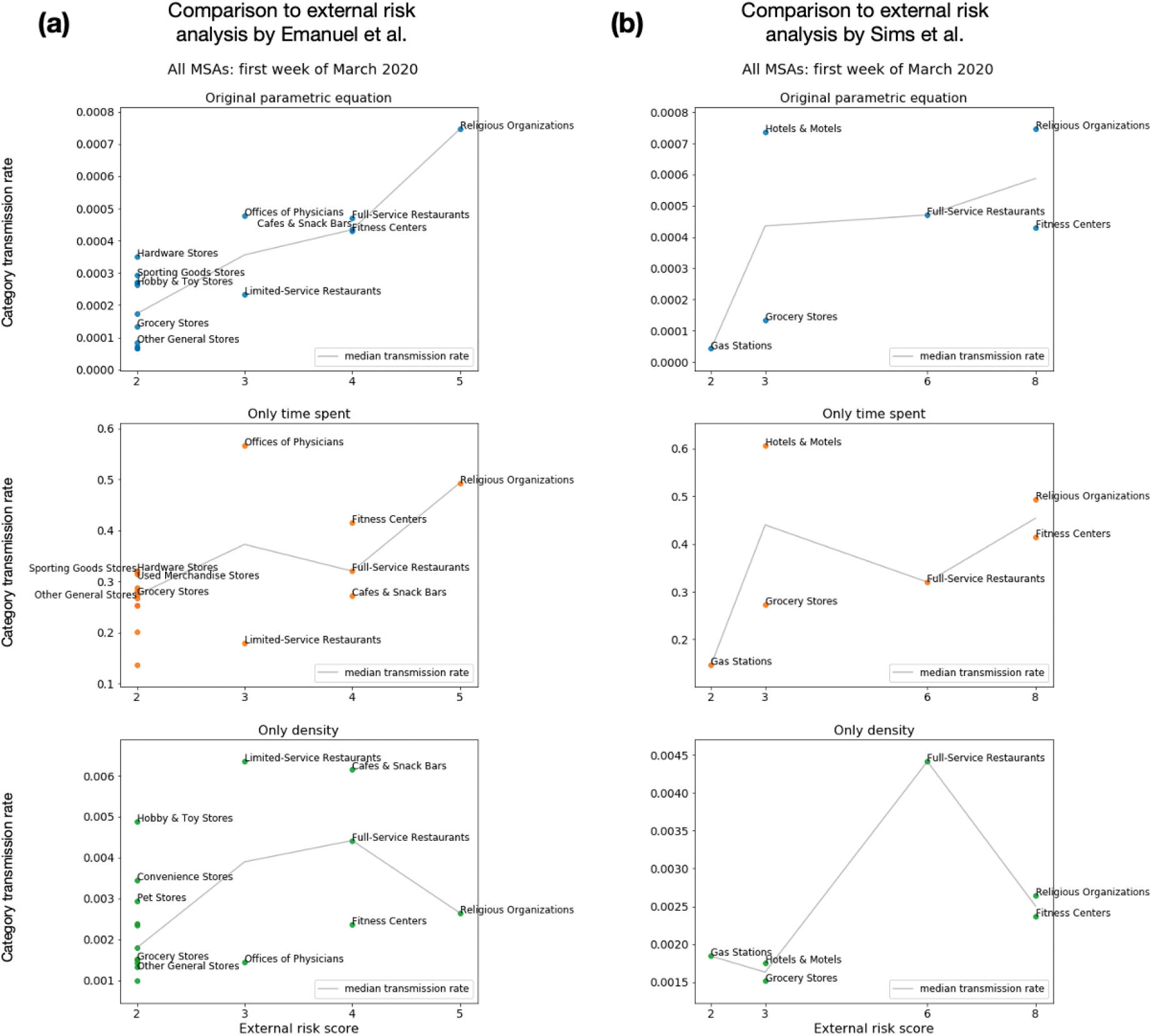
Sensitivity analysis on the parametric form for transmission rate. Our model assumes that POI transmission rates depend on two factors: time spent at the POI and the density of individuals per square foot. We tested this assumption by computing an alternate transmission rate that only included time spent (removing density) and another version that only included density (removing time spent); see Section M5.2 for details. We found that the relative risks predicted by our original transmission rate formula concorded best with the assessments of risk proposed by independent epidemiological experts.^64,65^ The x-axis represents their proposed risk scores; some scores are missing (e.g., 3 and 4 on the right) because there was no overlap between the categories they assigned that score and categories that we analyzed. The y-axis represents each category’s predicted average transmission rate in the first week of March, taking the median over MSAs. Due to space constraints, only a subset of the categories scored at 2 by Emanuel et al. (left) are labeled - the labels are reserved for either the 2 most visited categories in this group (Grocery Stores and Other General Stores) and/or the 3 categories with highest predicted transmission rates within the group.

**Figure S6:**
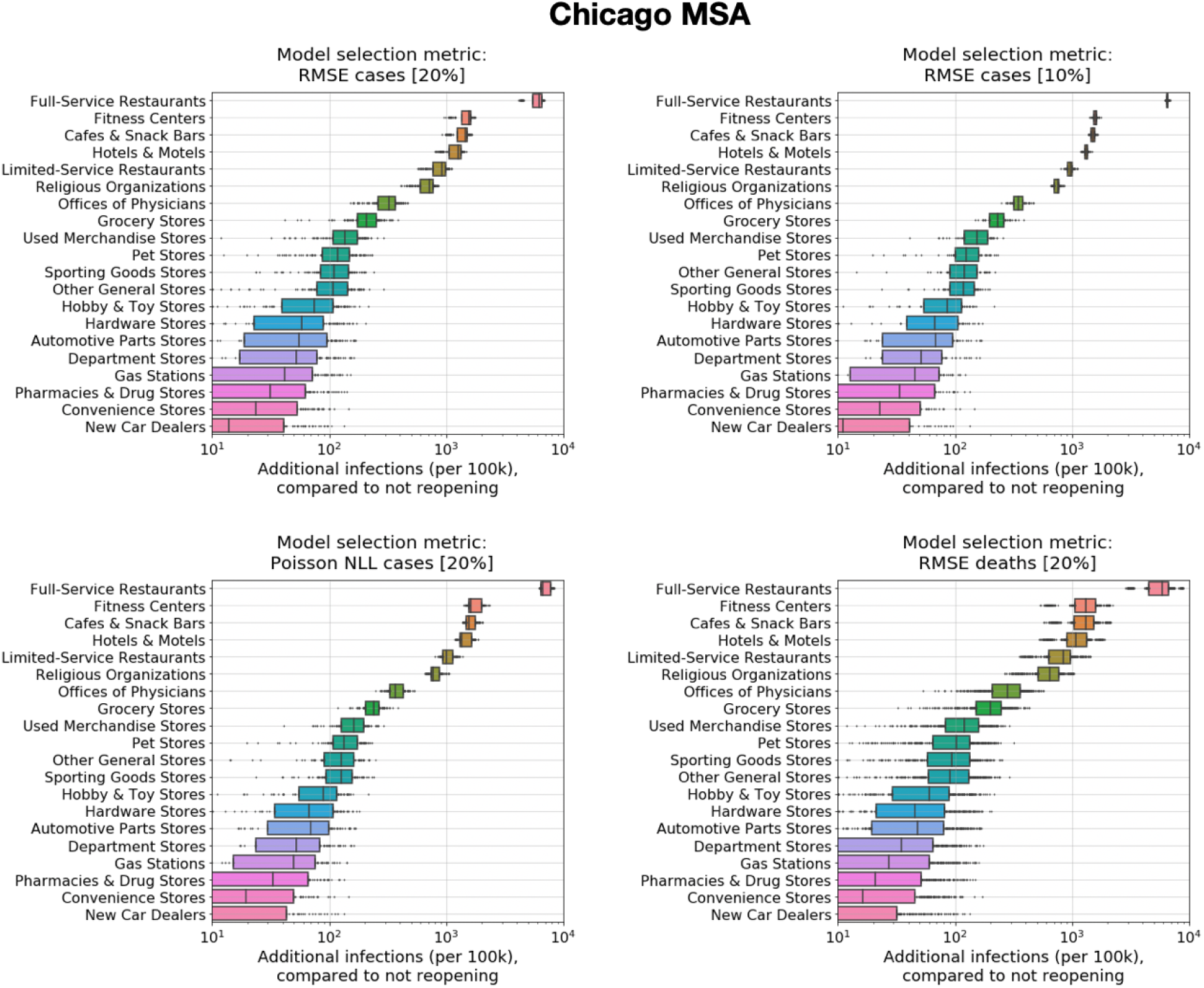
Sensitivity analysis on model calibration metrics and reopening risks. We conducted a sensitivity analysis on which metric was used for model calibration, comparing our default metric (top left) to three other metrics (Methods M5.5). We ran our reopening experiments forward with the model parameters selected by each metric. The predicted ranking of risk from reopening different POI categories remains consistent across all metrics.

**Figure S7:**
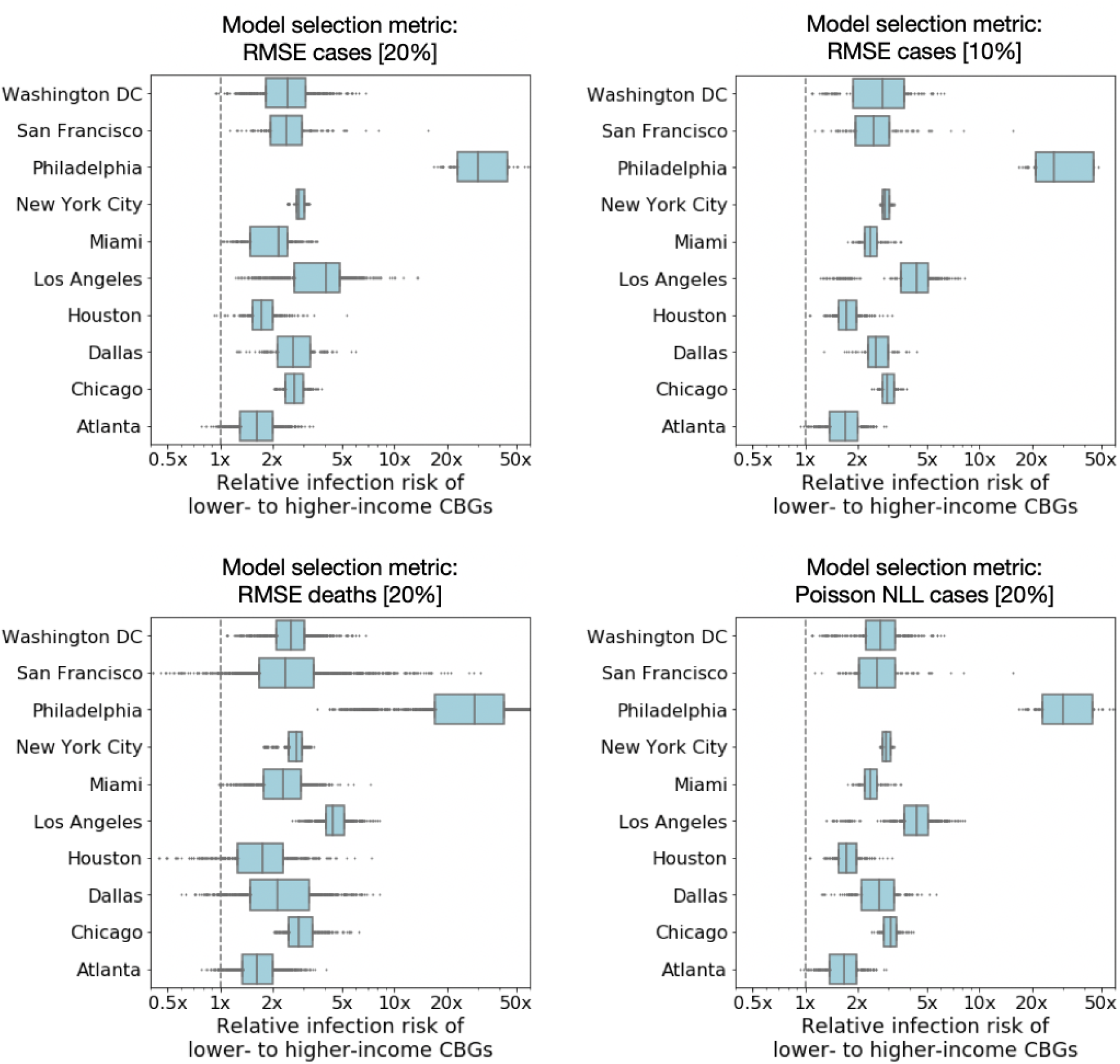
Sensitivity analysis on model calibration metrics and predicted socioeconomic disparities. We conducted a sensitivity analysis on which metric was used for model calibration, comparing our default metric (top left) to three other metrics (Methods M5.5). We then analyzed the socioeconomic disparities in each MSA predicted by the model parameters selected by each metric. The predicted disparities remain remarkably consistent across all metrics, and, for every metric, the best fit models predict that lower-income CBGs are at higher infection risk.

**Figure S8:**
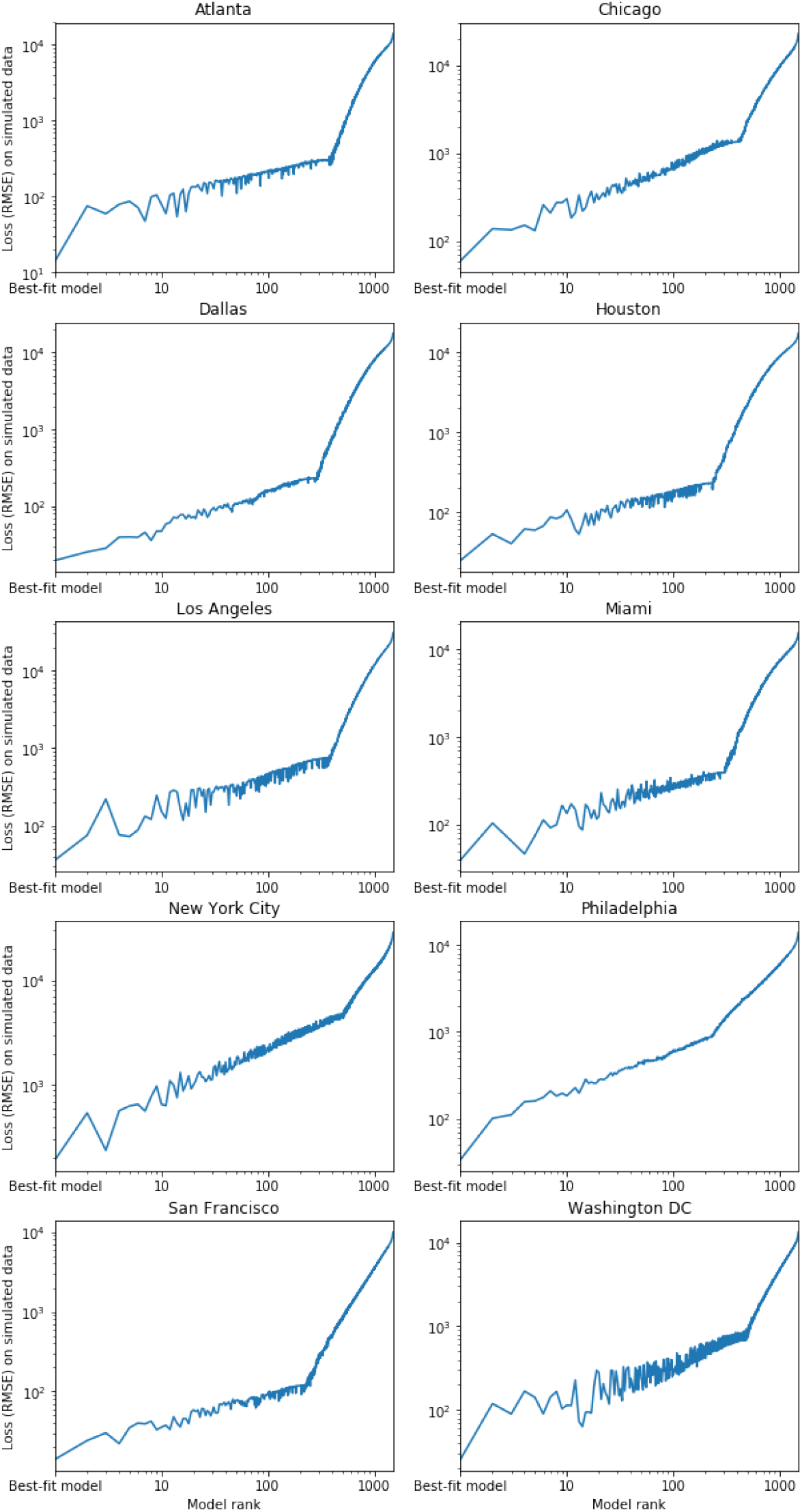
Assessing model identifiability on simulated data. The horizontal axis ranks grid search parameter settings by how well they fit real data (measured by RMSE to daily case count), with the best-fit parameter settings on the left. The vertical axis plots plots RMSE on simulated case count data generated using the best-fit parameter settings. For all 10 MSAs, the parameters that obtain the lowest RMSE on the simulated data are always the true parameters that were used to generate that data (as shown by the left-most point on the plot). This demonstrates that the model and fitting procedure can correctly recover the true parameters on simulated data. Methods M5.3 provides more details.

**Figure S9:**
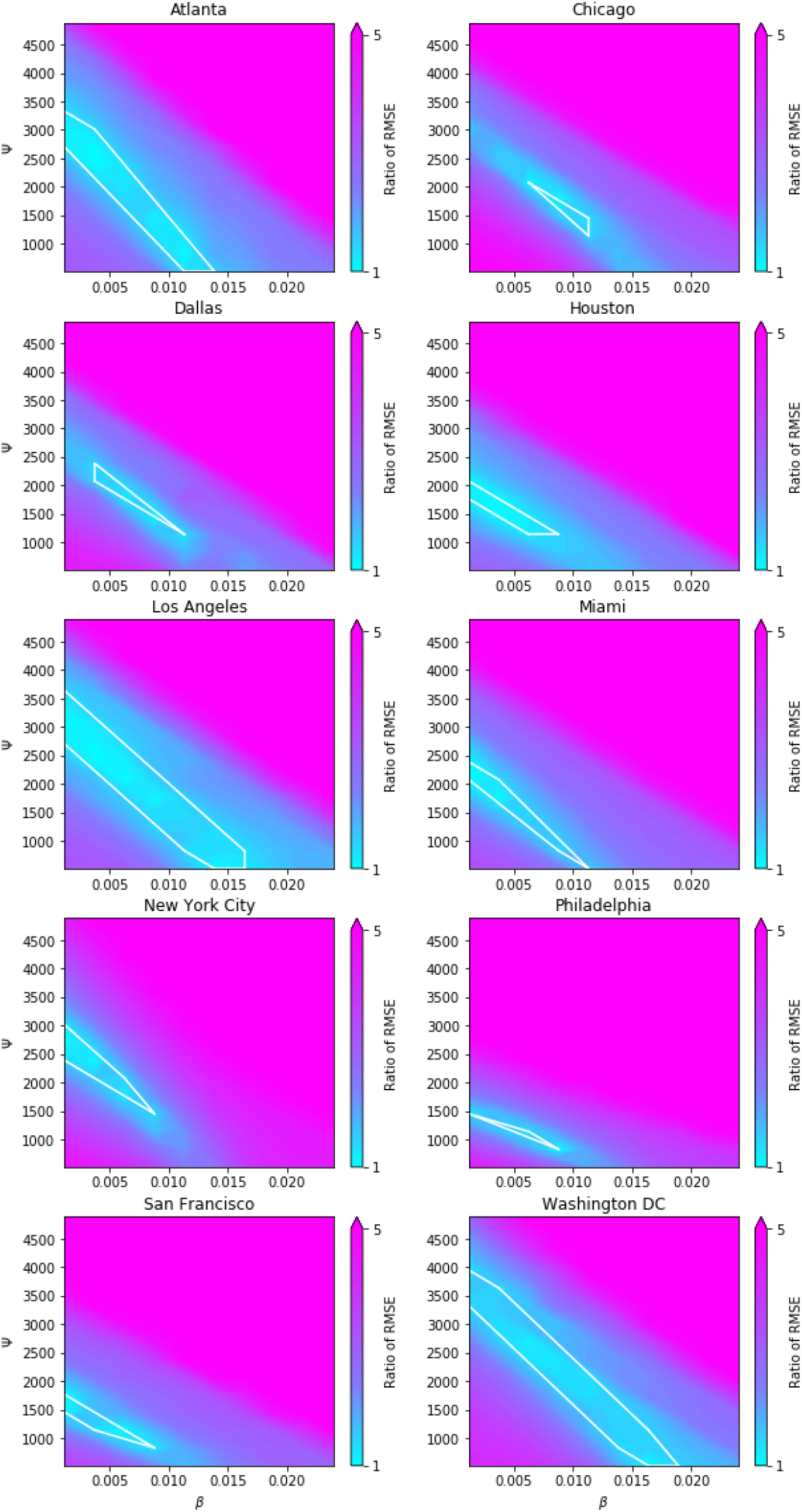
RMSE on daily case count data as a function of parameters *β*_base_ (horizontal axis) and *ψ* (vertical axis). Color indicates the ratio of RMSE to that of the best-fit model. The white polygon shows the convex hull of the parameter settings used to generate results: i.e., all models with an RMSE less than 1.2× that of the best-fit model. For all parameter combinations, we take the minimum RMSE over *p*_0_. Methods M5.3 provides more details.

**Figure S10:**
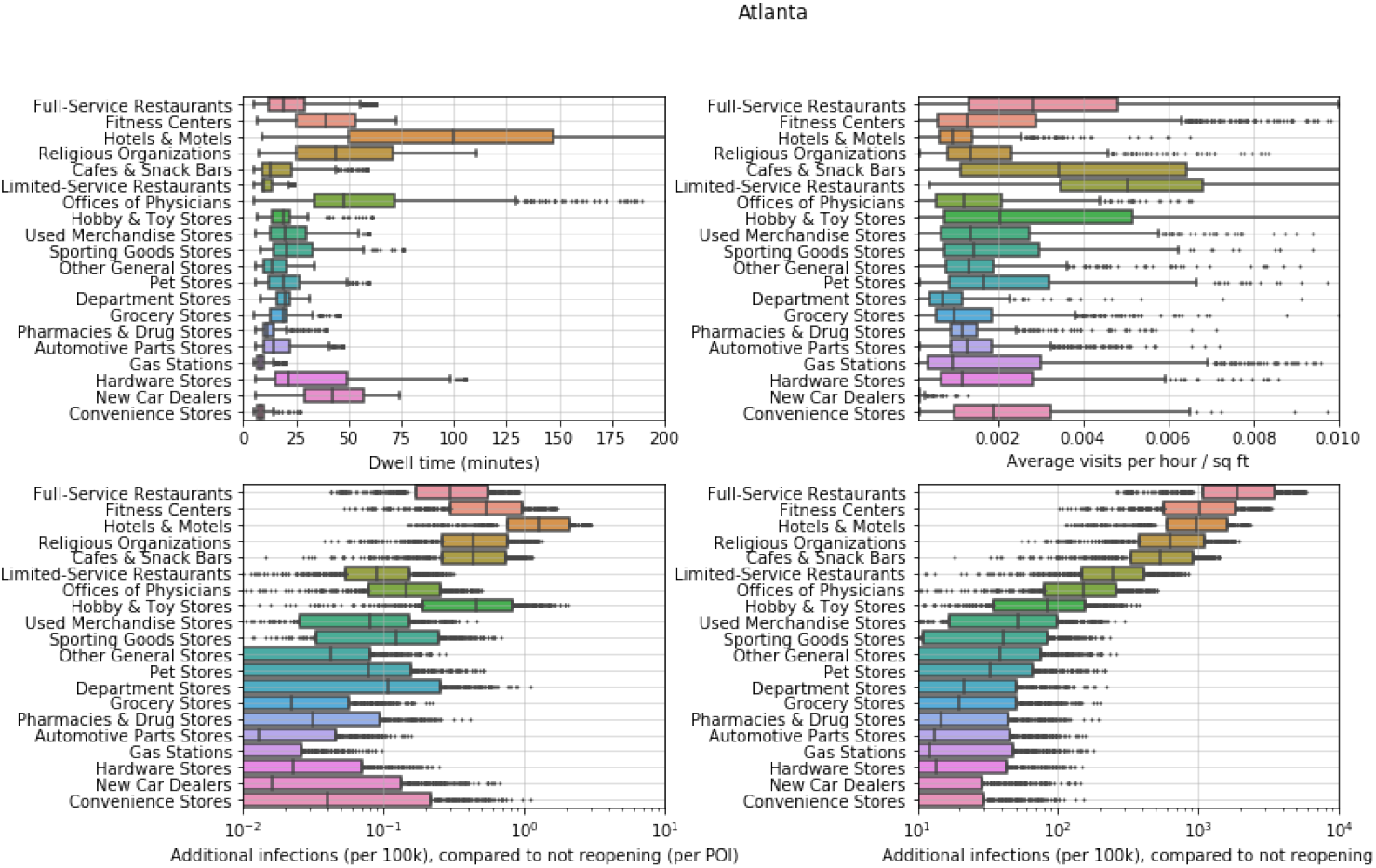
POI attributes in Atlanta. The top two plots show the distribution of dwell time and the average number of hourly visitors divided by the area of the POI in square feet. Each point represents one POI; boxes depict the interquartile range across POIs. The bottom two plots show predictions for the increase in infections (per 100,000 people) from reopening a POI category: per POI (left bottom) and for the category as a whole (right bottom). Each point represents one model realization; boxes depict the interquartile range across sampled parameters and stochastic realizations.

**Figure S11:**
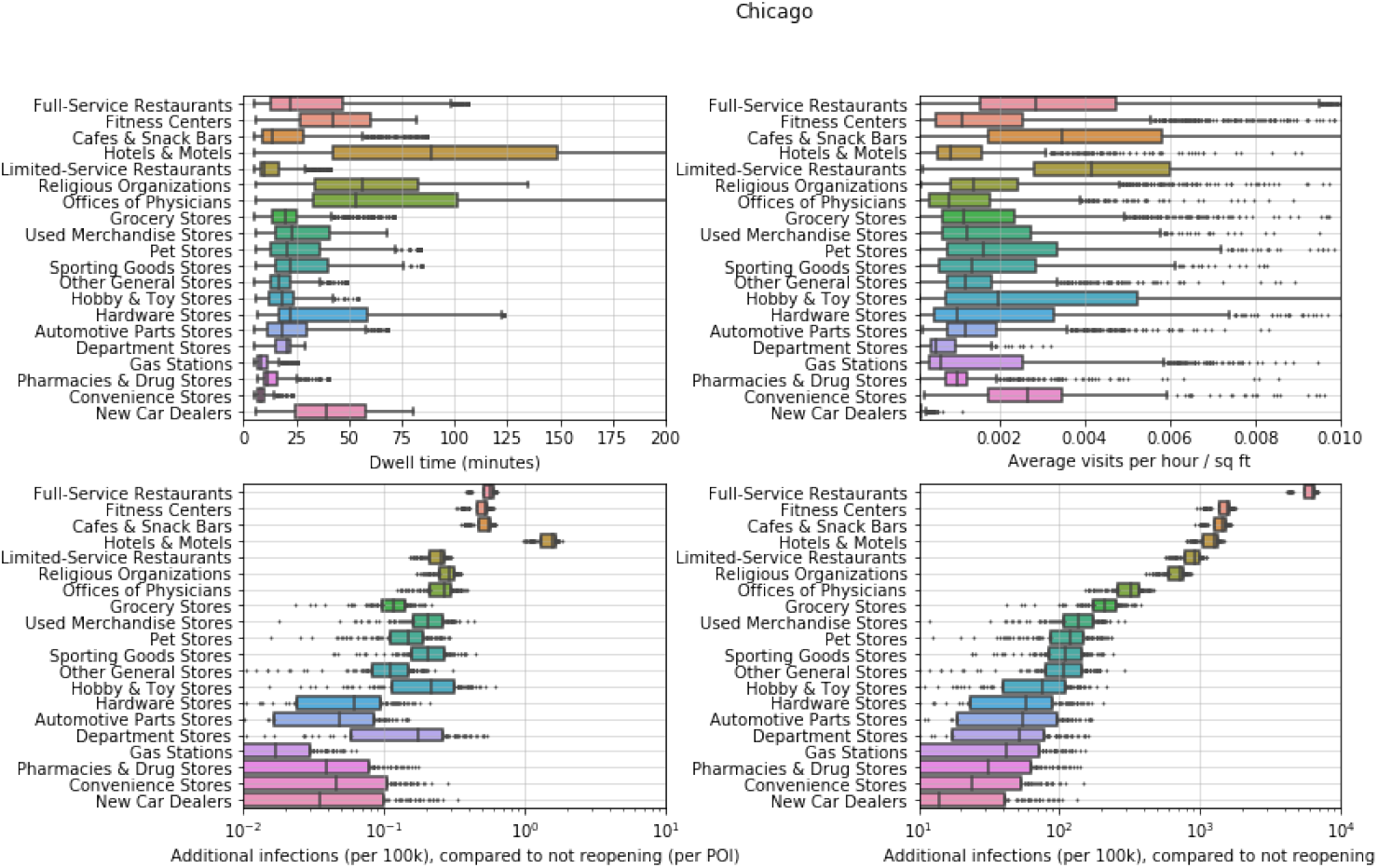
POI attributes in Chicago. See Figure S10 for details.

**Figure S12:**
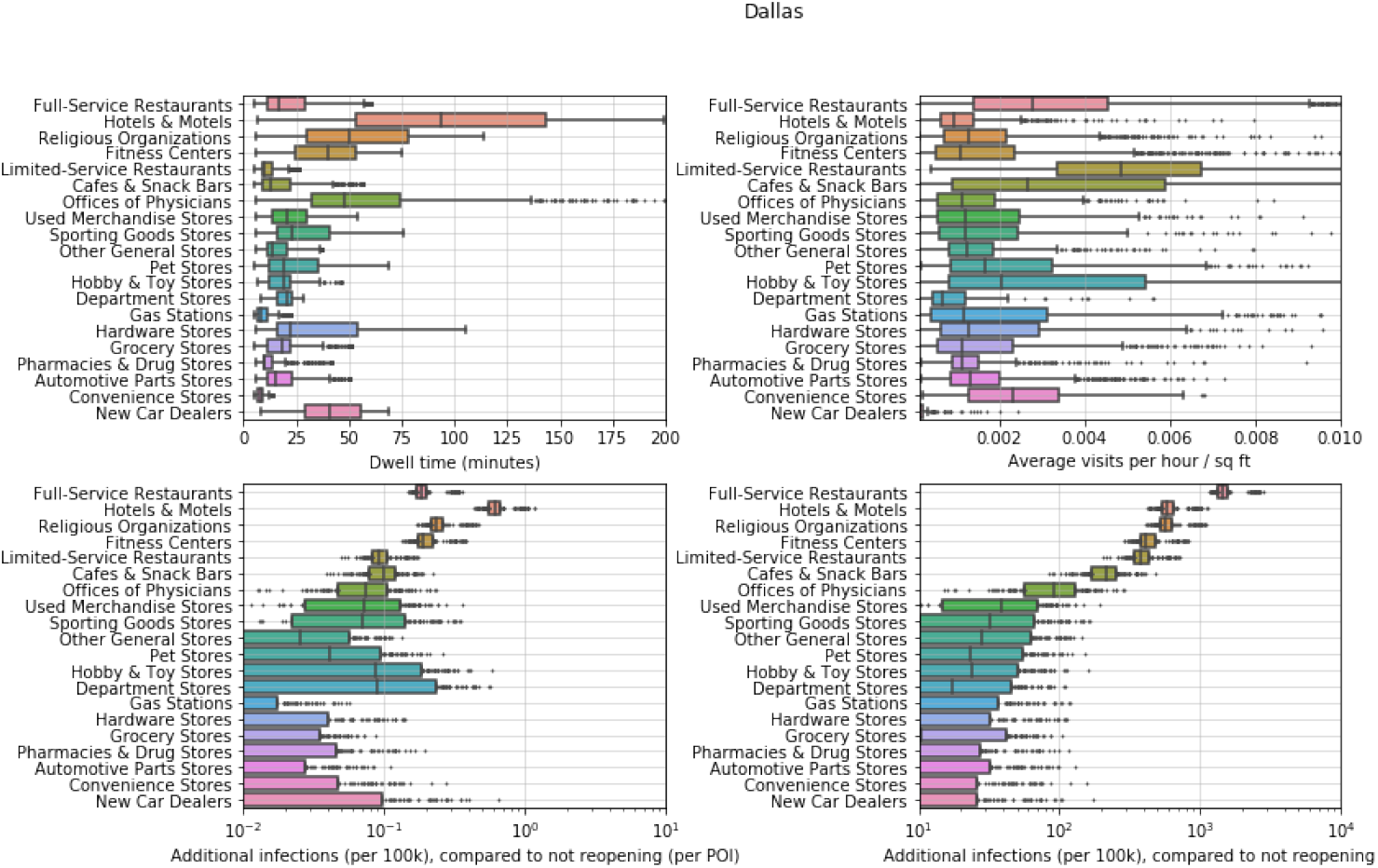
POI attributes in Dallas. See Figure S10 for details.

**Figure S13:**
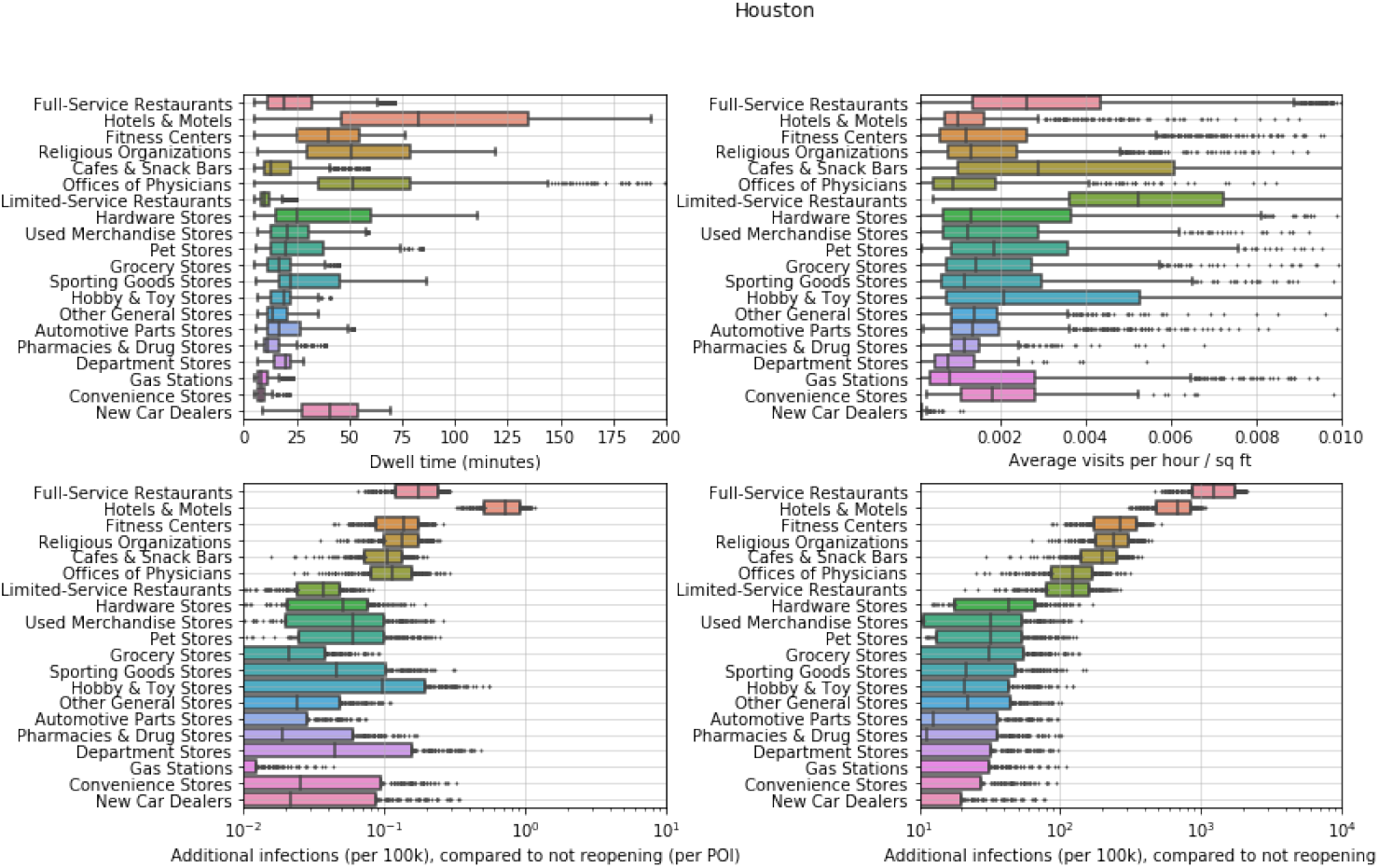
POI attributes in Houston. See Figure S10 for details.

**Figure S14:**
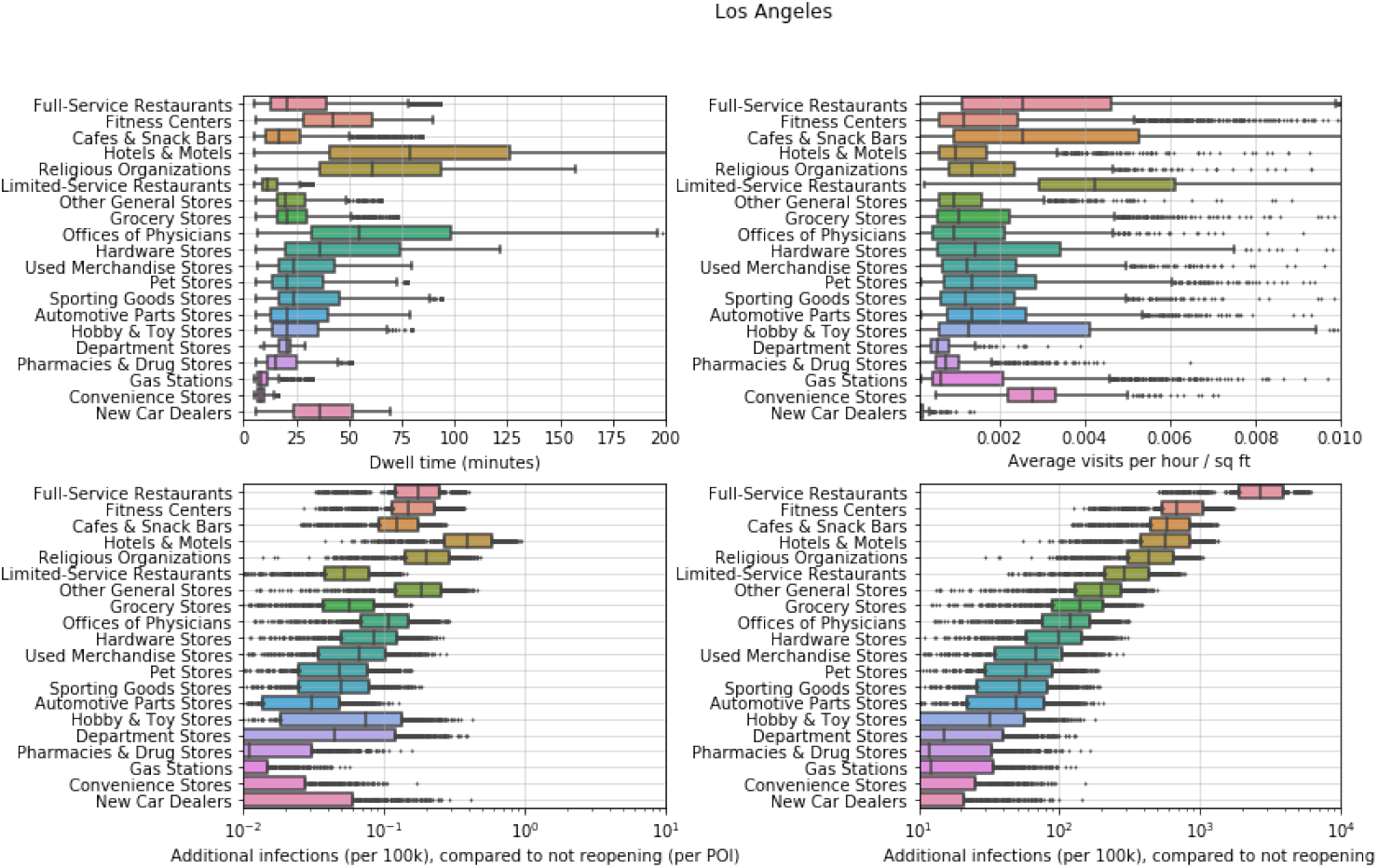
POI attributes in Los Angeles. See Figure S10 for details.

**Figure S15:**
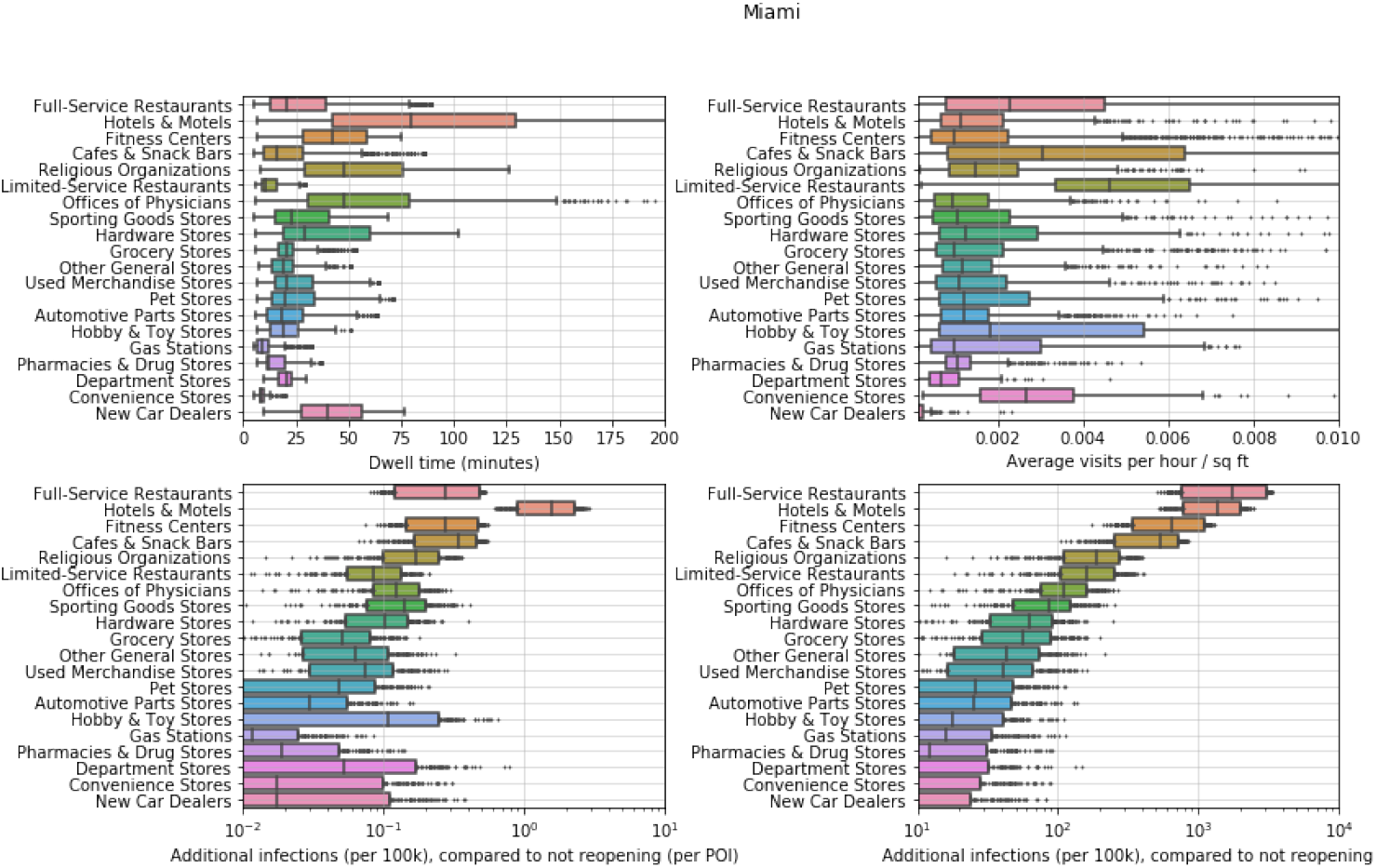
POI attributes in Miami. See Figure S10 for details.

**Figure S16:**
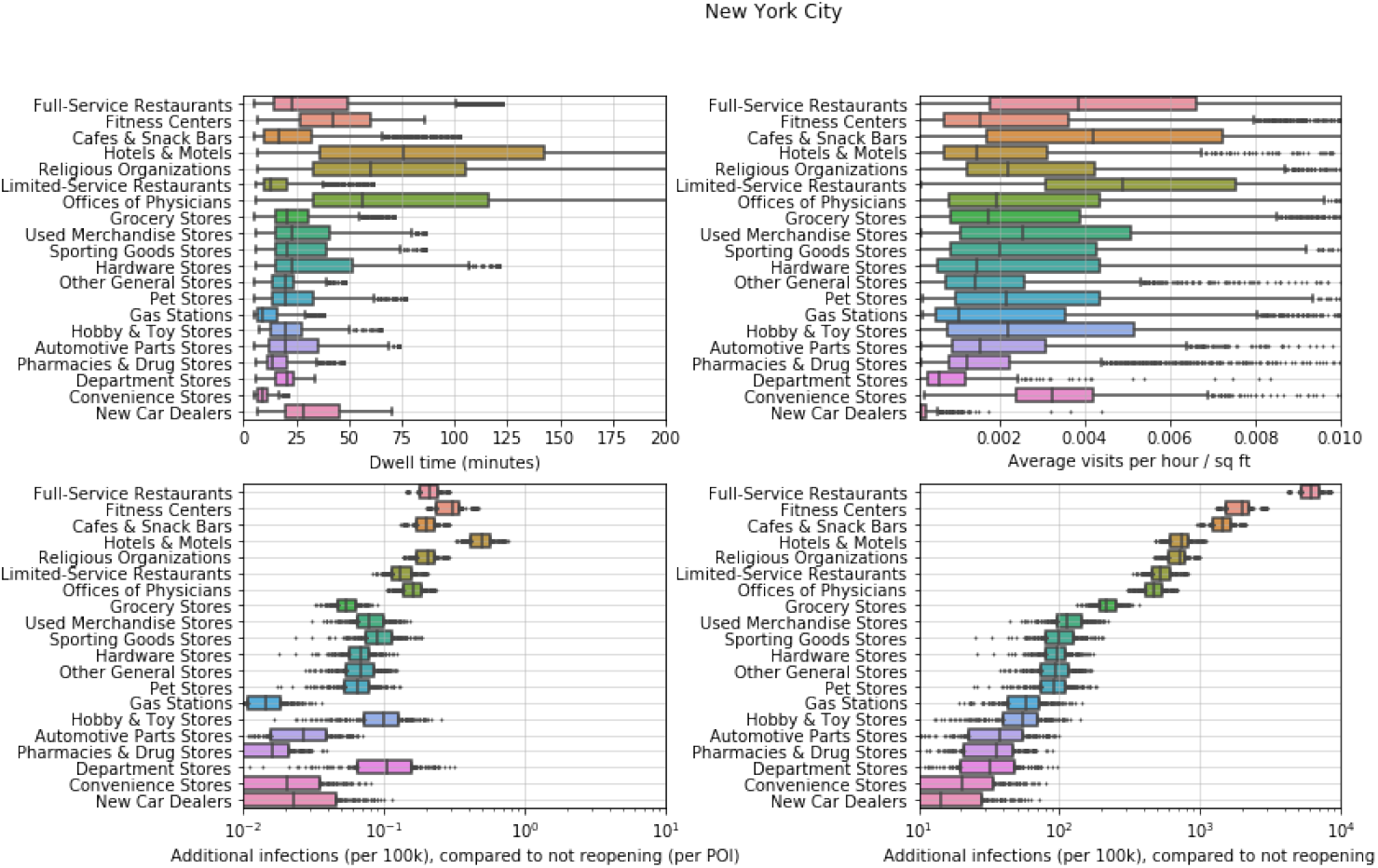
POI attributes in New York. See Figure S10 for details.

**Figure S17:**
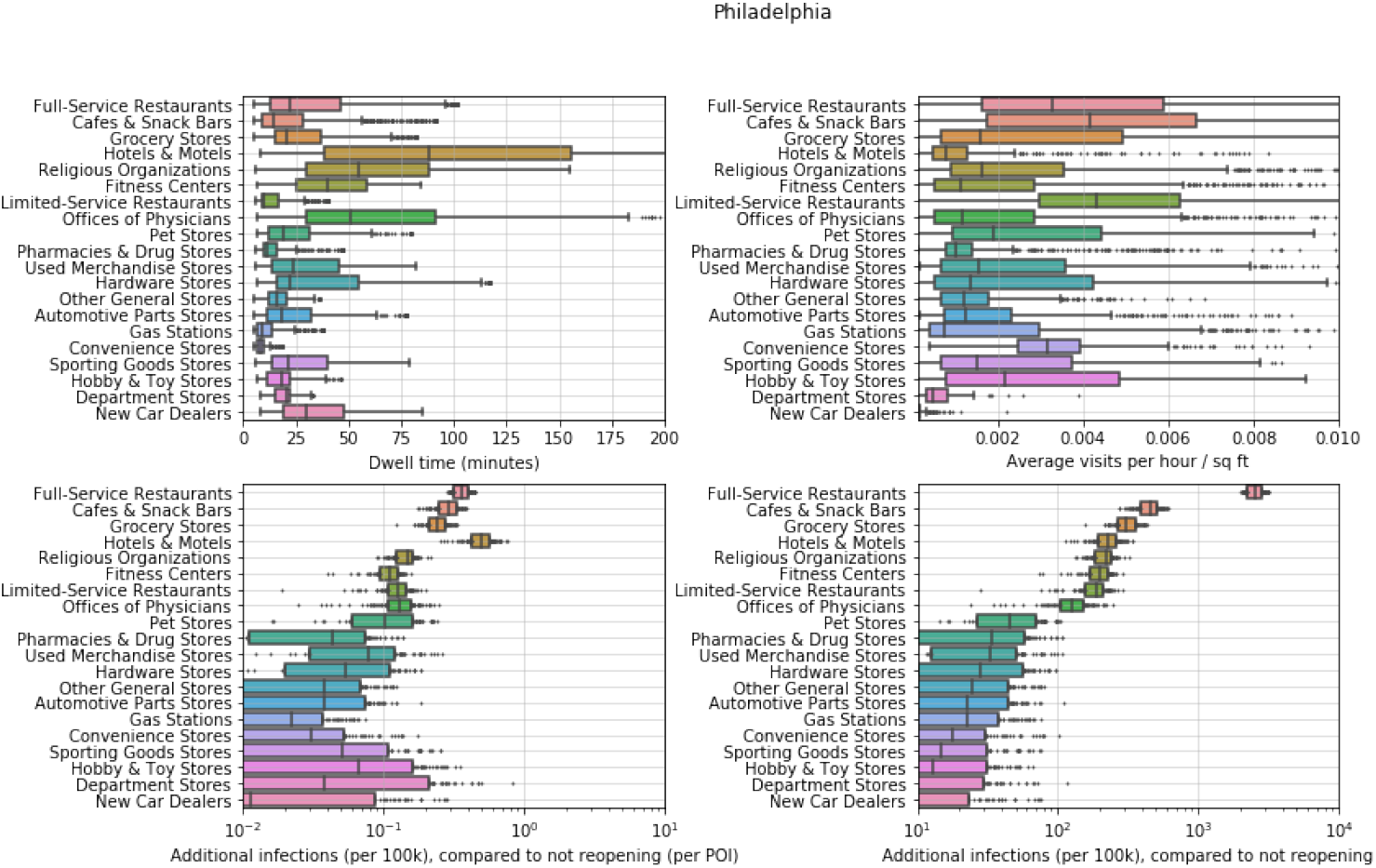
POI attributes in Philadelphia. See Figure S10 for details.

**Figure S18:**
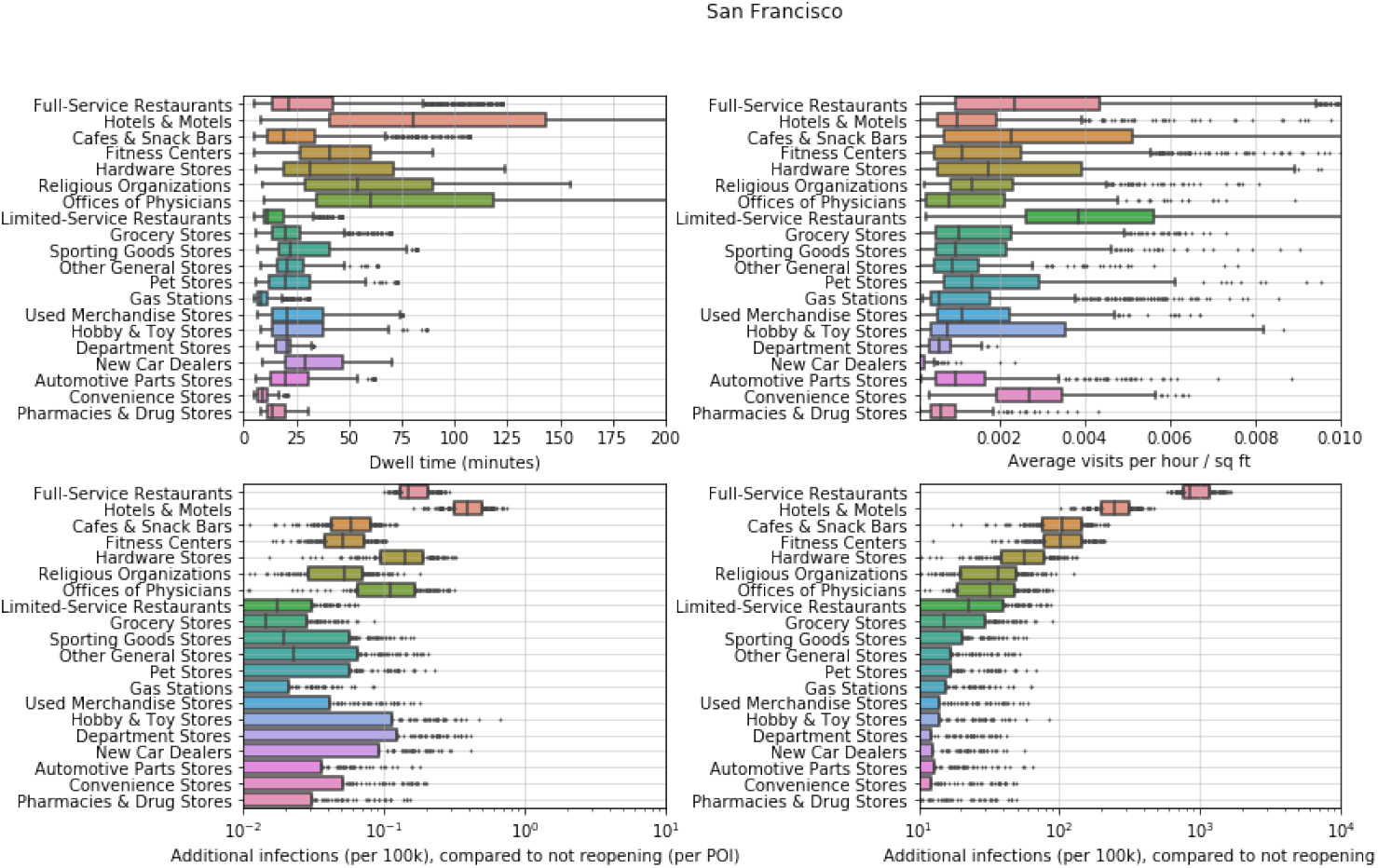
POI attributes in San Francisco. See Figure S10 for details.

**Figure S19:**
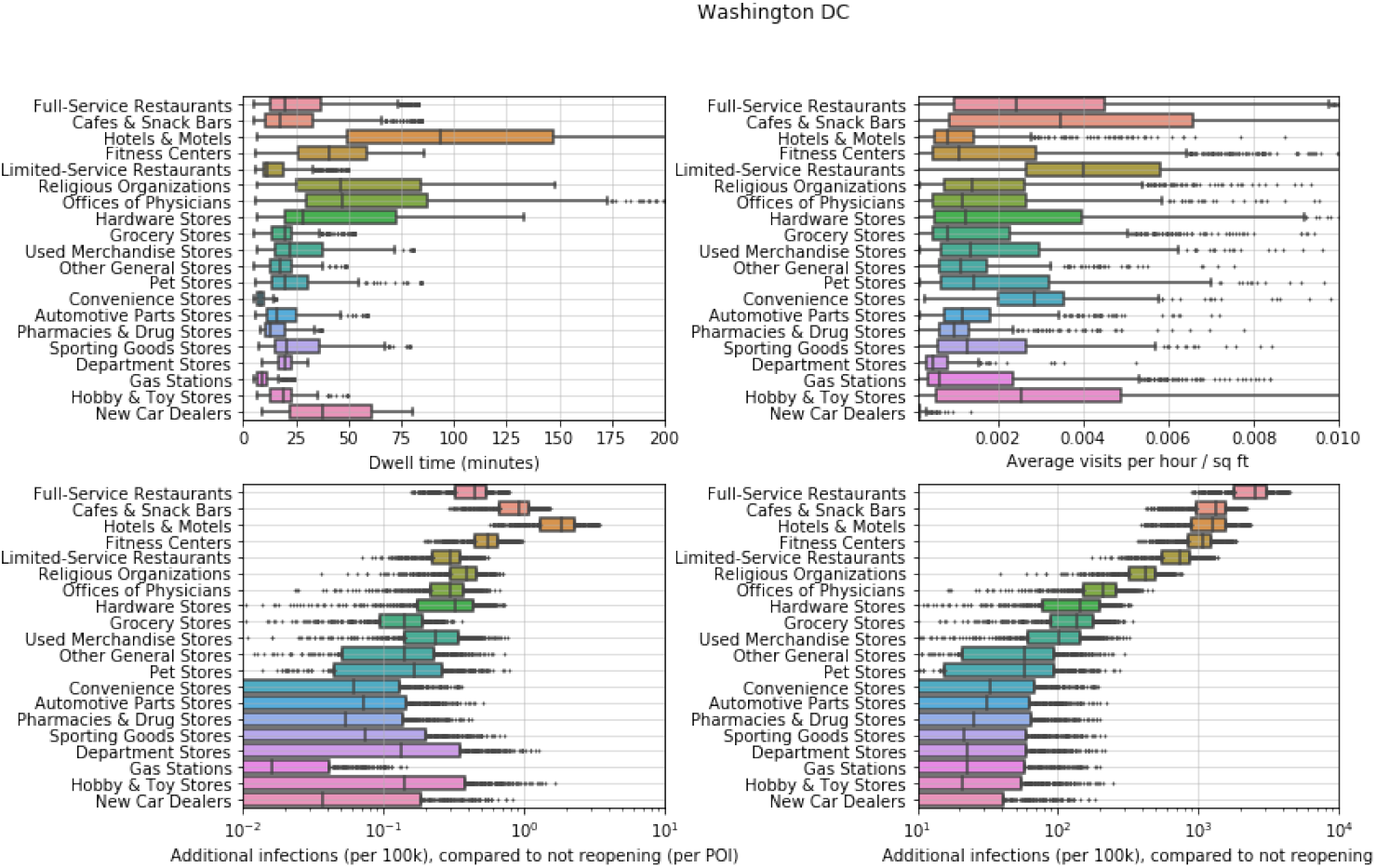
POI attributes in Washington DC. See Figure S10 for details.

